# Scoping review and meta-analysis of COVID-19 epidemiological parameters for modeling from early Asian studies

**DOI:** 10.1101/2022.10.23.22281408

**Authors:** Eva S. Fonfría, M. Isabel Vigo, David García-García, Zaida Herrador, Miriam Navarro, Cesar Bordehore

## Abstract

Retrospective epidemiological models are powerful tools to understand its transmission dynamics and to assess the efficacy of different control measures. This study summarises key epidemiological parameters of COVID-19 for retrospective mathematical and clinical modeling. A review of scientific papers and preprints published in English between 1 January and 15 April 2020 in PubMed, MedRxiv and BioRxiv was performed to obtain epidemiological parameters of the initial stage of COVID-19 pandemic in Asia. After excluding articles with unacceptable risks of bias and those that remained as preprints as of 15 November 2021, meta-analyses were performed to derive summary effect estimates from the data collected using the statistical software R. Out of 4,893 articles identified, 88 provided data for 22 parameters for the overall population and 7 specifically for children. Meta-analyses were conducted considering time period as a categorical moderator when it was statistically significant. The results obtained are essential for building more reliable models to help clinicians and policymakers improve their knowledge on COVID-19 and apply it in future decisions.

## Introduction

Without any doubt the COVID-19, caused by the novel coronavirus SARS-CoV-2, is the most significant global public health threat in recent decades. It was declared a “Public Health Emergency of International Concern” and “pandemic” by the World Health Organization (WHO) in 30 January and 11 March 2020, respectively. Since it was first detected in December 2019 in Wuhan, China, it has spread over 215 countries and territories, with almost 600 million confirmed cases and more than 6,4 million deaths worldwide as of 21 August 2022 ^1^, although these figures are likely underestimated ^2–4^.

Currently, preventive therapeutic strategies are being implemented worldwide, to a greater or lesser extent depending on the country ^5^. Although vaccines are effective in reducing infection and contributing to community protection by reducing the likelihood of virus transmission, their effectiveness varies among the different variants of SARS-COV-2 that have emerged ^6,7^. In this context, where the appearance of the Omicron variant has once again placed restrictions and non-pharmaceutical actions on the agenda of many governments ^8^, retrospective epidemiological modeling may allow us to gain understanding on the chronology of the epidemic progression, the effectiveness of different control measures, etc. ^4,9,10^ which can be very useful in trying to contain ongoing and future outbreaks.

Since the beginning of the pandemic, thousands of preprints and papers have been published about COVID-19. Given the high volume of existing literature and the heterogeneity of the studies on COVID-19, it has been very difficult for researchers to agree on the most accurate parameters. Additionally, most studies have examined only few variables, making data mining particularly onerous and tedious. A broader perspective on generalised values for the key parameters would be extremely useful for improving retrospective epidemiological models and the knowledge of past decisions and events, which could help future decisions. A meta-analysis is the best tool to systematically assess the results of previous research to derive conclusions. Thus, the aim of this study was to identify and summarise key epidemiological parameters of the initial stages of COVID-19 from the vast existing literature through meta-analysis in order to make that scattered information useful for retrospective mathematical modeling and also, when data are available, for comparative analysis with other pandemic stages, where new variants predominate over the initial one.

## Results

During the study period, 6,969 scientific articles related to COVID-19 were published or posted in the consulted libraries. Those were filtered using the keywords and terms described in the method section and the titles and abstracts screened, leaving the full text of 361 papers to review. Of those, 131 contained extractable data (see Supplementary Table S1), but only 88 were considered in our meta-analysis for one or more parameters (see Figure 1, the PRISMA flow diagram, for details in exclusion criteria). Unacceptable risk of bias was detected in 5 studies, mainly due to unclear selection approach, and insufficient follow-up period (see the complete evaluation in the Supplementary Table S2). As of 15 November 2021, 13 papers remained unpublished, and they were also excluded.

**Fig. 1.**
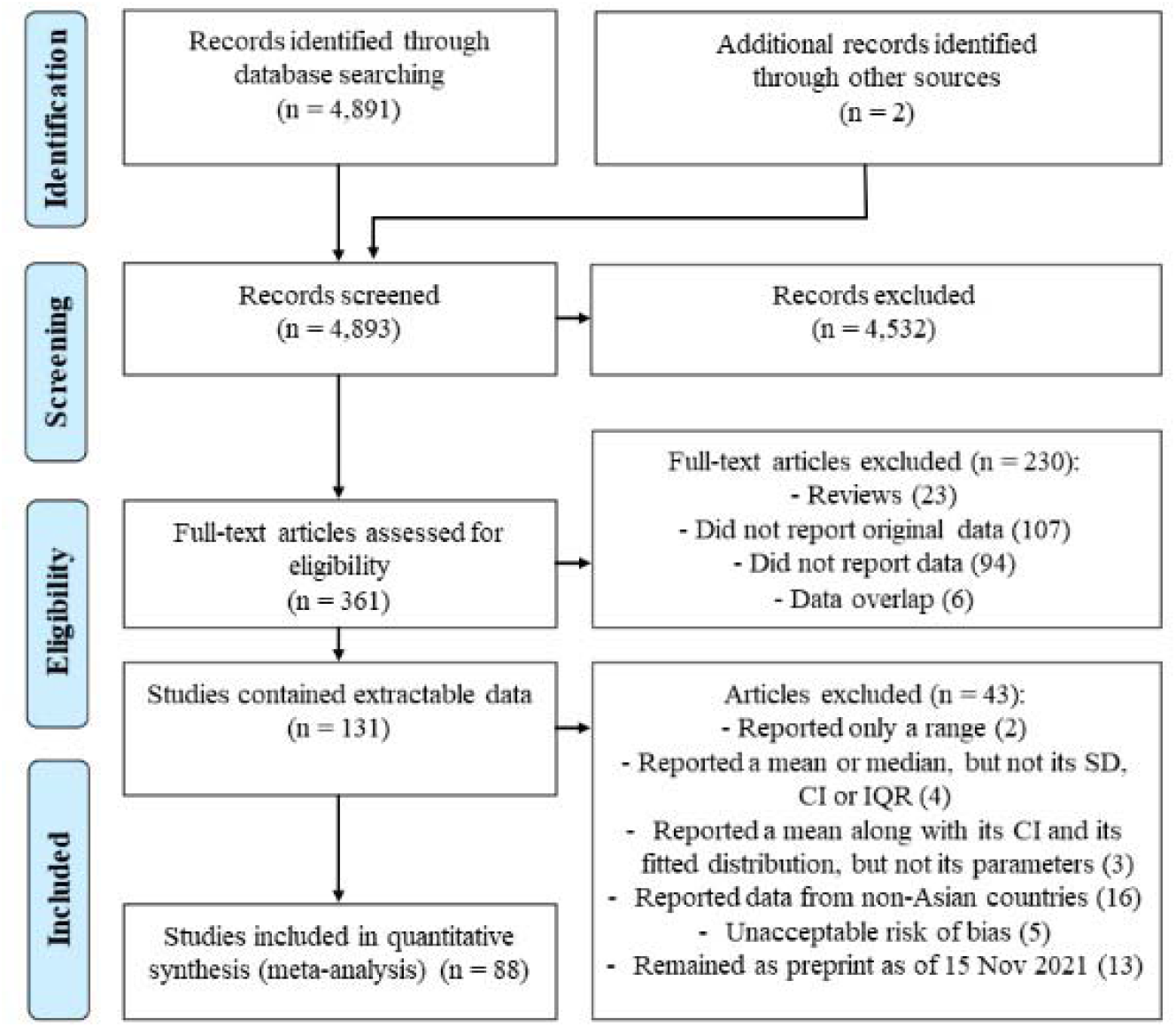
PRISMA flow diagram. Abbreviations: SD, Standard deviation; CI, Confidence Intervals; IQR, Interquartile range.

Pooled means or percentages and their associated 95% CI were determined for 22 epidemiological parameters for the general population (disaggregated into survivor/non-survivor when possible) and 7 for children. Tables 1 and 2 show the results provided by the meta-analysis for those parameters for which, having performed the meta-regression using the period of admission of the patients in the hospital as a categorical moderator, the results were not significant and those that did, respectively.

**Table 1.** Meta-analysis results for parameters where period is not a significant moderator.

| Parameter | All studies included |  |  |  |  | Outliers removed |  |
| --- | --- | --- | --- | --- | --- | --- | --- |
|  | Mean | 95% CI | Studies included | N (patients) | I<sup>2</sup> (%) | Mean | I<sup>2</sup> (%) |
| **General Population** |  |  |  |  |  |  |  |
| **Serial interval (days)** | 4.97 | 3.88-6.36 | 7 | 662 | 91 | 5.80 | 18 |
| **Incubation period (days)** | 5.61 | 4.74-6.63 | 16 | 2437 | 96 | 5.30 | 52 |
| **OS to hospital admission (all)(days)** | 6.29 | 5.32-7.44 | 27 | 4323 | 98 | 6.51 | 12 |
| **OS to hospital admission (non-survivor) (days)** | 10.15 | 9.61-10.71 | 7 | 574 | 56 | 10.18 | 47 |
| **OS to hospital ICU admission (all) (days)** | 9.74 | 9.20-10.31 | 6 | 187 | 0 | ND | ND |
| **Hospital stay length (non-survivor) (days)** | 8.63 | 7.23-10.31 | 12 | 785 | 93 | 8.12 | 0 |
| **ICU stay length (survivor)(days)** | 9.08† | 6.47-12.76 | 4 | 212 | 92 | NA | NA |
| **OS to death (days)** | 17.79 | 16.06-19.71 | 10 | 516 | 86 | 16.66 | 57 |
| **OS to discharge (days)** | 21.76† | 18.13-26.12 | 3 | 362 | 98 | NA | NA |
| **Pre-symptomatic transmission (%)** | 29.33 | 11.29-57.50 | 5 | 870 | 97 | ND | ND |
| **Asymptomatic patients (%)** | 8.54 | 4.04-17.18 | 14 | 2430 | 90 | 5.44 | 18 |
| **ICU admissions vs all hospital admissions (%)** | 13.09 | 9.14-18.41 | 19 | 4994 | 94 | 16.75 | 40 |
| **Deaths vs all hospitalized patients (%)** | 7.62 | 4.59-12.41 | 20 | 5771 | 94 | 13.23 | 41 |
| **Deaths from ICU (%)** | 35.03 | 19.42-54.67 | 5 | 453 | 79 | 38.30† | 0 |
| **Discharged vs all hospitalized patients (%)** | 49.34 | 36.01-62.76 | 25 | 4840 | 97 | 34.93 | 0 |
| **Discharged from ICU (%)** | 44.96 | 25.73-65.82 | 6 | 487 | 85 | 54.61† | 2 |
| **In hospital (%)** | 59.79 | 47.01-71.36 | 21 | 3902 | 97 | 63.13 | 21 |
| **In hospital (ICU) (%)** | 16.63 | 8.53-29.91 | 7 | 537 | 86 | 16.30 | 57 |
| **Children (<18years)** |  |  |  |  |  |  |  |
| **Incubation period (days)** | 6.69† | 5.49-8.15 | 4 | 25 | 0 | NA | NA |
| **OS to diagnosis (days)** | 3.27† | 1.35-7.92 | 3 | 744 | 92 | NA | NA |
| **Hospital stay length (days)** | 12.77† | 8.51-19.17 | 3 | 52 | 90 | NA | NA |
| **Asymptomatic patients (%)** | 16.66† | 11.52-23.48 | 4 | 945 | 56 | NA | NA |
| **ICU admissions vs all hospital admissions (%)** | 5.06† | 1.64-14.60 | 4 | 211 | 42 | NA | NA |
| **Discharged vs all hospitalized patients (%)** | 19.39† | 3.39-62.23 | 3 | 205 | 86 | NA | NA |
| **In hospital (%)** | 80.27† | 37.87-96.45 | 3 | 205 | 86 | NA | NA |

Abbreviations: CI, Confidence Intervals; ICU, Intensive Care Unit; NA, Not Applicable; ND, Not
Detected; OS, Onset of symptoms.
† Results are based on fewer than 5 studies.

**Table 2.**
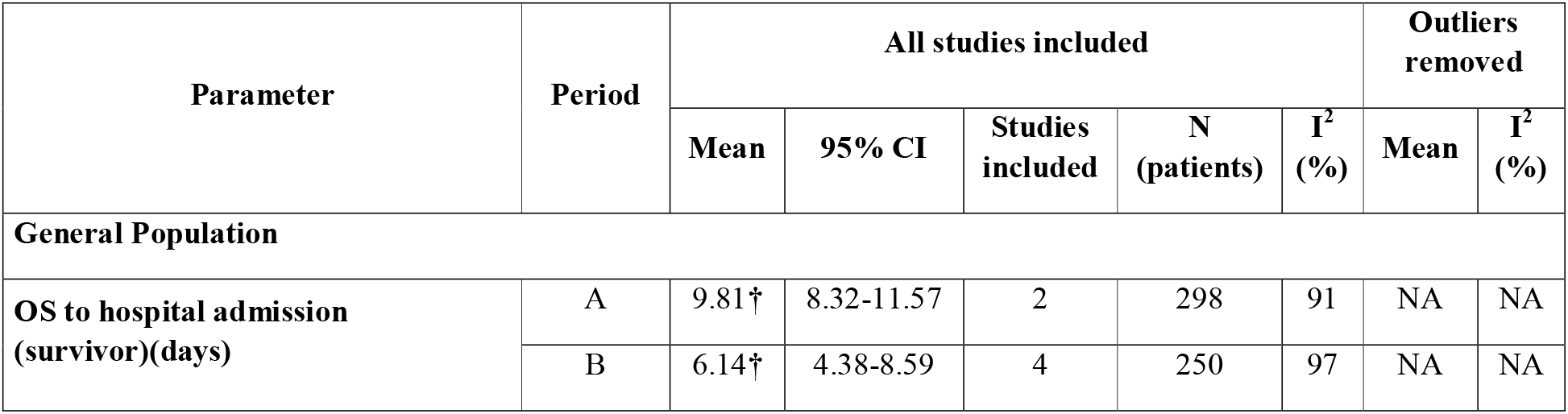
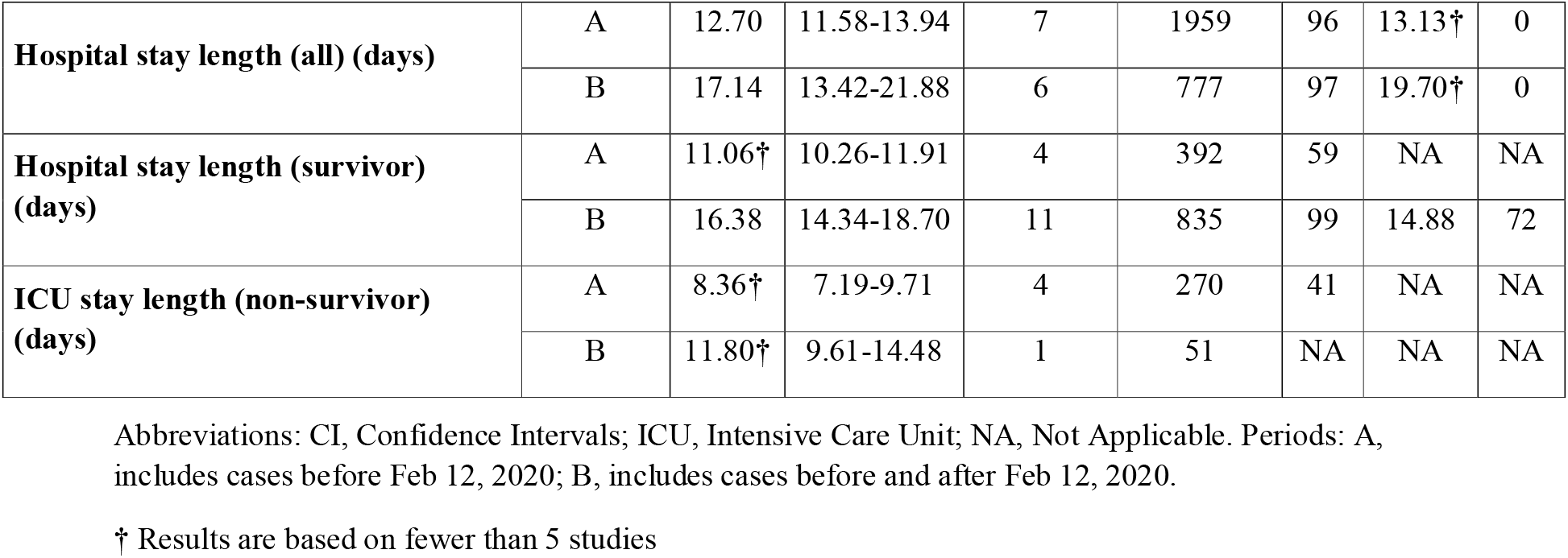
Meta-analysis results for parameters where period is a significant moderator.

| Parameter | Period | All studies included |  |  |  |  | Outliers removed |  |
| --- | --- | --- | --- | --- | --- | --- | --- | --- |
|  |  | Mean | 95% CI | Studies included | N (patients) | I<sup>2</sup> (%) | Mean | I<sup>2</sup> (%) |
| General Population |  |  |  |  |  |  |  |  |
| OS to hospital admission (survivor)(days) | A | 9.81† | 8.32-11.57 | 2 | 298 | 91 | NA | NA |
|  | B | 6.14† | 4.38-8.59 | 4 | 250 | 97 | NA | NA |
| **Hospital stay length (all) (days)** | A | 12.70 | 11.58-13.94 | 7 | 1959 | 96 | 13.13<sup>†</sup> | 0 |
|  | B | 17.14 | 13.42-21.88 | 6 | 777 | 97 | 19.70<sup>†</sup> | 0 |
| **Hospital stay length (survivor) (days)** | A | 11.06<sup>†</sup> | 10.26-11.91 | 4 | 392 | 59 | NA | NA |
|  | B | 16.38 | 14.34-18.70 | 11 | 835 | 99 | 14.88 | 72 |
| **ICU stay length (non-survivor) (days)** | A | 8.36<sup>†</sup> | 7.19-9.71 | 4 | 270 | 41 | NA | NA |
|  | B | 11.80<sup>†</sup> | 9.61-14.48 | 1 | 51 | NA | NA | NA |

Abbreviations: CI, Confidence Intervals; ICU, Intensive Care Unit; NA, Not Applicable. Periods: A, includes cases before Feb 12, 2020; B, includes cases before and after Feb 12, 2020.
<sup>†</sup> Results are based on fewer than 5 studies

For the general population parameters, the number of studies considered ranged from 3 to 27 (median 8.5, Q1 6 and Q3 15.75) and the number of patients involved ranged from 187 to 5771 (mean 1692), whereas for children studies ranged from 3 to 4 (median 3, Q1 3 and Q3 4) and patients from 25 to 945 (mean 341). The estimated I^2^, τ^2^, and Q (values summarised in the Supplementary Meta-analysis Report for each parameter) indicated that high heterogeneity was present for most parameters (except for *onset of symptoms to hospital ICU admission*) which reduced their usefulness. The last two columns of both tables also show the results of the parameters for the general population that allowed removal of outliers without compromising their utility, i.e. yielding a lower heterogeneity (values summarised in the Supplementary Meta-analysis Report) and maintaining a minimum of 3 included studies. Given these constraints, the number of studies considered ranges from 3 to 12 (median 7, Q1 5 and Q3 9.75), with the parameters with 9 or more studies included: *onset of symptoms to hospital admission (all), hospital stay length (survivor, period B), asymptomatic patients, percentage of ICU admissions, percentage of deaths, percentage of discharged* and *percentage in hospital* at the end of the study. Parameters with fewer than 5 studies comprised *onset of symptoms to hospital admission (survivor), hospital stay length (all patients, both periods), percentage of deaths from ICU* and *percentage of discharged from ICU*. For children, all parameters were initially estimated from fewer than 5 studies and neither the meta-regression with period as moderator nor outlier diagnosis were carried out.

Forest plots for each parameter (considering all studies or group by period if significant differences between the two subsets were detected) are provided in the Supplementary Material (see Supplementary Figures S1-31). In all cases, a summary of the results is provided when all the studies are included in the meta-analysis, and with outliers/influencers removed if any.

## Discussion

We present a review and meta-analysis of essential epidemiological parameters (22 for the general population, 7 for children) of COVID-19 based on Asian studies during the initial stage of the pandemic. Our main goal was to provide a summary of key parameters useful for modeling, and therefore an exhaustive clinical explanation of our results is beyond the scope of this study. Discussion of the most relevant findings follows.

In order to see the influence of the changes in the way of dealing with the virus resulting from the COVID-19 Forum convened by WHO on the key epidemiological parameters we register the time period of the different studies. We classified and, therefore compared, studies with cases occurring prior February 12, 2020 (period A) and studies with patients enrolled before and after (period B). Significant results in the meta-regression considering period as a categorical moderator were obtained for *hospital stay length (all patients), hospital stay length (survivor), ICU stay length (non-survivor) and onset of symptoms to hospital admission (survivor)*(table 2). Note that since period A is included in period B, the differences are more remarkable. The length of hospital stay (both for all patients and for the survivors) were longer in period B than in A, probably because, but not limited to, the shortage of medical resources during period A. On the other hand, results for *ICU stay length (non-survivor) and onset of symptoms to hospital admission (survivor)*, are not conclusive because only one and two studies are available in period B and period A, respectively.

From the parameters analysed in both groups (overall population versus children), results point that children had longer incubation periods, higher percentages of asymptomatic individuals, fewer ICU admissions and shorter hospital stays. This suggests, as found previously ^11^, less effect of the disease on children than in the overall population.

For the overall population, the mean serial interval obtained (4.97 days, 95% CI: 3.88-6.36) when all studies were considered was shorter than the incubation period (5.61 days, 95% CI: 4.74-6.63), supporting the hypothesis of substantial pre-symptomatic transmission of the virus ^12^. Our own results showed that 29% of disease transmission would be pre-symptomatic. Nevertheless, the results of meta-analysis are sensitive to outliers, and when outliers are removed for this parameter, this theory is not supported. This is due to the fact that Du et al. ^13^ was identified as an outlier. Outliers always must be taken with caution, for example, Du et al. conducted a study with a large population (468 out of a total of 662 patients from all the 7 studies) and reported a mean serial interval (3.96 days) lower than the other studies. Thus, when outliers were removed, the number of studies was reduced to 5 with a total of 187 patients considered and the mean serial interval increased to 5.80 days (95% CI: 5.69-5.92). In a meta-analysis any conclusions that are sensitive to outliers must be stated with caution, while those that are robust to outliers are more reliable.

Our study has some limitations. First, only papers published in English were evaluated. Although many papers have been published in Chinese and a few in other languages during the period analysed, papers in English provided a large enough sample. Second, we performed the search strategy only with three libraries until 15 April 2020. Our work might not include all the published data with the selected parameters, but given the large amount of papers that was posted daily, and considering that the initial phase of the pandemic lasted until April 2020 in China and other Asian countries ^14–16^ we took the pragmatic decision to limit the search in this way. Third, data was restricted to Asian countries. We detected a few useful papers from the USA, Italy and Spain, but we decided not include them in the analysis in order to not introduce additional heterogeneity due to cultural (illness is shaped by cultural factors governing perception, labelling, explanation and valuation of the discomforting experience ^17^) and care pathways differences between Eastern and Western countries. Finally, we presented some results based on fewer than 5 studies and results that showed high heterogeneity, all of which also should be interpreted with caution. Outlier analysis were not appropriate for small groups of studies. We only perform a moderated analysis by time period, but considering other characteristics of the studies would also help to identify causes of heterogeneity. Nevertheless, several of the primary studies had incomplete or ambiguous information for some characteristics that could serve as potential moderators of the disease. Thus, we were unable to identify moderators without drastically reducing the number of studies in the meta-analysis. Therefore, we performed our analysis carefully and considering all these difficulties.

In this paper we summarised more epidemiological parameters than other available multi-parameter reviews on COVID-19 ^18–24^. However, most of them use more studies for most of the parameters than us, mainly due to our language and geographic restrictions, as well as the application of risk of bias analysis and preprints exclusion. Since a mathematical model is as good as the data it uses ^25^, we believe that the considerations we have applied are essential to obtain the most reliable and accurate results available for modeling purposes.

In summary, we reviewed and gathered the most relevant epidemiological parameters obtained during early phase of COVID-19 pandemic to date in a single article. Improvement of our understanding of the dynamics of this infectious disease and evaluation of the effectiveness of the adopted intervention measures at initial stages of pandemic are crucial to successfully cope with COVID-19 until new vaccines (or boosters) or antiviral treatments are implemented. These data may be of interest both to modelers, clinicians, managers, and national and regional policymakers when creating and interpreting the retrospective epidemiological models. Similar studies for different stages of the pandemic in several regions would be highly desirable upon data availability, not only to provide a complete picture of the pandemic, but also to be able to identify differences between healthcare systems and their efficiency on dealing with the COVID-19 pandemic, which could also be useful for future pandemics.

## Methods

Following the drafted protocol (freely available in the Open Science Framework repository, https://osf.io/26mvb), we searched PubMed, MedRxiv and BioRxiv for articles and preprints published in English between 1 January and 15 April 2020 with the keywords “COVID-19”, “SARS-CoV-2”, “severe acute respiratory syndrome coronavirus 2”, “2019-nCoV” or “novel coronavirus”, as well as key terms to obtain data about the following parameters (see Supplementary Text S1 for our search strategy and Supplementary Text S2 for definitions and reclassifications):

- *pre-symptomatic transmission period*,
- *serial interval*,
- *incubation period*,
- *onset of symptoms/illness onset to first medical visit*,
- *from first medical visit to diagnosis*
- *from first medical visit hospital admission*
- *onset of symptoms/illness onset to diagnosis*,
- *onset of symptoms/illness onset to hospital admission*,
- *onset of symptoms/illness onset to ICU (Intensive Care Unit) admission*,
- *hospital stay length*,
- *ICU stay length*,
- *hospital admission to death*,
- *hospital admission to discharge*,
- *onset of symptoms/illness onset to death*,
- *onset of symptoms/illness onset to discharge/recovery*,
- *percentage of pre-symptomatic transmissions*,
- *percentage of asymptomatic patients*,
- *percentage of ICU admissions*,
- *percentage of deaths*,
- *percentage of deaths from ICU*,
- *percentage of discharged*,
- *percentage of discharged from ICU*,
- *percentage in hospital*,
- *percentage in hospital from ICU*,
- *percentage transferred from ICU to general hospital wards*.

Preliminary screening by title and abstract identified potential relevant studies. Subsequently, the full texts of those studies were evaluated and their reference lists examined for additional records (Figure 1). Data related to these epidemiological parameters, such as sample size, mean, standard deviation (SD), confidence intervals, median, interquartile range (IQR) and the fitted distribution used in its estimation, when applicable, as well as sociodemographic information (i.e. patient’s age, gender and location) and the period time of the studies, were extracted in Supplementary Table S1. Reviews, non-original research papers and articles not based on Asian studies were excluded. In case of data overlap, the article with the largest sample size was chosen. Finally, papers that remain as preprints as of 15 November 2021 were also removed from the analysis (the corresponding authors were contacted by e-mail for confirmation). In addition, because COVID-19 does not seem to affect children and teens in the same way as adults ^11^, we analysed the data for pediatric patients separately. Initial screening was performed by the author ESF, followed by an ultimate extraction of data assessed independently by the authors ESF, DGG and MIV. When discrepancies were detected, MIV and ESF, experienced researchers, made the final decision whether or not to include the study.

Critical appraisal of selected studies was conducted following the assessment tool developed by Murad et al. ^26^, which was based on four domains: selection, ascertainment, causality, and reporting. The results from this tool provided a quality assessment of the studies for quantitative synthesis. Articles with unacceptable risk of bias (unclear or high risk of bias in ≥ 1 domain) were excluded from the analysis. Risk of bias was summarised in Supplementary Table S2.

For each parameter, in order to integrate all the reviewed information from studies with a low/acceptable risk of bias in one overall estimate that could be introduced in mathematical models, we performed different analyses. First of all, taking into account that in Asia the way to deal with the virus changed over time as they gained experience and that on February 12, 2020, WHO convened a Global Research and Innovation Forum on COVID-19 where 300 experts from 48 countries attended ^27^, we classified the studies considered in this review between those which cases were reported before February 12, 2020 (period A) and, given the impossibility of finding studies only with cases recorded after that date, those who include cases throughout the whole study period (period B). Subsequently, we performed a meta-regression for all the parameters with five or more studies considering the period as a categorical moderator, allowing the amount of residual heterogeneity to be different in each subset. Afterwards, we carried out the meta-analysis for all included studies and for the two subsets separately when they were confirmed to significantly affect the results, as following: for those parameters including mean and SD, or median and IQR, we conducted a meta-analysis of single means (when mean and SD were not reported, they were estimated from median and IQR ^28,29^), and when parameters were presented as percentages, a meta-analysis of proportions was performed. To account for the variability between and within studies, random-effects models were used. We considered model fitting by both the method of moments ^30^ and the Restricted Maximum Likelihood Method (REML)^31^. The natural logarithm transformation was applied to meet the normality assumption underlying the meta-analysis. The null hypothesis of no variance among studies (τ^2^=0) was tested using the Q-statistic and the degree of heterogeneity was quantified by the I^2^ index ^32^. Given the high heterogeneity among studies for most parameters considered, outliers and influencers diagnoses were also performed ^33^. Thus, we avoided biasing the conclusions from the meta-analysis by a few (potentially unusual) studies. Data were analysed using the statistical software R version 3.6.2 ^34^ and the “meta” ^35^, “metafor” ^36^ and “dmetar” ^37^ packages.

The PRISMA-ScR (Preferred Reporting Items for Systematic Reviews and Meta-analyses extension for Scoping Reviews) checklist was used to endorse good reporting in this article (see Supplementary PRISMA ScR Checklist) ^38^.

## Supporting information

PRISMA

Meta-analysis report

## Data Availability

All data produced in the present work are contained in the manuscript and the references cited in the text.

## Data availability

The data used to support the findings of this study are included in Supplementary table S1.

## Code availability

The meta-analysis has been conducted in R version 3.6.2 ^34^. Core packages for replicating the results are publicly available and include and include “meta” ^35^, “metafor” ^36^ and “dmetar” ^37^.

## Acknowledgements

We acknowledge Parques Nacionales (Ministerio para la Transición Ecológica y Reto Demográfico, Spain), and Generalitat Valenciana (Regional Government of Valencia, Spain) for the support of the Montgó-Dénia Research Station. We also thank John Y. Dobson and Elena Fonfría for their valuable comments during the editing process.

## Author Contributions

CB, ZH and MN: conceptualization. ESF and MIV: Study design, data analysis and results interpretation. ESF, MIV and DGG: literature review and data curation. ESF and MIV: writing-original draft preparation. CB: funding acquisition and project administration. All authors contributed to the manuscript writing-review and editing, and approved the submitted version.

