## Supplementary material for "Scoping review and meta-analysis of COVID-19 epidemiological parameters for modeling from early Asian studies": Table S1

**Supplementary Table S1.** Essential epidemiological parameters of COVID-19 for general population and children: common characteristics of selected studies

| | | | | 95% Credible interval | | | 95% Credible interval | | | | 95% Credible interval | | Interquartil Range | | | Age (years) | | | | | | |
|---|---|---|---|---|---|---|---|---|---|---|---|---|---|---|---|---|---|---|---|---|---|---|
| **Parameter** | **N** | **Mean** | **Range** | **Min** | **Max** | **SD** | **Min** | **Max** | **Units** | **Median** | **Min** | **Max** | **Min** | **Max** | **Geographic provenance of cases** | **Mean** | **Median** | **Other data** | **Male sex (%)** | **Other comments** | **Reference** | **DOI** |
| **General population** | | | | | | | | | | | | | | | | | | | | | | |
| **Serial interval (days)** | | | | | | | | | | | | | | | | | | | | | | |
| Serial interval | 468 transmision events | 3.96 | | 3.53 | 4.39 | 4.75 | 4.46 | 5.07 | Days | | | | | | Mainland China outside Hubei province | | | | | Data from Jan 21 to Feb 8, 2020 | Du Zhanwei et al. 2020 | https://doi.org/10.3201/eid2606.200357 |
| Serial interval | 48 pairs | 6.3 | | 5.2 | 7.6 | 4.2 | 3.1 | 5.3 | Days | | | | | | China | | | | | Dara from Jan 14 to Feb 12. | Bi et al. 2020 | https://doi.org/10.1016/S1473-3099(20)30287-5 |
| Serial interval | 35 secondary cases and 28 correponding primary cases | 5 | | 1.3 | 11.6 | 3.2 | | | Days | | | | | | Mainland China outside Hubei province | | | | | Gamma distribution Data from Jan 19 to Feb 17, 2020 | Zhang Juanjuan et al. 2020 | https://doi.org/10.1016/S1473-3099(20)30230-9 |
| Serial interval | 6 pairs | 7.5 | | 5.3 | 19 | 3.4 | | | Days | | | | | | Wuhan (China) | | | | | Data up to January 22, 2020 | Li et al. 2020 | https://doi.org/10.1056/NEJMoa2001316 |
| Serial interval | 28 pairs | 4.7 | | 3.7 | 6 | 2.9 | 1.9 | 4.9 | Days | 4 | 3.1 | 4.9 | | | Vietnam/Taiwan/China/Singapore/Germany/South Korea | | | | | Right truncation and lognormal distribution. | Nishiura et al. 2020 | https://doi.org/10.1016/j.ijid.2020.02.060 |
| Serial interval | 28 pairs | | | | | | | | Days | 3.9 | 3.1 | 4.8 | | | Vietnam/Taiwan/China/Singapore/Germany/South Korea | | | | | Without truncation. lognomal distribution | Nishiura et al. 2020 | https://doi.org/10.1016/j.ijid.2020.02.060 |
| Serial interval | 18 pairs | 4.8 | | 3.8 | 6.1 | 2.3 | 1.6 | 3.5 | Days | 4.6 | 3.5 | 5.9 | | | Vietnam/Germany/South Korea/China | | | | | weibull distribution | Nishiura et al. 2020 | https://doi.org/10.1016/j.ijid.2020.02.060 |
| Serial interval | 18 pairs | | | | | | | | Days | 4.1 | 3.2 | 5 | | | Vietnam/Germany/South Korea/China | | | | | Without truncation. lognomal distribution | Nishiura et al. 2020 | https://doi.org/10.1016/j.ijid.2020.02.060 |
| Serial interval | 28 | 6.6 | 3-15 | | | | | | Days | 4 | | | | | South Korea | | | | 53.6 | Data from Jan 20 to Feb 10 ,2020 | Ki et al. 2020 | https://doi.org/10.4178/epih.e2020007 |
| Serial interval observed | 21 transmission chain | 4.3 | 1-13 | | | | | | Days | 4 | | | 2 | 5 | Hong Kong | | | | | Data from Jan 16 to Feb 15, 2020 | Zhao Shi et al. 2020 | https://doi.org/10.3389/fphy.2020.00347 |
| Serial interval estimated | 21 transmission chain | 4.9 | | 3.6 | 6.2 | 4.4 | 2.9 | 8,3 | Days | | | | | | Hong Kong | | | | | Right- truncated lognormal distribution. Data from Jan 16 to Feb 15, 2020 | Zhao Shi et al. 2020 | https://doi.org/10.3389/fphy.2020.00347 |
| Serial interval estimated | 21 transmission chain | 3.9 | | 2.8 | 7.2 | 2.6 | 1.6 | 9,3 | Days | | | | | | Hong Kong | | | | | Lognormal distribution Data from Jan 16 to Feb 15, 2020 | Zhao Shi et al. 2020 | https://doi.org/10.3389/fphy.2020.00347 |
| Serial interval observed | 12 transmission chain | 3 | 1-8 | | | | | | Days | 2 | | | 2 | 4 | Hong Kong | | | | | Data from Jan 16 to Feb 15, 2020 | Zhao Shi et al. 2020 | https://doi.org/10.3389/fphy.2020.00347 |

| | | | | | | | | | | | | | | | | | | | | | | |
|---|---|---|---|---|---|---|---|---|---|---|---|---|---|---|---|---|---|---|---|---|---|---|
| Serial interval estimated | 12 transmissi on chain | 3.0 | | 2.1 | 3.9 | 2.0 | 1.2 | 4.6 | Days | | | | | | Hong Kong | | | | | Right- truncated lognormal distribution. Data from Jan 16 to Feb 15, 2020 | Zhao Shi et al. 2020 | https://doi.org/10.3389/fphy.2020.00347 |
| Serial interval estimated | 12 transmissi on chain | 3.0 | | 1.9 | 6.8 | 2.0 | 1.0 | 10.5 | Days | | | | | | Hong Kong | | | | | Lognormal distribution<br>Data from Jan 16 to Feb 15, 2020 | Zhao Shi et al. 2020 | https://doi.org/10.3389/fphy.2020.00347 |
| Serial interval | 77 pairs | 5.8 | | 4.8 | 6.8 | | | | Days | 5.2 | 4.1 | 6.4 | | | Guangzhou (China) | | | | | gamma distribution<br>Data from Dec 18 to March 5, 2020 | He Xi et al. 2020 | https://doi.org/10.1038/s41591-020-0869-5 |
| Serial interval | 57 | 4 | | 2.73 | 5.57 | | | | Days | | | | | | Singapore | | | | | Without intermediates.<br>Data from Jan 18 to Feb 17,2020 | Tindale et al. 2020 | https://doi.org/10.7554/eLife.57149 |
| Serial interval | 72 | 5 | | 3.82 | 6.12 | | | | Days | | | | | | Tianjin (China) | | | | | Without intermediates. Data from Jan 13 to Feb 20, 2020 | Tindale et al. 2020 | https://doi.org/10.7554/eLife.57149 |
| Serial interval | 57 | 4.17 | | 2.44 | 5.89 | | | | Days | | | | | | Singapore | | | | | With intermediates.<br>Data from Jan 18 to Feb 17,2020 | Tindale et al. 2020 | https://doi.org/10.7554/eLife.57149 |
| Serial interval | 72 | 4.31 | | 2.91 | 5.72 | | | | Days | | | | | | Tianjin (China) | | | | | With intermediates.<br>Data from Jan 13 to Feb 20, 2020 | Tindale et al. 2020 | https://doi.org/10.7554/eLife.57149 |
| Serial interval | 90 | 6.6 | | 0.7 | 19 | | | | Days | | | | | | Lombardy (Italy) | | | | | gamma distribution | Cereda et al. 2020 | https://arxiv.org/abs/2003.09320 |
| Serial interval ** | 7 | 1.57 | | | | 1.5 1 | | | Days | | | | | | Anhui (China) | | 22 | | 71.4 | Data from Jan 25 – Feb 20, 2020 | Huang Lei et al. 2020 | https://doi.org/10.1016/j.jinf.2020.03.006 |
| Serial interval | 28 | 5.5 | | 4.1 | 7 | | | | Days | | | | | | Shenzhen (China) | | | | | Cases from Jan 1 Feb 28, 2020 | Wen et al. 2020 | https://doi.org/10.1101/2020.03.22.20035246 |
| Serial interval | 12 | 6.5 | | 2.45 | 17.38 | | | | Days | 1.91 | 0.37 | 6.16 | | | Chongqing (China) | | | 10-35 | 52.2 | Data from Jan 25 to Feb 18, 2020.<br>Outcomes until Feb 23, 2020 | Liao et al. 2020 | https://doi.org/10.1016/j.xinn.2020.04.001 |
| **Incubation period (days)** | | | | | | | | | | | | | | | | | | | | | | |
| Incubation period | 88 | 6.4 | | 5.6 | 7.7 | | | | Days | | | | | | Travelers infected in Wuhan (63 residents and visitors) | | | Range 2-72 | | Weibull distribution | Backer et al. 2020 | https://doi.org/10.2807/1560-7917.ES.2020.25.5.2000062 |
| Incubation period | 88 | 6.5 | | 5.6 | 7.9 | | | | Days | | | | | | Travelers infected in Wuhan (63 residents and visitors) | | | Range 2-72 | | Gamma distribution | Backer et al. 2020 | https://doi.org/10.2807/1560-7917.ES.2020.25.5.2000062 |
| Incubation period | 88 | 6.8 | | 5.7 | 8.8 | | | | Days | | | | | | Travelers infected in Wuhan (63 residents and visitors) | | | Range 2-72 | | Lognormal distribution | Backer et al. 2020 | https://doi.org/10.2807/1560-7917.ES.2020.25.5.2000062 |
| Incubation period | 10 | 5.2 | | 4.1 | 7 | | | | Days | | | | | | Wuhan (China) | | 59 | | | N=245 but the estimation was performed with 10 confirmed cases.<br><br>Confirmed cases reported by Jan 22, 2020 | Li et al. 2020 | https://doi.org/10.1056/NEJMoa2001316 |
| Incubation period | 181 | 5.5 | | | | 2.2 | | | Days | 5.1 | 4.5 | 5.8 | | | 50 provinces, regions and countries outside Wuhan. Hubei province | | 44.5 | | 59.7 | 2.2% unknown sex | Lauer et al. 2020 | https://doi.org/10.7326/M20-0504 |

| | | | | | | | | | | | | | | | | | | | | | | |
|---|---|---|---|---|---|---|---|---|---|---|---|---|---|---|---|---|---|---|---|---|---|---|
| Incubation period | 108 | | | | | | | | Days | 5.5 | 4.4 | 7 | | | Only cases detected outside mainland China | | | | | | Lauer et al. 2020 | https://doi.org/10.7326/M20-0504 |
| Incubation period | 73 | | | | | | | | Days | 4.8 | 4.2 | 5.6 | | | Cases inside mainland China | | | | | Data from Jan 4 to February 24, 2020 | Lauer et al. 2020 | https://doi.org/10.7326/M20-0504 |
| Incubation period | 52 | 5.4 | | 4.3 | 6.6 | | | | Days | 4.7 | 3.6 | 5.8 | | | Residents from other locations who travelled to Wuhan | | | | | Accounting without right truncation. Weibull | Linton et al. 2020 | https://doi.org/10.3390/jcm9020538 |
| Incubation period | 52 | 5.3 | | 4.3 | 6.6 | | | | Days | 4.7 | 3.8 | 5.7 | | | Residents from other locations who travelled to Wuhan | | | | | Accounting without right truncation. Gamma | Linton et al. 2020 | https://doi.org/10.3390/jcm9020538 |
| Incubation period | 52 | 5 | | 4.2 | 6 | | | | Days | 4.3 | 3.5 | 5.1 | | | Residents from other locations who travelled to Wuhan | | | | | Accounting without right truncation. Lognormal | Linton et al. 2020 | https://doi.org/10.3390/jcm9020538 |
| Incubation period | | 5.6 | | 4.4 | 7.4 | | | | Days | 4.6 | 3.7 | 5.7 | | | Residents from other locations who travelled to Wuhan | | | | | Right-truncated incubation. Lognormal | Linton et al. 2020 | https://doi.org/10.3390/jcm9020538 |
| Incubation period | 158 | 5.8 | | 5.2 | 6.5 | | | | Days | 5.3 | 4.7 | 6 | | | Residents from other locations who travelled to Wuhan + Wuhan residents | | | Range 30-59 | 58 | Accounting without right truncation. Weibull | Linton et al. 2020 | https://doi.org/10.3390/jcm9020538 |
| Incubation period | 158 | 6 | | 5.3 | 6.7 | | | | Days | 5.6 | 4.9 | 6.4 | | | Residents from other locations who travelled to Wuhan + Wuhan residents | | | Range 30-59 | 58 | Accounting without right truncation. Gamma | Linton et al. 2020 | https://doi.org/10.3390/jcm9020538 |
| Incubation period | 158 | 5.6 | | 5 | 6.3 | | | | Days | 5 | 4.4 | 5.6 | | | Residents from other locations who travelled to Wuhan + Wuhan residents | | | Range 30-59 | 58 | Accounting without right truncation. Lognormal | Linton et al. 2020 | https://doi.org/10.3390/jcm9020538 |
| Incubation period | 1099 | | | | | | | | Days | 4 | | | 2 | 7 | Mainland China | | 47 | | 58.1 | Data up to January 29 | Guan et al. 2020 | https://doi.org/10.1056/NEJMoa2002032 |
| Incubation period | 183 | 5.95 | | | | 4.3 | | | Days | 4.8 | 4.2 | 5.4 | | | China | | | | | Dara from Jan 14 to Feb 12. | Bi et al. 2020 | https://doir.org/10.1016/S1473-3099(20)30287-5 |
| Incubation period | 49 | 5.2 | | 1.8 | 12.4 | | | | Days | | | | | | Mainland China outside Hubei province | | | | | N=8759. but mean incubation period was calculated with 49 cases. Lognormal Data from Jan 19 to Feb 17, 2020 | Zhang Juanjuan et al. 2020 | https://doi.org/10.1016/S1473-3099(20)30230-9 |
| Incubation period | 50 | 4.9 | | 4.4 | 5.5 | | | | Days | | | | | | Global | | | | | | Jiang et al. 2020 | https://doi.org/10.1002/jmv.25708 |
| Incubation period | 25 | 3.9 | 0-15 | | | | | | Days | 3 | | | | | South Korea | | 42 | | 53.6 | Data from Jan 20 to Feb 10, 2020 | Ki et al. 2020 | https://doi.org/10.4178/epih.e2020007 |
| Incubation period | 16 | 4.8 | | | | 2.6 | | | Days | | | | | | Wuhan (China) | | | | | Data up to February 7 | Liu Tao et al. 2020 | https://doi.org/10.1101/2020.01.25.919787 |
| Incubation period | 483 | 7.4 | | | | | | | Days | 7 | | | | | Henan province (China) | | | | | Data from Jan 21 to Feb 14, 2020 | Wang et al. 2020 | https://doi.org/10.1016/j.ijid.2020.04.051 |
| Incubation period | 24 | 4.2 | | 3.5 | 5.1 | | | | Days | | | | | | China/Mexico/ South Korea/Thailand | | | | | | Sanche et al. 2020 | https://doi.org/10.3201/eid2607.200282 |

| | | | | | | | | | | | | | | | | | | | | | | |
|---|---|---|---|---|---|---|---|---|---|---|---|---|---|---|---|---|---|---|---|---|---|---|
| Incubation period | 19 | | | | | | | | Days | 4 | | | 3 | 6 | Singapore | | | | | Data up to February 15 | Pung et al. 2020 | https://doi.org/10.1016/S0140-6736(20)30528-6 |
| Incubation period | 91 | | | | | | | | Days | 6 | | | 3 | 8 | Zhejiang (China) | | 50 | | 40.66 | Data from Jan 20 to Feb 11, 2020 | Quian et al. 2020 | https://doi.org/10.1093/qjmed/hcaa089 |
| Incubation period (imported) | 15 | | | | | | | | Days | 8 | | | 4 | 10 | Changzhou city. Jiangsu province (China) | | 35 | | 66.7 | Data from Jan 23 to February 27,2020 | Xu Tianmin et al. 2020 | https://doi.org/10.1016/j.ijid.2020.03.022 |
| Incubation period (secondary) | 17 | | | | | | | | Days | 8 | | | 4 | 11 | Changzhou city. Jiangsu province (China) | | 37 | | 41.2 | Data from Jan 23 to February 27,2020 | Xu Tianmin et al. 2020 | https://doi.org/10.1016/j.ijid.2020.03.022 |
| Incubation period (tertiary) | 19 | | | | | | | | Days | 12 | | | 9 | 14 | Changzhou city. Jiangsu province (China) | | 53 | | 41.2 | Data from Jan 23 to February 27,2020 | Xu Tianmin et al. 2020 | https://doi.org/10.1016/j.ijid.2020.03.022 |
| Incubation period | 6 | | | | | | | | Days | 7.5 | | | 1 | 16 | China | | | | | Data from Jan 8 to Feb26, 2020 | Shen et al. 2020 | https://doi.org/10.1002/ppul.24762 |
| Incubation period | 25 | | 2-12 | | | | | | Days | 4 | | | | | China | | 44 | | 48 | Data from Jan 31 to Feb 16, 2020 | Han et al. 2020 | https://doi.org/10.1002/jmv.25835 |
| Incubation period | 11 clusters | 6.28 | 1-14 | | | | | | Days | | | | | | Qingdao (China) | | | | | Data from Jan 29 to Feb 26,2020 | Jia et al. 2020 | https://doi.org/10.1017/dmp.2020.59 |
| Incubation period | 32 | | | | | | | | Days | 5 | | | 2 | 9 | Zhejiang (China) | 41.15 | | <60 | | Data from Jan 17 to Feb 12, 2020 | Lian et al. 2020 | https://doi.org/10.1093/cid/ciaa242 |
| Incubation period | 23 | | | | | | | | Days | 4 | | | 3 | 7 | Beijing (China) | | | | | Patients admitted from Jan 21 to February 8, 2020 | Zhao Wen et al. 2020 | https://doi.org/10.1101/2020.03.13.20035436 https://doi.org/10.15212/CVIA.2021.0019 |
| Incubation period | 56 | | | | | | | | Days | 4 | | | 3 | 5 | Zhejiang (China) | | 41 | | 56 | Data from Jan 10 to Jan 26, 020 | Xu XIao-Wei et al. 2020 | http://dx.doi.org/10.1136/bmj.m606 |
| Incubation period | 93 | 5.99 | | 4.97 | 7.14 | | | | Days | | | | | | Singapore | | | | | Without intermediaries. All. Data from Jan 18 to Feb 28,2020 | Tindale et al. 2020 | https://doi.org/10.7554/eLife.57149 |
| Incubation period | 93 | 5.91 | | 4.50 | 7.64 | | | | Days | | | | | | Singapore | | | | | Without intermediaries. Early, OS on or prior Jan 31, 2020 | Tindale et al. 2020 | https://doi.org/10.7554/eLife.57149 |
| Incubation period | 93 | 6.06 | | 4.7 | 7.67 | | | | Days | | | | | | Singapore | | | | | Without intermediaries. Late, OS after Jan 31, 2020 | Tindale et al. 2020 | https://doi.org/10.7554/eLife.57149 |
| Incubation period | 93 | 1.71 | | | | 3.0 1 | | | Days | | | | | | Singapore | | | | | From end of possible viral exposure window to onset. Data from Jan 23 to Feb 26,2020 | Tindale et al. 2020 | https://doi.org/10.7554/eLife.57149 |
| Incubation period | 135 | 8.68 | | 7.7 | 9.7 | | | | Days | | | | | | Tianjin (China) | | | | | Without intermediaries. All Data from Jan 2 to Feb 22,2020 | Tindale et al. 2020 | https://doi.org/10.7554/eLife.57149 |
| Incubation period | 135 | 6.88 | | 5.97 | 7.87 | | | | Days | | | | | | Tianjin (China) | | | | | Without intermediaries Early OS on or prior Jan 31, 2020 | Tindale et al. 2020 | https://doi.org/10.7554/eLife.57149 |
| Incubation period | 135 | 12.4 | | 11.1 | 13.7 | | | | Days | | | | | | Tianjin (China) | | | | | Without intermediaries Late, OS after Jan 31, 2020 | Tindale et al. 2020 | https://doi.org/10.7554/eLife.57149 |
| Incubation period | 135 | 4.98 | | | | 4.8 3 | | | Days | | | | | | Tianjin (China) | | | | | From end of possible viral exposure window to onset. Data from Jan 2 to Feb 22,2020 | Tindale et al. 2020 | https://doi.org/10.7554/eLife.57149 |
| Incubation period | 59 | | | | | | | | Days | 4 | | | 2 | 7 | Jilin (China) | | 41 | | 57.6 | Data from Jan 21 to March 5, 2020 | Tian Suyan et al .2020 | https://doi.org/10.1007/s12250-020-00317-z |

| | | | | | | | | | | | | | | | | | | | | | | |
|---|---|---|---|---|---|---|---|---|---|---|---|---|---|---|---|---|---|---|---|---|---|---|
| Incubation period | 25 | 3.9 | 0-15 | | | | | | Days | 3 | | | | | Republic of Korea | | | | | Data from Jan 20 to Feb 10, 2020 | Ki et al. 2020 | https://doi.org/10.4178/epih.e2020007 |
| Incubation period | 197 | 6.14 | 0-55 | | | 9.27 | | | Days | | | | | | Hubei (China) | | | | | Data from Jan 17 to Feb 26, 2020 | Zhou Fating et al. 2020 | https://doi.org/10.1101/2020.03.26.20041426 |
| Incubation period * | 55 | | | | | | | | Days | 7 | | | 3.5 | 10 | Jiangsu (China) | | 45 | | 49.1 | Data from Jan 23 to April 6 | Jiang et al. 2020 | https://doi.org/10.1101/2020.04.10.20060335 |
| Incubation period | 10 | 7 | | | | 2.59 | | | Days | | | | | | China | 56.5 | | | 50 | Data from Dec 18, 2019 to Jan 28,2020 | Xia et al. 202 | https://doi.org/10.1016/j.jcv.2020.104360 |
| Incubation period * | 262 | 6.7 | | | | 5.2 | | | Days | | | | | | Beijing (China) | | 47.5 | | 48.5 | Data up to February 10 | Tian Suyan et al .2020 | https://doi.org/10.1016/j.jinf.2020.02.018 |
| Incubation period ** | 6 | 2.17 | | | | 1.17 | | | Days | | | | | | Anhui (China) | | 22 | | 83.3 | Late january 2020 | Huang et al. 2020 | https://doi.org/10.1016/j.jinf.2020.03.006 |
| Incubation period | 59 | 5.84 | | | | 2.93 | | | Days | | | | | | 10 regions of China except Hubei province (China) | 41.9 | | | 57.6 | Data from Dec 29,2019 to Feb 5, 2020 | Men et al. 2020 | https://doi.org/10.1101/2020.02.24.20027474 |
| Incubation period | 92 | | | | | | | | Days | 5 | | | 3.1 | 8.2 | Shenzhen (China) | | | | | Cases from Jan 1 to Feb 28, 2020 | Wen et al. 2020 | https://doi.org/10.1101/2020.03.22.20035246 |
| Exposure to system onset | 62 | | | | | | | | Days | 7 | | | 3.5 | 9 | Two provinces outside Hubei (China) | 43.8 | | | 51.6 | Data from Jan 12 to Feb 13, 2020 | Miao et al. 2020 | https://doi.org/10.1101/2020.03.22.20040782 |
| Incubation period | 55 | 8.42 | | 6.55 | 10.29 | | | | Days | | | | | | Beijing (China) | | 44 | | 60.0 | Patients admitted from Dec 27, 2020 to Feb 18, 2020 | Yang et al. 2020 | https://doi.org/10.1101/2020.02.28.20028068 |
| Incubation period | 11 | 7.17 | | 5..15 | 10.1 | | | | Days | 6.58 | 4.43 | 9.6 | | | Chongqing (China) | | | 10-35 | 52.2 | Data from Jan 25 to Feb 18, 2020. Outcomes until Feb 23, 2020 | Liao et al. 2020 | https://doi.org/10.1016/j.xinn.2020.04.001 |
| **Onset of symptoms/illness onset to first medical visit (days)** | | | | | | | | | | | | | | | | | | | | | | |
| Illness onset to first medical visit | 45 | 5.8 | | 4.3 | 7.5 | | | | Days | | | | | | Wuhan (China) | | | 26-82 | | Weibull distribution. Before January 1 | Li et al. 2020 | https://doi.org/10.1056/NEJMoa2001316 |
| Illness onset to first medical visit | 207 | 4.6 | | 4.1 | 5.1 | | | | Days | | | | | | Wuhan (China) | | | 21-89 | | Weibull distribution. Between January 1 and January 11 | Li et al. 2020 | https://doi.org/10.1056/NEJMoa2001316 |
| Onset of symptom to first medical visit | 168 | | | | | | | | Days | 1 | | | 0 | 4 | Hainan (China) | | 51 | | | Data from Jan 22 to March 13, 2020 (final date of follow-up) | Yan et al. 2020 | https://doi.org/10.1101/2020.03.19.20038539 |
| Illness onset to visit hospital | 262 | 4.5 | | | | 3.7 | | | Days | | | | | | Beijing (China) | | 47,5 | | | Data from Jan 20 to Feb 10, 2020 | Tian Sijia et al. 2020 | https://doi.org/10.1016/j.jinf.2020.02.018 |
| Illness onset to consultation | 788 | | | | | | | | Days | 2 | | | 1 | 4 | Zhejiang province (China) | 45.8 | | | 51.6 | All ages together. Data from Jan 17 to Feb 12, 2020. Clinical outcomes followed up to Feb 12. | Lian et al. 2020 | https://doi.org/10.1093/cid/ciaa242 |
| Illness onset to first medical visit | 417 | | | | | | | | Days | 1 | | | 0 | 3 | Shenzhen (China) | 45.4 | | | 47.2 | Cases from Jan 1 Feb 28, 2020 | Wen et al. 2020 | https://doi.org/10.1101/2020.03.22.20035246 |
| Symptoms onset to first medical visit | 62 | | | | | | | | Days | 3 | | | 1 | 6 | Two provinces outside Hubei (China) | 43.8 | | | 51.6 | Data from Jan 12 to Feb 13, 2020 | Miao et al. 2020 | https://doi.org/10.1101/2020.03.22.20040782 |
| Symptoms onset to first medical visit | 42 | | | | | | | | Days | 1.4 | 0.8 | 2.4 | | | Chongqing (China) | | | 10-35 | 52.2 | Data from Jan 25 to Feb 18, 2020. Outcomes until Feb 23, 2020 | Liao et al. 2020 | https://doi.org/10.1016/j.xinn.2020.04.001 |
| Onset to first medical visit | 57 | 5 | 2-10 | | | 1.9 | | | Days | | | | | | China | 46.5 | | | 54.4 | Data from Jan 21 to Feb 15, 2020 | Qi Shi et al. 2020 | https://doi.org/10.1101/2020.04.02.20042614 |

| | | | | | | | | | | | | | | | | | | | | | | |
|---|---|---|---|---|---|---|---|---|---|---|---|---|---|---|---|---|---|---|---|---|---|---|
| **From first visit to diagnosis/admission** | | | | | | | | | | | | | | | | | | | | | | |
| From visit hospital to defined confirmed case | 262 | 2.1 | | | | 1.9 | | | Days | | | | | | Beijing (China) | | 47.5 | | 48.5 | Data from Jan 20 to Feb 10, 2020 | Tian Sijia et al. 2020 | https://doi.org/10.1016/j.jinf.2020.02.018 |
| From visit to confirmed diagnosis | 91 | | | | | | | | Days | 1 | | | 1 | 2 | Zhejiang province (China) | | 50 | | 40.7 | Data colected from Jan 20 to Feb 11, 2020. Final folow-up until Feb 16, 2020 | Qian et al. 2020 | https://doi.org/10.1093/qjmed/hcaa089 |
| Medical visit to hospital admission | 417 | | | | | | | | Days | 0 | | | 0 | 2 | Shenzhen (China) | 45.4 | | | 47.2 | Cases from Jan 1 Feb 28, 2020 | Wen et al. 2020 | https://doi.org/10.1101/2020.03.22.20035246 |
| **Onset of symptoms/illness onset to diagnosis (days)** | | | | | | | | | | | | | | | | | | | | | | |
| Onset of symptom to diagnosis | 73 | | | | | | | | Days | 11 | | | 8 | 14 | Wuhan (China) | | 67 | | 61.6 | Data from December 24, 2019 to February 7,2020 | Tang et al. 2020 | https://doi.org/10.1016/j.chest.2020.03.032 |
| Onset of symptom to diagnosis * | 267 | | | | | | | | Days | 4 | | | 2 | 8 | Chongqing (China) | | 48 | | 55.8 | Data from Jan 19 to Feb 16, 2020 | Qi Di et al. 2020 | https://doi.org/10.1101/2020.03.01.20029397 |
| Onset of symptom to diagnosis * | 51 | | 1-16 | | | | | | Days | 6 | | | 3 | 8 | Chongqing (China) | | 45 | | 37.3 | Data from Jan 20 to Feb 11, 2020 | Liu Lei et al. 2020 | https://doi.org/10.1101/2020.02.20.20025536 |
| Onset of symptom to diagnosis | 135 | 5.2 | | | | 4.2 | | | Days | | | | | | Tianjin (China) | | | | | Data from Jan 2 to Feb 22, 2020 | Tindale et al. 2020 | https://doi.org/10.7554/eLife.57149 |
| Onset of symptom to diagnosis | 93 | 7.43 | | | | 5.2 8 | | | Days | | | | | | Singapore | | | | | Data from Jan 23 to Feb 26, 2020 | Tindale et al. 2020 | https://doi.org/10.7554/eLife.57149 |
| Onset of symptom to diagnosis | 652 | | | | | | | | Days | 4 | | | 2 | 7 | Zhejiang (China) | 41.15 | | <60 | 53.53 | Data from Jan 17 to Feb 12, 2020 | Lian et al. 2020 | https://doi.org/10.1093/cid/ciaa242 |
| Onset of symptom to diagnosis | 136 | | | | | | | | Days | 4 | | | 2 | 7 | Zhejiang (China) | 68.27 | | >60 | 42.65 | Data from Jan 17 to Feb 12, 2020 | Lian et al. 2020 | https://doi.org/10.1093/cid/ciaa242 |
| Onset of symptom to diagnosis | 168 | | | | | | | | Days | 4.5 | | | 2 | 7 | Hainan (China) | | 51 | | 48.2 | Data from Jan 22 to March 13,2020 | Yan et al. 2020 | https://doi.org/10.1101/2020.03.19.20038539 |
| Onset of symptom to diagnosis | 66 | | 0-20 | | | | | | Days | 4 | | | | | Republic of Korea | | | | | Data up to March 12, 2020 | Choe et al 2020 | https://doi.org/10.24171/j.phrp.2020.11.2.05 |
| Onset of symptom to diagnosis | 9 | | | | | | | | Days | 5 | | | 1 | 11 | Macau | | 54 | | 30 | Data from Jan 21 to Feb 16 | Lo et al. 2020 | https://doi.org/10.7150/ijbs.45357 |
| Onset of symptom to diagnosis | 82 | | | | | | | | Days | 7 | | | 4 | 10 | Wuhan (China) | | 72.5 | | 65.9 | Data from Jan 11 to Feb 10, 2020 | Zhang Bicheng et al. 2020 | https://doi.org/10.1371/journal.pone.0235458 |
| Onset of symptom to diagnosis * | 102 | | | | | | | | Days | 8 | | | 5 | 14 | Wuhan (China) | | 54 | | 52 | Data from Jan 3 to Feb 1, 2020 | Cao et al. 2020 | https://doi.org/10.1093/cid/ciaa243 |
| Onset of symptom to diagnosis | 25 | 7.44 | | | | 3.1 | | | Days | | | | | | Anhui (China) | 49.4 | | | 64 | Data from Jan 20 to Feb 9, 2020. Outcomes follow-up to Feb 18, 2020 | Wang et al. 2020 | https://doi.org/10.1016/j.ijid.2020.03.070 |
| Onset of symptom to diagnosis | 100 | | | | | | | | Days | 5 | | | 3 | 9 | Anhui (China) | 39.47 | | | 55 | Data from Jan 20 to Feb 9, 2020 Outcomes follow-up to Feb 18, 2020 | Wang et al. 2020 | https://doi.org/10.1016/j.ijid.2020.03.070 |
| Onset of symptom to diagnosis | 133 | | | | | | | | Days | 8 | | | 5 | 11 | Wuhan (China) | | 70 | | 55.6 | Patients enrolled from Jan 25 to Feb 25, 2020 | Wang Yang et al. 2020 | https://doi.org/10.1164/rccm.202003-0736LE |
| Illness onset to diagnosis | 25 | 5.2 | 0-16 | | | | | | Days | | | | | | South Korea | | | | 53.6 | Data from Jan 20 to Feb 15, 2020 | Ki et al. 2020 | https://doi.org/10.4178/epih.e2020007 |
| Illness onset to diagnosis | 4 | 5 | | | | 4.5 | | | Days | | | | | | Shangai (China) | 44.25 | | | 75 | Data from Jan 21 to Feb 4, 2020 | Wang Zhenwei et al. 2020 | https://doi.org/10.5582/bst.2020.01030 |
| Onset to PCR confirmation (symptom-based surveillance) | 292 | 5.46 | | 4.99 | 5.92 | 3.7 3 | | | Days | | | | | | China | | | | | Gamma distribution Data from Jan 14 to Feb 9, 2020 | Bi et al. 2020 | https://doi.org/10.1016/S1473-3099(20)30287-5 |

| | | | | | | | | | | | | | | | | | | | | | | |
|---|---|---|---|---|---|---|---|---|---|---|---|---|---|---|---|---|---|---|---|---|---|---|
| Onset to PCR confirmation (contact-based surveillance) | 87 | 3.18 | | 2.65 | 3.72 | 1.79 | | | Days | | | | | | China | | | | | Gamma distribution Data from Jan 14 to Feb 9, 2020 | Bi et al. 2020 | https://doi.org/10.1016/S1473-3099(20)30287-5 |
| **Onset of symptoms/illness onset to hospital admission (days)** | | | | | | | | | | | | | | | | | | | | | | |
| Onset of symptoms to hospital admission | 1310 | 4.4 | | 0 | 14 | | | | Days | | | | | | Mainland China outside Hubei province | | | | | Data from Dec 24, 2019 to Jan 27, 2020 | Zhang Juanjuan et al. 2020 | https://doi.org/10.1016/S1473-3099(20)30230-9 |
| Onset of symptoms to hospital admission | 691 | 2.6 | | 0 | 9 | | | | Days | | | | | | Mainland China outside Hubei province | | | | | Data from Jan 28 - Feb 17, 2020 | Zhang Juanjuan et al. 2020 | https://doi.org/10.1016/S1473-3099(20)30230-9 |
| Onset of symptoms to hospital admission | 41 | | | | | | | | Days | 7 | | | 4 | 8 | Patients from Wuhan Hospital (China) | | 49 | | 73 | Data from Dec 1, 2019 to Jan 2,2020 | Huang Chaolin et al. 2020 | https://doi.org/10.1016/S0140-6736(20)30183-5 |
| Onset of symptoms to hospital admission | 138 | | | | | | | | Days | 7 | | | 4 | 8 | Wuhan (China) | | 56 | | 54.3 | Data from Jan 1 to Jan 28, 2020 | Wang Dawei et al. 2020 | https://doi.org/10.1001/jama.2020.1585 |
| Onset of symptoms to hospital admission | 181 | | | | | | | | Days | 1.2 | | | | | 24 countries and regions outside mainland china and 25 provinces mainland | | 44.5 | | 59.7 | 2.2% unknown sex Data from Jan 4 to Feb 2, 2020 | Lauer et al. 2020 | https://doi.org/10.7326/M20-0504 |
| Onset of symptoms to hospital admission | 1012 | | | | | | | | Days | 10 | | | 7 | 14 | Wuhan (China) | | 50 | | 51.8 | Patients admitted between Feb 7 and Feb 12, 2020 Clinical course recorded up to Feb 22, 2020 | Wang X et al. 2020 | https://doi.org/10.1016/j.cmi.2020.03.032 |
| Onset of symptoms to hospital admission * | 102 | | | | | | | | Days | 6 | | | 3 | 7 | Wuhan (China) | | 54 | | 52 | Data from Jan 3 to Feb 1, 2020 | Cao et al. 2020 | https://doi.org/10.1093/cid/ciaa243 |
| Onset of symptoms to hospital admission | 24 | 7 | | | | 4 | | | Days | | | | | | Seattle region (USA) | 64 | | | 63 | | Bhatraju et al. 2020 | https://doi.org/10.1056/NEJMoa2004500 |
| Onset of symptoms to hospital admission * | 79 | | | | | | | | Days | 12 | | | 10 | 15 | Wuhan (China) | | 60 | | 55.7 | Data from Feb 2 to Feb 23, 2020 | Xie et al. 2020 | https://doi.org/10.1111/liv.14449 |
| Onset of symptoms to hospital admission * | 221 | | | | | | | | Days | 7 | | | 4 | 10 | Wuhan (China) | | 55 | | 48.9 | Data from Jan 2 to Feb 10, 2020 | Zhang Guqin et al. 2020 | https://doi.org/10.1016/j.jcv.2020.104364 |
| Onset of symptoms to hospital admission * | 267 | | | | | | | | Days | 7 | | | 3 | 10 | Chongqing (China) | | 48 | | 55.8 | Data from Jan 19 to Feb 16 2020 | Qi Di et al. 2020 | https://doi.org/10.1101/2020.03.01.20029397 |
| Onset of symptoms to hospital admission | 67 | | | | | | | | Days | 6 | | | 4 | 9 | Wuhan (China) | | 42 | | 46 | Patients hospitalized from Jan 16 to Jan 29, 2020 | Wang Zhongliang et al. 2020 | https://doi.org/10.1093/cid/ciaa272 |
| Onset of symptoms to hospital admission | 82 | | | | | | | | Days | 10 | | | 7 | 15 | Wuhan (China) | | 72.5 | | 65.9 | Data from Jan 11 to Feb 10, 2020 | Zhang Bicheng et al. 2020 | https://doi.org/10.1371/journal.pone.0235458 |
| Onset of symptoms to hospital admission | 168 | | | | | | | | Days | 5 | | | 2 | 7 | Hainan (China) | | 51 | | 48.2 | Data from Jan 22 to March 13, 2020 | Yan et al. 2020 | https://doi.org/10.1101/2020.03.19.20038539 |
| Onset of symptoms to hospital admission | 21 | 3.5 | | | | | | | Days | | | | | | Whashington State (USA) | | | | | | Arentz et al. 2020 | https://doi.org/10.1001/jama.2020.4326 |
| Onset of symptoms to hospital admission | 17 | | | | | | | | Days | 4 | | | 3 | 9 | Singapore | | 40 | | 41 | Data up to February 15, 2020 | Pung et al. 2020 | https://doi.org/10.1016/S0140-6736(20)30528-6 |
| Onset of symptoms to hospital admission | 652 | | | | | | | | Days | 3 | | | 1 | 7 | Zhejiang (China) | 41.15 | | <60 | 53.53 | Data from Jan 17 to Feb 12, 2020 | Lian et al. 2020 | https://doi.org/10.1093/cid/ciaa242 |
| Onset of symptoms to hospital admission | 136 | | | | | | | | Days | 3 | | | 1 | 6 | Zhejiang (China) | 68.27 | | >60 | 42.65 | Data from Jan 17 to Feb 12, 2020 | Lian et al. 2020 | https://doi.org/10.1093/cid/ciaa242 |

| | | | | | | | | | | | | | | | | | | | | | | |
|---|---|---|---|---|---|---|---|---|---|---|---|---|---|---|---|---|---|---|---|---|---|---|
| Onset of symptoms to hospital admission | 8 | | | | | | | | Days | 7 | | | 4 | 10 | Hong Kong | | 64.5 | | 50 | Patients admitted from Jan 22 to Feb 11, 2020 | Ling lowell et al. 2020 | https://www.ncbi.nlm.nih.gov/pubmed/32248675 |
| Onset of symptoms to hospital admission | 66 | | | | | | | | Days | 4.5 | | | | | Republic of Korea | | | | | Up to March 12, 2020 | Choe et al 2020 | https://doi.org/10.24171/j.phrp.2020.11.2.05 |
| Onset of symptoms to hospital admission | 62 | | | | | | | | Days | 2 | | | 1 | 4.3 | Zhejiang (China) | | 41 | | 56 | Data from Jan 10 to Jan 26, 2020 | Xu Xiao-Wei et al. 2020 | http://dx.doi.org/10.1136/bmj.m606 |
| Onset of symptoms to hospital admission | 48 | 8.5 | 1-12 | | | 3.9 | | | Days | | | | | | Hong Kong | | | | | Data from Jan 26 to Feb 28, 2020 | Leung et al. 2020 | https://doi.org/10.3201/eid2701.201543 |
| Onset of symptoms to hospital admission | 24 | 5.5 | | | | 7 | | | Days | | | | | | Liaocheng city (China) | 48 | | | 37.5 | Period unknown, but at least one admission after Jan 9, 2020 | Tian Suochen et al .2020 | https://doi.org/10.1186/s12889-021-10713-z |
| Onset of symptoms to hospital admission | 75 | | | | | | | | Days | 4 | | | 2 | 7.5 | Zhejiang (China) | 46.37 | | | 55 | Data from January to Feb 20, 2020 | Hong et al. 2020 | http://dx.doi.org/10.21037/atm.2020.03.147 |
| Onset of symptoms to hospital admission | 125 | | | | | | | | Days | 4 | | | | | Anhui (China) | 38.78 | | | 55 | Data from Jan 20 to Feb 9,2020 | Wang et al. 2020 | https://doi.org/10.1016/j.ijid.2020.03.070 |
| Onset of symptoms to hospital admission | 134 | | | | | | | | Days | 10 | | | 8 | 13 | Wuhan (China) | | | | | Cases from Dec 30, 2019 to Feb 20, 2020 | Zhang Lin et al. 2020 | https://doi.org/10.1017/s0950268820002010 |
| Symptom onset to hospital admission | 18 | | 2-15 | | | | | | Days | 7 | | | | | Zhengzou (China) | | 39 | | 55.6 | Data from Jan 21 to Feb 5. Final follow-up was Feb 7, 2020 | WangL et al. 2020 | https://doi.org/10.1183/13993003.00398-2020 |
| Symptom onset to hospitalization * | 339 | | | | | | | | Days | 10 | | | 7 | 14 | Wuhan (China) | | | | | Data up to Feb 28, 2020 | Wang et al. 2020 | https://doi.org/10.1016/j.jinf.2020.03.019 |
| Symptom onset to hospitalization | 274 | | | | | | | | Days | 10 | | | 7 | 12 | Wuhan (China) | | 62 | | 62.3 | Data from Jan 13 to Feb 12, 2020 | Chen et al. 2020 | http://dx.doi.org/10.1136/bmj.m1091 |
| Illness onset to hospital admission | 44 | 12.5 | | 10.3 | 14.8 | | | | Days | | | | | | Wuhan (China) | | | | | Before January 1, 2020 | Li et al. 2020 | https://doi.org/10.1056/NEJMoa2001316 |
| Illness onset to hospital admission | 189 | 9.1 | | 8.6 | 9.7 | | | | Days | | | | | | Wuhan (China) | | | | | Data from Jan 1 to Jan 11 | Li et al. 2020 | https://doi.org/10.1056/NEJMoa2001316 |
| Illness onset to hospital admission | 203 | | | | | | | | Days | 5.8 | | | 1 | 20 | Wuhan (China) | | 54 | | 53.2 | Data from Jan 1 – Feb 10, 2020 | Chen et al. 2020 | https://doi.org/10.1093/gerona/glaa089 |
| Illness onset to hospital admission | 191 | | | | | | | | Days | 11 | | | 8 | 14 | Patients from Wuhan Hospitals | | 56 | | 62 | Data from Dec 29 to Jan 31. | Zhou et al. 2020 | https://doi.org/10.1016/S0140-6736(20)30566-3 |
| Illness onset to hospital admission * | 291 | | | | | | | | Days | 5 | | | 3 | 8 | Hunan (China) | | 46 | | 49.8 | Data from Jan 23 to Feb 14, 2020 | Chen X et al. 2020 | https://doi.org/10.1101/2020.03.03.20030353 |
| Illness onset to hospital admission | 77 | | | | | | | | Days | 5 | | | 3 | 6 | Beijing (China) | | | | | Patients admitted from Jan 21 to February 8, 2020 | Zhao Wen et al. 2020 | https://doi.org/10.15212/CVIA.2021.0019 |
| Illness onset to hospital admission ** | 8 | 2.25 | | | | 0.7 1 | | | Days | | | | | | Anhui (China) | | 22 | | 75 | Data up to Jan 27, 2020 | Huang et al. 2020 | https://doi.org/10.1016/j.jinf.2020.03.006 |
| Onset to hospitalization (symptom-based surveillance) | 292 | 4.64 | | 4.23 | 5.1 | 4.3 7 | | | Days | | | | | | China | | | | | Lognormal distribution Data from Jan 14 to Feb 9, 2020 | Bi et al. 2020 | https://doi.org/10.1016/S1473-3099(20)30287-5 |
| Onset to admission | 54 | 5.81 | | 4.95 | 6.68 | | | | Days | | | | | | Beijing (China) | | | | | Patients admitted from Dec 27, 2019 to Feb 18, 2020 | Yang et al. 2020 | https://doi.org/10.1101/2020.02.28.20028068 |
| Onset to hospitalization (contact-based surveillance) | 87 | 2.57 | | 2.06 | 3.16 | 1.8 3 | | | Days | | | | | | China | | | | | Lognormal distribution Data from Jan 14 to Feb 9, 2020 | Bi et al. 2020 | https://doi.org/10.1016/S1473-3099(20)30287-5 |
| Illness onset to hospital admission (all) | 403 | | | | | | | | Days | 9 | | | 7 | 12 | Wuhan (China) | | 56 | | 47.9 | Data from Jan 30 to Feb 25, 2020 (outcomes included) | Luo et al. 2020 | https://doi.org/10.1101/2020.03.19.20033175 |

| | | | | | | | | | | | | | | | | | | | | | | |
|---|---|---|---|---|---|---|---|---|---|---|---|---|---|---|---|---|---|---|---|---|---|---|
| **Onset of symptoms/illness onset to hospital admission (survivors) (days)** | | | | | | | | | | | | | | | | | | | | | | |
| Symptom onset to hospitalization (survivors) | 37 | 4.1 | | | | 3.7 | | | Days | | | | | | Outside Hubei (China) | 44.3 | | | 45.9 | Period unknown, considered both Period unknown | Tian Suochen et al .2020 | https://doi.org/10.3389/fmed.2020.00210 |
| Symptom onset to hospitalization (survivors) | 116 | | | | | | | | Days | 7 | | | 5 | 10 | Wuhan (China) | | 49 | | 44 | From Jan 1 to Feb 21, 2020 | Deng et al. 2020 | https://doi.org/10.1097/CM9.0000000000000824 |
| Symptom onset to hospitalization (survivors) | 161 | | | | | | | | Days | 9 | | | 6 | 12 | Wuhan (China) | | 51 | | 55 | Data from Jan 13 to Feb 12, 2020 | Chen T et al. 2020 | http://dx.doi.org/10.1136/bmj.m1091 |
| Illness onset to hospital admission (survivor) | 137 | | | | | | | | Days | 11 | | | 8 | 13 | Patients from Wuhan Hospital (China) | | 52 | | 59 | Data from Dec 29 to Jan 31, 2020 | Zhou Fei et al. 2020 | https://doi.org/10.1016/S0140-6736(20)30566-3 |
| Illness onset to hospital admission (survivor) | 64 | | | | | | | | Days | 5 | | | 4 | 6 | Beijing (China) | | | | | Patients admitted from Jan 21 to February 8, 2020 | Zhao Wen et al. 2020 | https://doi.org/10.15212/CVIA.2021.0019 |
| Illness onset to hospital admission among living cases | 155 | 3.4 | | 2.7 | 4.2 | | | | Days | 1.8 | 1.4 | 2.3 | | | Residents from other locations who travelled to Wuhan + Wuhan residents | | | | | Accounting without right truncation. Weibull Data up to Jan 31, 2020 | Linton et al. 2020 | https://doi.org/10.3390/jcm9020538 |
| Illness onset to hospital admission | 33 | | | | | | | | Days | 9.5 | | | 7 | 10.25 | Wuhan (China) | | | | | Cases fom Dec 20, 2019 to Feb 20,2020 | Zhang Lin et al. 2020 | https://doi.org/10.1017/s0950268820002010 |
| Illness onset to hospital admission among living cases | 155 | 3.3 | | 2.7 | 4 | | | | Days | 1.7 | 1.2 | 2.2 | | | Residents from other locations who travelled to Wuhan + Wuhan residents | | | | | Accounting without right truncation. Gamma Data up to Jan 31, 2020 | Linton et al. 2020 | https://doi.org/10.3390/jcm9020538 |
| Illness onset to hospital admission among living cases | 155 | 3.9 | | 2.9 | 5.3 | | | | Days | 1.5 | 1.2 | 1.9 | | | Residents from other locations who travelled to Wuhan + Wuhan residents | | | | | Accounting without right truncation. Lognormal Data up to Jan 31, 2020 | Linton et al. 2020 | https://doi.org/10.3390/jcm9020538 |
| Illness onset to hospital admission among living cases | | 9.7 | | 5.4 | 17 | | | | Days | 2.6 | 1.9 | 3.8 | | | Residents from other locations who travelled to Wuhan + Wuhan residents | | | | | Right-truncated incubation. Lognormal Data up to Jan 31, 2020 | Linton et al. 2020 | https://doi.org/10.3390/jcm9020538 |
| Illness onset to hospital admission (survivor) | 303 | | | | | | | | Days | 9 | | | 6 | 11 | Wuhan (China) | | 49 | | 44,9 | Data from Jan 30 to Feb 25, 2020 (outcomes included) | Luo et. al 2020 | https://doi.org/10.1101/2020.03.19.20033175 |
| **Onset of symptoms/illness onset to hospital admission (non-survivor) (days)** | | | | | | | | | | | | | | | | | | | | | | |
| Symptom onset to hospitalization (non-survivor) | 109 | | | | | | | | Days | 10 | | | 6.5 | 12 | Wuhan (China) | | 69 | | 67 | From Jan 1 to Feb 21, 2020 | Deng et al. 2020 | https://doi.org/10.1097/CM9.0000000000000824 |
| Symptom onset to hospital admission (non-survivor) | 85 | 10.1 | | | | 6.2 | | | Days | | | | | | Wuhan (China) | | 65.8 | | 72.9 | Data from Jan 9 to Feb 15, 2020 | Du Yingzhen et al. 2020 | https://doi.org/10.1164/rccm.202003-0543OC |
| Symptom onset to hospital admission (non-survivor) | 109 | 9.7 | | | | 5.3 | | | Days | | | | | | Wuhan (China) | 70 | | | 67.9 | Data up to February 24, 2020 | Du Rong-Hui et al. 2020 | https://doi.org/10.1513/AnnalsATS.202003-225OC |
| Symptom onset to hospital admission (non-survivor) | 113 | | | | | | | | Days | 10 | | | 7 | 13 | Wuhan (China) | | 68 | | 73 | Data from Jan 13 to Feb 12, 2020 | Chen T et al. 2020 | http://dx.doi.org/10.1136/bmj.m1091 |

| | | | | | | | | | | | | | | | | | | | | | | |
|---|---|---|---|---|---|---|---|---|---|---|---|---|---|---|---|---|---|---|---|---|---|---|
| Onset of symptoms to hospital admission (dead cases) | 101 | | | | | | | | Days | 11 | | | 8 | 13.5 | Patients from Wuhan Hospitals | 65.46 | | | 64 | Cases from Dec 30, 2019 to Feb 20, 2020 | Zhang Lin et al. 2020 | https://doi.org/10.1017/s0950268820002010 |
| Illness onset to hospital admission among deceased cases | 34 | 6.5 | | 5.2 | 8 | | | | Days | 5.1 | 4.1 | 6.3 | | | Residents from other locations who travelled to Wuhan + Wuhan residents | | | | | Accounting without right truncation. Weibull Data up to Jan 31, 2020 | Linton et al. 2020 | https://doi.org/10.3390/jcm9020538 |
| Illness onset to hospital admission among deceased cases | 34 | 6.5 | | 5.2 | 8 | | | | Days | 5.6 | 4.5 | 6.9 | | | Residents from other locations who travelled to Wuhan + Wuhan residents | | | | | Accounting without right truncation. Gamma Data up to Jan 31, 2020 | Linton et al. 2020 | https://doi.org/10.3390/jcm9020538 |
| Illness onset to hospital admission among deceased cases | 34 | 6.2 | | 5 | 7.8 | | | | Days | 5.1 | 4.1 | 6.3 | | | Residents from other locations who travelled to Wuhan + Wuhan residents | | | | | Accounting without right truncation. Lognormal Data up to Jan 31, 2020 | Linton et al. 2020 | https://doi.org/10.3390/jcm9020538 |
| Illness onset to hospital admission among deceased cases | | 6.6 | | 5.2 | 8.8 | | | | Days | 5.3 | 4.2 | 6.8 | | | Residents from other locations who travelled to Wuhan + Wuhan residents | | | | | Right-truncated incubation. Lognormal Data up to Jan 31, 2020 | Linton et al. 2020 | https://doi.org/10.3390/jcm9020538 |
| Illness onset to hospital admission (non-survivor) | 5 | | | | | | | | Days | 4 | | | 2 | 6 | Beijing (China) | | | | | Patients admitted from Jan 21 to February 8, 2020 | Zhao Wen et al. 2020 | https://doi.org/10.15212/CVIA.2021.0019 |
| Illness onset to hospital admission (non-survivor) | 54 | | | | | | | | Days | 11 | | | 8 | 15 | Patients from Wuhan Hospital (China) | | 69 | | 70 | Data from Dec 29 to Jan 31, 2020 | Zhou Fei et al. 2020 | https://doi.org/10.1016/S0140-6736(20)30566-3 |
| Illness onset to hospital admission (non-survivor) | 100 | | | | | | | | Days | 9 | | | 7 | 13 | Wuhan (China) | | 71 | | 57 | Data from Jan 30 to Feb 25, 2020 (outcomes included) | Luo et al. 2020 | https://doi.org/10.1101/2020.03.19.20033175 |
| **Onset of symptoms to ICU admission (days)** | | | | | | | | | | | | | | | | | | | | | | |
| Onset of symptoms to ICU admission | 44 | | | | | | | | Days | 10 | | | 7 | 13 | Wuhan (China) | | | | | Data from Jan 2 to Feb 10, 2020 | Zhang Guqin et al. 2020 | https://doi.org/10.1016/j.jcv.2020.104364 |
| Onset of symptoms to ICU admission | 52 | | | | | | | | Days | 9.5 | | | 7 | 12.5 | Wuhan (China) | | | | | Data from late Dec, 2019 to Jan 26, 2020 | Yang et al. 2020 | https://doi.org/10.1016/S2213-2600(20)30079-5 |
| Onset of symptoms to ICU admission | 8 | | | | | | | | Days | 10 | | | 9 | 11 | Hong Kong | 64.5 | | | 50 | Patients admitted from Jan 22 to Feb 11, 2020 | Ling Lowell et al. 2020 | https://www.ncbi.nlm.nih.gov/pubmed/32248675 |
| Onset of symptoms to ICU admission | 13 | | | | | | | | Days | 7 | | | 4 | 8 | Patients from Wuhan Hospital (China) | | 49 | | 85 | Data from Dec 1 to Jan 2,2020 | Huang Chaolin et al. 2020 | https://doi.org/10.1016/S0140-6736(20)30183-5 |
| Onset of symptoms to ICU admission | 36 | | | | | | | | Days | 10 | | | 6 | 12 | Wuhan (China) | | 66 | | 61.1 | Data from Jan 1 to Jan 28, 2020 | Wang Dawei et al. 2020 | https://doi.org/10.1001/jama.2020.1585 |
| Onset of symptoms to ICU admission | 34 | | | | | | | | Days | 10 | | | 7 | 11.3 | Hangzhou (China) | | 66 | | 67.6 | Data from Jan 22 to March 5, 2020 | Zheng Xi et al. 2020 | https://doi.org/10.1631/jzus.b2000174 |
| Onset of symptoms to ICU admission | 48 | | | | | | | | Days | 7 | | | 5 | 12 | Vitoria (Spain) | | 63 | | 56.3 | | Barrasa et al. 2020 | https://doi.org/doi:10.1016/j.accpm.2020.04.001 |
| **Onset of symptoms to ICU admission (non-survivors)(days)** | | | | | | | | | | | | | | | | | | | | | | |
| Onset of symptoms to ICU admission (non-survivor) | 51 | 14 | | | | 7.6 | | | Days | | | | | | Wuhan (China) | 68.4 | | | | Data up to February 24, 2020 | Du Rong-Hui et al. 2020 | https://doi.org/10.1513/AnnalsATS.202003-225OC |
| **Hospital stay length (days)** | | | | | | | | | | | | | | | | | | | | | | |
| Hospital stay (all) * | 1099 | 12.8 | | | | | | | days | 12 | | | 10 | 14 | Mainland China | | 47 | | | Data up to Jan 29, 2020 | Guan et al. 2020 | https://doi.org/10.1056/NEJMoa2002032 |

| | | | | | | | | | | | | | | | | | | | | | | |
|---|---|---|---|---|---|---|---|---|---|---|---|---|---|---|---|---|---|---|---|---|---|---|
| Hospital stay (all) | 201 | | | | | | | | days | 13 | | | 10 | 16 | Patients from Wuhan Hospital (China) | | 51 | | 63.7 | Patients admitted from Dec 25, 2019 to Jan 26, 2020. Outcome follow-up to Feb 13, 2020 | Wu Chaomin et al. 2020 | https://doi.org/10.1001/jamainternmed.2020.0994 |
| Hospital stay (all) | 191 | | | | | | | | days | 11 | | | 7 | 14 | Patients from Wuhan Hospital (China) | | 51 | | 62 | Data from Dec 29 to Jan 31, 2020 | Zhou Fei et al. 2020 | https://doi.org/10.1016/S0140-6736(20)30566-3 |
| Hospital stay (all) | 7 | 11.7 | | | | | | | days | | | | | | Chongqing (China) | | | 0-14 | | Data from January to March, 2020 | Tao et al. 2020 | https://doi.org/10.1101/2020.03.16.20037259 |
| Hospital stay (all) | 21 | 15.3 | | | | | | | days | | | | | | Chongqing (China) | | | 15-29 | | Data from January to March, 2020 | Tao et al. 2020 | https://doi.org/10.1101/2020.03.16.20037259 |
| Hospital stay (all) | 39 | 15.5 | | | | | | | days | | | | | | Chongqing (China) | | | 30-39 | | Data from January to March, 2020 | Tao et al. 2020 | https://doi.org/10.1101/2020.03.16.20037259 |
| Hospital stay (all) | 36 | 17.5 | | | | | | | days | | | | | | Chongqing (China) | | | 40-49 | | Data from January to March, 2020 | Tao et al. 2020 | https://doi.org/10.1101/2020.03.16.20037259 |
| Hospital stay (all) | 33 | 18 | | | | | | | days | | | | | | Chongqing (China) | | | 50-59 | | Data from January to March, 2020 | Tao et al. 2020 | https://doi.org/10.1101/2020.03.16.20037259 |
| Hospital stay (all) | 19 | 17.9 | | | | | | | days | | | | | | Chongqing (China) | | | 60-69 | | Data from January to March, 2020 | Tao et al. 2020 | https://doi.org/10.1101/2020.03.16.20037259 |
| Hospital stay (all) | 12 | 20.2 | | | | | | | days | | | | | | Chongqing (China) | | | ≥ 70 | | Data from January to March, 2020 | Tao et al. 2020 | https://doi.org/10.1101/2020.03.16.20037259 |
| Hospital stay (all) | 197 | 15.08 | | | | 17. 47 | | | days | | | | | | Hubei (China) | | | | | Patients discharged from Jan 17 to Feb 26, 2020 | Zhou Fating et al. 2020 | https://doi.org/10.1101/2020.03.26.20041426 |
| Hospital stay (all) | 73 | | | | | | | | days | 13 | | | 10 | 18 | Wuhan (China) | | 67 | | 61.1 | Patients admitted from Dec 24 to Feb 7, 2020 | Tang et al. 2020 | https://doi.org/10.1016/j.chest.2020.03.032 |
| Hospital stay (all) * | 94 | 14.28 | | 13.61 | 14.95 | | | | days | | | | | | Shenzhen (China) | | 40 | | 45 | Patients admitted from Jan 5 to Feb 4, 2020. Outcomes Follow up Feb 13, 2020 | Yuan et al. 2020 | https://doi.org/10.1007/s00011-020-01342-0 |
| Hospital stay (all) | 339 | | | | | | | | days | 28 | | | 15 | 28 | Wuhan (China) | 71 | | | 49 | Data up to Feb 28, 2020 | Wang et al. 2020 | https://doi.org/10.1016/j.jinf.2020.03.019 |
| Hospital stay (all) | 199 | | | | | | | | days | 15 | | | 12 | 17 | Wuhan (China) | | 58 | | 39.7 | Data from Jan 18 to Feb 3, 2020 | Cao Bin et al. 2020 | https://doi.org/10.1056/NEJMoa2001282 |
| Hospital stay (all) | 24 | | | | | | | | days | 12 | | | 8 | 18 | Seattle region | 64 | | | 63 | | Bhatraju et al. 2020 | https://doi.org/10.1056/NEJMoa2004500 |
| Hospital stay (all) | 102 | | | | | | | | days | 11 | | | 7 | 15 | Wuhan (China) | | 54 | | 52 | Patients admitted from Jan 3 to Feb 1, 2020. Outcomes follow up to Feb 15,2020 | Cao Jianlei et al. 2020 | https://doi.org/10.1093/cid/ciaa243 |
| Hospital stay (all) | 538 | | 3-41 | | | | | | days | 19 | | | 14 | 23 | Sichuan (China) | | | | 53 | Data from January to March, 2020 | Wang et al. 2020 | https://doi.org/10.1101/2020.04.07.20057299 |
| Hospital stay (all) | 24 | 16 | | | 6.15 | | | | days | | | | | | Liaocheng city (China) | 48 | | | 37.5 | Period unknown, but at least one admission after Jan 9, 2020 | Tian Souchen et al .2020 | https://doi.org/10.1186/s12889-021-10713-z |
| Hospital stay (all) | 75 | | | | | | | | days | 11 | | | 5 | 15 | Zhejiang (China) | 46.37 | | | 55 | Data from January to Feb 20, 2020 | Hong et al. 2020 | http://dx.doi.org/10.21037/atm.2020.03.147 |
| Hospital stay (all) | 25 | 19.7 | | | 4.6 | | | | days | | | | | | Wuhu (China) | 46.8 | | | 56 | Patients admitted from jan 22 to Feb 28, 2020 | Huang Guoan et al. 2020 | https://doi.org/10.2214/AJR.20.23078 |
| Hospital stay (all) | 203 | | | | | | | | days | 11 | | | 1 | 45 | Wuhan (China) | | 54 | | 53.2 | Data from Jan 1 to Feb 20, 2020 | Chen TL et al. 2020 | https://doi.org/10.1093/gerona/glaa089 |
| Hospital stay (all) | 58 | 19.8 | | | | 10. 82 | | | days | | | | | | Wuhan (China) | 42.6 | | | 44.8 | Patients admitted from Jan 1 to Feb 23, 2020 | Meng et al. 2020 | https://doi.org/10.1016/j.jinf.2020.04.004 |
| Hospital stay * | 51 | | | | | | | | days | 12 | | | 9 | 13 | Chongqing (China) | | 45 | | 62.7 | Data from Jan 20 To Feb 11, 2020 | Liu Lei et al. 2020 | https://doi.org/10.1101/2020.02.20.20025536 |

| | | | | | | | | | | | | | | | | | | | | | | |
|---|---|---|---|---|---|---|---|---|---|---|---|---|---|---|---|---|---|---|---|---|---|---|
| Hospital stay | 18 | 13.39 | | | | 2.64 | | | days | | | | | | Tibet | 29.67 | 31 | | 61.11 | Patients recruited from Jan 26 to Feb 28, 2020 | Song et al. 2020 | https://doi.org/10.1101/2020.03.27.20043836 |
| Hosptial stay | 20 | 14.3 | | | | | | | days | | | | | | Chongqing (China) | | 47 | | | Data from January to March, 2020 | Tao et al. 2020 | https://doi.org/10.1101/2020.03.16.20037259 |
| Hosptial stay | 77 | | | | | | | | days | 13 | | | 10 | 18 | Beijing (China) | | | | | Patients admitted from Jan 21 to February 8, 2020 Outcomes follow-up to Feb 29,2020 | Zhao Wen et al. 2020 | https://doi.org/10.15212/CVIA.2021.0019 |
| **Hospital stay length (survivors)(days)** | | | | | | | | | | | | | | | | | | | | | | |
| Hospital stay (survivors) | 6 | 11.5 | | 8 | 17.3 | | | | days | | | | | | China | 34.5 | | | 83.3 | January 2020 | Sanche et al. 2020 | https://doi.org/10.3201/eid2607.200282 |
| Hospital stay (survivors) | 137 | | | | | | | | days | 12 | | | 9 | 15 | Patients from Wuhan Hospital (China) | | 52 | | 59 | Data from Dec 29 to Jan 31, 2020 | Zhou Fei et al. 2020 | https://doi.org/10.1016/S0140-6736(20)30566-3 |
| Hospital stay (survivors) | 47 | | | | | | | | days | 10 | | | 7 | 14 | Wuhan (China) | | | | | Data from Jan 1 to Feb 3, 2020 | Wang Dawei et al. 2020 | https://doi.org/10.1001/jama.2020.1585 |
| Hospital stay (survivors) | 274 | | | | | | | | days | 28 | | | 26 | 29 | Wuhan (China) | | 68 | | 46.4 | Data up to Feb 28, 2020 | Wang Lang et al. 2020 | https://doi.org/10.1016/j.jinf.2020.03.019 |
| Hospital stay (survivors) | 85 | | | | | | | | days | 11 | | | 7 | 14 | Wuhan (China) | | 53 | | 47.1 | Patients admitted from Jan 3 to Feb 1, 2020. Outcomes follow up to Feb 15,2020 | Cao Jianlei et al. 2020 | https://doi.org/10.1093/cid/ciaa243 |
| Hospital stay (survivors) | 12 | | | | | | | | days | 17 | | | 16 | 23 | Seattle region (USA) | | | | | | Bhatraju et al. 2020 | https://doi.org/10.1056/NEJMoa2004500 |
| Hospital stay (survivors) | 10 | | | | | | | | days | 20 | | | 19 | 21 | Macau | | 54 | | 30 | Patients enrolled from Jan 21 to Feb 16, 2020 | Lo et al. 2020 | https://doi.org/10.7150/ijbs.45357 |
| Hospital stay (survivors) | 123 | | | | | | | | days | 10 | | | 7 | 15 | Wuhan (China) | | | | | Patients admitted from Jan 1 to Feb 5, 2020 | Mo et al 2020 | https://doi.org/10.1093/cid/ciaa270 |
| Hospital stay (Survivors) | 93 | 13.3 | | | | 6.0 | | | days | | | | | | Singapore | | | | | Data from Jan 18 to Feb 28,2020 | Tindale et al. 2020 | https://doi.org/10.7554/eLife.57149 |
| Hospital stay (survivors) | 56 | | | | | | | | days | 17 | | | 11 | 20 | Jilin (China) | | 40 | | 55.4 | Data from Jan 5 to March 5, 2020 | Tian Suochen et al .2020b | https://doi.org/10.1007/s12250-020-00317-z |
| Hospital stay (survivors) | 116 | | | | | | | | days | 16 | | | 12 | 20 | Wuhan (China) | 40.33 | 40 | | 44 | From Jan 1 to after Feb 21, 2020 | Deng et al. 2020 | https://doi.org/10.1097/CM9.0000000000000824 |
| Hospital stay (survivors) | 64 | | | | | | | | days | 13 | | | 10 | 18 | Beijing (China) | 52 | | | 44.2 | Patients admitted from Jan 21 to February 8, 2020 Outcomes follow-up to Feb 29,2020 | Zhao Wen et al. 2020 | https://doi.org/10.15212/CVIA.2021.0019 |
| Hospital stay (survivors) | 37 | 16.1 | | | | 6.2 | | | days | | | | | | Outside Hubei (China) | 44.3 | | | | Period unknown, considered both | Tian Suochen et al .2020b | https://doi.org/10.3389/fmed.2020.00210 |
| Hospital stay (survivors) | 18 | 17.7 | 8-35 | | | 7.7 | | | days | | | | | | Hong Kong | | | | | Data from Jan 26 to Feb 28, 2020 | Leung et al. 2020 | https://doi.org/10.3201/eid2701.201543 |
| Hospital stay (survivors) * | 55 | | | | | | | | days | 16 | | | 13 | 21 | Jiangsu (China) | | 45 | | 49.1 | Data from Jan 23 to April 6 | Jiang et al. 2020 | https://doi.org/10.1101/2020.04.10.20060335 |
| Hospital stay (survivors) | 125 | 14.8 | | | | 4.16 | | | days | | | | | | Anhui (China) | 38.76 | | | 56.8 | Data from Jan 20 to Feb 18,2020 | Wang R et al. 2020 | https://doi.org/10.1016/j.ijid.2020.03.070 |
| Hospital stay (survivors) | 80 | 8 | | | | | | | days | | | | | | Jiangsu (China) | 46.1 | | | 48.75 | Patients enrolled from Jan 22 to Feb 14, 2020 | Wu et al. 2020 | https://doi.org/10.1093/cid/ciaa199 |
| Time from hospital admission to discharge | 160 | | | | | | | | days | 17 | | | 12 | 21 | Hainan (China) | | | | | Data from Jan 22 to March 13, 2020 | Yan et al. 2020 | https://doi.org/10.1101/2020.03.19.20038539 |
| Time from hospital admission to discharge * | 103 | | | | | | | | days | 15 | | | 11.5 | 16 | Chongqing (China) | | 48 | | 55.8 | Data from Jan 19 to Feb 16, 2020 | Qi Di et al. 2020 | https://doi.org/10.1101/2020.03.01.20029397 |

| | | | | | | | | | | | | | | | | | | | | | | |
|---|---|---|---|---|---|---|---|---|---|---|---|---|---|---|---|---|---|---|---|---|---|---|
| Time from hospital admission to discharge * | 159 | | | | | | | | days | 12 | | | 10 | 15 | Hunan (China) | | 46 | | 49.8 | Data from Jan 23 to Feb 14, 2020. Outcomes until Feb 20, 2020 | Chen X et al. 2020 | https://doi.org/10.1101/2020.03.03.20030353 |
| Time from hospital admission to discharge | 161 | | | | | | | | days | 16 | | | 14 | 19 | Wuhan (China) | | 51 | | 55 | Data from Jan 13 to Feb 28, 2020 | Chen T et al. 2020 | http://dx.doi.org/10.1136/bmj.m1091 |
| Time from hospital admission to discharge | 11 | 14.5 | | | | 8.7 | | | days | | | | | | Thailand | | 61 | | 54.5 | Patients admitted from Jan 8-31, 2020 | Pongpirul et al. 2020 | https://doi.org/10.3201/eid2607.200598 |
| Time from hospital admission to discharge ** | 4 | 18 | | | | 2.1 6 | | | days | | | | | | Anhui (China) | | 21.5 | | 75 | Data from Jan 22 to Feb 20, 2020 | Huang et al. 2020 | https://doi.org/10.1016/j.jinf.2020.03.006 |
| **Hospital stay lenth (non-survivors)(days)** | | | | | | | | | | | | | | | | | | | | | | |
| Hospital stay (non-survivors) | 54 | | | | | | | | days | 7.5 | | | 5 | 11 | Patients from Wuhan Hospital (China) | | 69 | | 70 | Data from Dec 29 to Jan 31, 2020 | Zhou Fei et al. 2020 | https://doi.org/10.1016/S0140-6736(20)30566-3 |
| Hospital stay (non-survivors) | 65 | | | | | | | | days | 5 | | | 3 | 8 | Wuhan (China) | | 76 | | 60 | Data up to Feb 28, 2020 | Wang Lang et al. 2020 | https://doi.org/10.1016/j.jinf.2020.03.019 |
| Hospital stay (non-survivors) | 17 | | | | | | | | days | 9 | | | 6 | 17 | Wuhan (China) | | 72 | | 76.5 | Patients admitted from Jan 3 to Feb 1, 2020. Outcomes follow up to Feb 15,2020 | Cao Jianlei et al. 2020 | https://doi.org/10.1093/cid/ciaa243 |
| Hospital stay (non-survivors) | 22 | | | | | | | | days | 10.5 | | | 8 | 6 | Wuhan (China) | | | | | Confirmed patients from Jan 1 to Feb 5, 2020 | Mo et al 2020 | https://doi.org/10.1093/cid/ciaa270 |
| Hospital stay (non-survivors) | 5 | | | | | | | | days | 10 | | | 7 | 17 | Beijing (China) | 52 | | | 44.2 | Patients admitted from Jan 21 to February 8, 2020 Outcomes follow-up to Feb 29,2020 | Zhao Wen et al. 2020 | https://doi.org/10.15212/CVIA.2021.0019 |
| Hospital stay (non-survivors) | 109 | | | | | | | | days | 8 | | | 4 | 13 | Wuhan (China) | | 69 | | 67 | From Jan 1 to after Feb 21, 2020 | Deng et al. 2020 | https://doi.org/10.1097/CM9.0000000000000824 |
| Hospital stay in general wards (non-survivors) | 101 | | | | | | | | days | 0 | | | 0 | 4 | Patients from Wuhan Hospital (China) | 65.46 | | | 64 | Cases from Dec 30, 2019 to Feb 16, 2020 | Zhang et al. 2020 | https://doi.org/10.1017/s0950268820002010 |
| Hospital admission to death | 39 | 8.9 | | 7.3 | 10.4 | | | | Days | 8 | 6.2 | 9.8 | | | Residents from other locations who travelled to Wuhan + Wuhan residents | | | | | Accounting without right truncation. Weibull Data up to Jan 31, 2020 | Linton et al. 2020 | https://doi.org/10.3390/jcm9020538 |
| Hospital admission to death | 39 | 8.8 | | 7.2 | 10.8 | | | | Days | 7.6 | 6 | 9.3 | | | Residents from other locations who travelled to Wuhan + Wuhan residents | | | | | Accounting without right truncation. Gamma Data up to Jan 31, 2020 | Linton et al. 2020 | https://doi.org/10.3390/jcm9020538 |
| Hospital admission to death | 39 | 8.6 | | 6.8 | 10.8 | | | | Days | 6.7 | 5.3 | 8.3 | | | Residents from other locations who travelled to Wuhan + Wuhan residents | | | | | Accounting without right truncation. Lognormal Data up to Jan 31, 2020 | Linton et al. 2020 | https://doi.org/10.3390/jcm9020538 |
| Hospital admission to death | | 13 | | 8.7 | 20.9 | | | | Days | 9.1 | 6.7 | 13.7 | | | Residents from other locations who travelled to Wuhan + Wuhan residents | | | | | Right-truncated incubation. Lognormal Data up to Jan 31, 2020 | Linton et al. 2020 | https://doi.org/10.3390/jcm9020538 |
| Hospital admission to death | 23 | 11.2 | | 8.7 | 14.9 | | | | Days | | | | | | Wuhan (China) | | | | 69.6 | January 2020 | Sanche et al. 2020 | https://doi.org/10.3201/eid2607.200282 |
| Hospital admission to death | 85 | 6.35 | | | | 4.5 1 | | | Days | | | | | | Wuhan (China) | | 65.8 | | 72.9 | Fatal cases from Jan 9 to Feb 15, 2020 | Du et al. 2020 | https://doi.org/10.1164/rccm.202003-0543OC |
| Hospital admission to death | 109 | 14.1 | | | | 8.8 | | | Days | | | | | | Wuhan (China) | 70.7 | | | | Data up to February 24, 2020 | Du Rong-Hui et al. 2020 | https://doi.org/10.1513/AnnalsATS.202003-225OC |

| | | | | | | | | | | | | | | | | | | | | | | |
|---|---|---|---|---|---|---|---|---|---|---|---|---|---|---|---|---|---|---|---|---|---|---|
| Hospital admission to death | 33 | 10.1 | | | | 5.4 | | | Days | | | | | | China | | | | | Gamma distribution. Data up to Feb 7, 2020 | Mizumoto et al. 2020 | https://doi.org/10.3201/eid2606.200233 |
| Hospital admission to death | 66 | | 0-16 | | | | | | Days | 5 | | | | | Republic of Korea | | | | | Data up to March 12, 2020 | Choe et al 2020 | https://doi.org/10.24171/j.phrp.2020.11.2.05 |
| Hospital admission to death | 6 | | | | | | | | Days | 13 | | | 1.5 | 28.3 | Hainan (China) | | | | | Data from Jan 22 to March 13, 2020 | Yan et al. 2020 | https://doi.org/10.1101/2020.03.19.20038539 |
| Hospital admission to death | 113 | | | | | | | | Days | 5 | | | 3 | 9.3 | Wuhan (China) | | 68 | | 73 | Data from Jan 13 to Feb 28, 2020 | Chen T et al. 2020 | http://dx.doi.org/10.1136/bmj.m1091 |
| Hospital admission to death | 133 | | | | | | | | Days | 10 | | | 6 | 15 | Wuhan (China) | | 70 | | 55.6 | Patients enrolled from Jan 25 to Feb 25, 2020 | Wang Yang et al. 2020 | https://doi.org/10.1164/rccm.202003-0736LE |
| Hospital admission to death ** | 40 | 11.83 | | | | 10.8 | | | Days | | | | | | Wuhan (China) | 70.5 | | | 67.5 | Admissions until Jan 23. Deaths until Jan 28, 2020 | Wu Joseph et al. 2020 | https://doi.org/10.1038/s41591-020-0822-7 |
| **ICU stay length (days)** | | | | | | | | | | | | | | | | | | | | | | |
| ICU Stay (all) | 191 | | | | | | | | days | 8 | | | 4 | 12 | Patients from Wuhan Hospital (China) | | 51 | | 62 | Data from Dec 29 to Jan 31, 2020 | Zhou Fei et al. 2020 | https://doi.org/10.1016/S0140-6736(20)30566-3 |
| ICU stay (all) | 199 | | | | | | | | days | 10 | | | 5 | 14 | Wuhan (China) | | 58 | | 60.3 | Data from Jan 18 to Feb 3, 2020 | Cao Bin et al. 2020 | https://doi.org/10.1056/NEJMoa2001282 |
| ICU stay (all) | 24 | | | | | | | | days | 9 | | | 4 | 12 | Seattle región (USA) | | | | | | Bhatraju et al. 2020 | https://doi.org/10.1056/NEJMoa2004500 |
| **ICU stay length (survivors)(days)** | | | | | | | | | | | | | | | | | | | | | | |
| ICU Stay (survivors) | 137 | | | | | | | | days | 7 | | | 2 | 9 | Patients from Wuhan Hospital (China) | | 52 | | 59 | Data from Dec 29 to Jan 31, 2020 | Zhou Fei et al. 2020 | https://doi.org/10.1016/S0140-6736(20)30566-3 |
| ICU Stay (survivors) | 44 | | | | | | | | days | 10 | | | 8 | 17 | Wuhan (China) | | 58 | | 61.1 | Data from Jan 18 to Feb 3, 2020 | Cao Bin et al. 2020 | https://doi.org/10.1056/NEJMoa2001282 |
| ICU Stay (survivors) | 12 | | | | | | | | days | 14 | | | 4 | 17 | Seattle region | | | | | | Bhatraju et al. 2020 | https://doi.org/10.1056/NEJMoa2004500 |
| ICU Stay (survivors) | 23 | | | | | | | | days | 8 | | | 5 | 13 | Wuhan (China) | | 62 | | 65.2 | Data from Jan 2 to Feb 10, 2020 Outcomes follow up to Feb 15, 2020 | Zhang Guqin et al. 2020 | https://doi.org/10.1016/j.jcv.2020.104364 |
| ICU Stay (survivors) | 8 | | | | | | | | days | 12 | | | 8 | 16 | Hong Kong | | 64 | | 50 | Patients admitted from Jan 22 to Feb 11, 2020 | Ling Lowell et al. 2020 | https://www.ncbi.nlm.nih.gov/pubmed/32248675 |
| **ICU stay length (non-survivors) (days)** | | | | | | | | | | | | | | | | | | | | | | |
| ICU Stay (non-survivor) | 52 | | | | | | | | Days | 7 | | | 3 | 11 | Wuhan (China) | | | | | Data from late Dec, 2019 to Feb 8, 2020 | Yang et al. 2020 | https://doi.org/10.1016/S2213-2600(20)30079-5 |
| ICU Stay (non-survivor) | 54 | | | | | | | | days | 8 | | | 4 | 12 | Patients from Wuhan Hospital (China) | | 69 | | 70 | Data from Dec 29 to Jan 31, 2020 | Zhou Fei et al. 2020 | https://doi.org/10.1016/S0140-6736(20)30566-3 |
| ICU Stay (non-survivor) | 155 | | | | | | | | days | 10 | | | 4 | 14 | Wuhan (China) | | 58 | | 59 | Data from Jan 18 to Feb 3, 2020 | Cao Bin et al. 2020 | https://doi.org/10.1056/NEJMoa2001282 |
| ICU Stay (non-survivor) | 9 | | | | | | | | days | 11 | | | 4.5 | 14.5 | Wuhan (China) | | 76 | | 77.8 | Data from Jan 2 to Feb 10, 2020 Outcomes follow up to Feb 15, 2020 | Zhang Guqin et al. 2020 | https://doi.org/10.1016/j.jcv.2020.104364 |
| ICU stay (non-survivor) | 51 | 11.8 | | | | 8.8 | | | days | | | | | | Wuhan (China) | 68.4 | | | 70.6 | Data up to February 24, 2020 | Du Rong-Hui et al. 2020 | https://doi.org/10.1513/AnnalsATS.202003-225OC |
| **Onset of symptoms/Illness onset to death (days)** | | | | | | | | | | | | | | | | | | | | | | |
| Onset of symptoms to death | 24 | 17.8 | | 16.9 | 19.2 | | | | Days | | | | | | Mainland China | | | | | Data up to Feb 8, 2020 | Verity et al. 2020 | https://doi.org/10.1016/S1473-3099(20)30243-7 |

| | | | | | | | | | | | | | | | | | | | | | | |
|---|---|---|---|---|---|---|---|---|---|---|---|---|---|---|---|---|---|---|---|---|---|---|
| Onset of symptoms to death | 6 | | | | | | | | Days | 17 | | | 10.5 | 37.3 | Hainan (China) | | | | | Data from Jan 22 to March 13, 2020 | Yan et al. 2020 | https://doi.org/10.1101/2020.03.19.20038539 |
| Onset of symptoms to death | 17 | | | | | | | | Days | 15 | | | 9 | 21 | Wuhan (China) | | 72 | | 76.5 | Patients admitted from Jan 3 to Feb 1, 2020. Outcomes follow up to Feb 15,2020 | Cao Jianlei et al. 2020 | https://doi.org/10.1093/cid/ciaa243 |
| Onset of symptoms to death | 82 | | | | | | | | Days | 15 | | | 11 | 20 | Wuhan (China) | | 72.5 | | 65.9 | Data from Jan 11 – Feb 10 | Zhang Bicheng et al. 2020 | https://doi.org/10.1371/journal.pone.0235458 |
| Onset of symptoms to death | 109 | 22.3 | | | | 9.2 | | | Days | | | | | | Wuhan (China) | 70 | | | 67.9 | Data up to February 24, 2020 | Du Rong-Hui et al. 2020 | https://doi.org/10.1513/AnnalsATS.202003-225OC |
| Onset of symptoms to death | 39 | 16 | | | | 8 | | | Days | | | | | | China | | | | | Gamma distribution Data up to Feb 5, 2020. | Mizumoto et al. 2020 | https://doi.org/10.1101/2020.02.19.20025163 https://doi.org/10.3201/eid2606.200233 |
| Onset of symptoms to death | 66 | | 1-24 | | | | | | Days | 10 | | | | | Republic of Korea | | | | | Data up to March 12, 2020 | Choe et al. 2020 | https://doi.org/ 10.24171/j.phrp.2020.11.2.05 |
| Onset of symptoms to death | 159 | | | | | | | | Days | 15 | | | 10 | 20 | Hubei + other provinces in mainland China (China) | | 71 | | 62.3 | Data of mortality from Jan 25 to March 20, 2020 | Guo et al. 2020 | https://doi.org/10.1101/2020.04.12.20062380 |
| Onset of symptoms to death | 113 | | | | | | | | Days | 16 | | | 12 | 20 | Wuhan (China) | | 68 | | 73 | Data from Jan 13 to Feb 28, 2020 | Chen T et al. 2020 | http://dx.doi.org/10.1136/bmj.m1091 |
| Onset of symptoms to death ** | 30 | 15.85 | | | | 7.17 | | | Days | | | | | | Wuhan (China) | 69.9 | | | 80 | Admissions until Jan 23. Deaths until Jan 28, 2020 | Wu Joseph et al. 2020 | https://doi.org/10.1038/s41591-020-0822-7 |
| Illness onset to death | 5 | | | | | | | | Days | 16 | | | 9 | 21 | Beijing (China) | | | | | Patients admitted from Jan 21 to February 8, 2020 Outcomes follow-up to Feb 29,2020 | Zhao Wen et al. 2020 | https://doi.org/10.15212/CVIA.2021.0019 |
| Illness onset to death | 34 | 15.1 | | 12.7 | 17.8 | | | | Days | 14.3 | 11.8 | 17.1 | | | Residents from other locations who travelled to Wuhan + Wuhan residents | | | | | Accounting without right truncation. Weibull Data up to Jan 31, 2020 | Linton et al. 2020 | https://doi.org/10.3390/jcm9020538 |
| Illness onset to death | 34 | 15 | | 12.8 | 17.5 | | | | Days | 13.9 | 11.8 | 17.1 | | | Residents from other locations who travelled to Wuhan + Wuhan residents | | | | | Accounting without right truncation. Gamma Data up to Jan 31, 2020 | Linton et al. 2020 | https://doi.org/10.3390/jcm9020538 |
| Illness onset to death | 34 | 14.5 | | 12.5 | 17 | | | | Days | 13.2 | 11.3 | 15.3 | | | Residents from other locations who travelled to Wuhan + Wuhan residents | | | | | Accounting without right truncation. Lognormal Data up to Jan 31, 2020 | Linton et al. 2020 | https://doi.org/10.3390/jcm9020538 |
| Illness onset to death | | 20.2 | | 15.1 | 29.5 | | | | Days | 17.1 | 13.5 | 24.1 | | | Residents from other locations who travelled to Wuhan + Wuhan residents | | | | | Right-truncated incubation. Lognormal Data up to Jan 31, 2020 | Linton et al. 2020 | https://doi.org/10.3390/jcm9020538 |
| Illness onset to death | 54 | | | | | | | | Days | 18.5 | | | 15 | 22 | Patients from Wuhan Hospitals | | | | | Data from Dec 29 to Jan 31, 2020 | Zhou Fei et al. 2020 | https://doi.org/10.1016/S0140-6736(20)30566-3 |
| Illness onset to death | 41 | 19.9 | | 14.9 | 29 | 11.4 | 6.5 | 21.6 | Days | | | | | | China | | | | | lognormal distribution and accounting for right truncation Cutoff Jan 24, 2020 | Jung et al. 2020 | https://doi.org/10.3390/jcm9020523 |

| | | | | | | | | | | | | | | | | | | | | | | |
|---|---|---|---|---|---|---|---|---|---|---|---|---|---|---|---|---|---|---|---|---|---|---|
| Illness onset to death | 26 | 22.3 | | 18 | 82 | 0.4 2 | 0.33 | 0.6 | Days | | | | | | Mainland China | | | | | Data up to February 10, 2020 | Dorigatti el al. 2020 | https://www.imperial.ac.uk/media/imperial-college/medicine/mrc-gida/2020-02-10-COVID19-Report-4.pdf |
| Illness onset to death | 23 | 16.1 | | 13.1 | 20.2 | | | | Days | | | | | | Wuhan (China) | 71.3 | | | 69.6 | | Sanche et al. 2020 | https://doi.org/10.3201/eid2607.200282 |
| **Onset of symptoms/illness onset to hospital discharge/recovery (days)** | | | | | | | | | | | | | | | | | | | | | | |
| Onset of symptoms to hospital discharge | 165 | 24.7 | | 22.9 | 28.1 | | | | Days | | | | | | Outside China (37 countries + Hong Kong +Macao) | | | | | Data up to Feb 25, 2020 | Verity et al. 2020 | https://doi.org/10.1016/S1473-3099(20)30243-7 |
| Onset of symptoms to hospital discharge | 160 | | | | | | | | Days | 22 | | | 17 | 17 | Hainan | | | | | Data from Jan to March 13, 2020 | Yan et al. 2020 | https://doi.org/10.1101/2020.03.19.20038539 |
| Onset of symptoms to hospital discharge | 161 | | | | | | | | Days | 26 | | | 21.8 | 29 | Wuhan (China) | | 51 | | 55 | Data from Jan 13 to Feb 28, 2020 | Chen T et al. 2020 | http://dx.doi.org/10.1136/bmj.m1091 |
| Illness onset to discharge | 137 | | | | | | | | Days | 22 | | | 18 | 25 | Patients from Wuhan Hospital (China) | | | | | Data from Dec 29 to Jan 31, 2020 | Zhou Fei et al. 2020 | https://doi.org/10.1016/S0140-6736(20)30566-3 |
| Illness onset to discharge | 64 | | | | | | | | Days | 18.5 | | | 15 | 22 | Beijing (China) | | | | | Patients admitted from Jan 21 to Feb 8. Outcomes follow-up to Feb 29, 2020 | Zhao Wen et al. 2020 | https://doi.org/10.15212/CVIA.2021.0019 |
| Onset of symptoms to recovery * | 228 | | | | | | | | Days | 20.8 | 20.1 | 21.5 | | | China | | | 0-70+ | | Data from Jan 14 to Feb 22, 2020 | Bi et al. 2020 | https://doi.org/10.1016/S1473-3099(20)30287-5 |
| Onset of symptoms to recovery | 29 | 22.2 | | 18 | 83 | 0.4 5 | 0.35 | 0.62 | Days | | | | | | Outside Mainland China | | | | | Data up to February 10, 2020 | Dorigatti el al. 2020 | https://www.imperial.ac.uk/media/imperial-college/medicine/mrc-gida/2020-02-10-COVID19-Report-4.pdf |
| **Presymptomatic transmission (%)** | | | | | | | | | | | | | | | | | | | | | | |
| % presymptomatic transmission | 468 | 12.6 | | | | | | | % | | | | | | China | | | | | Data from Jan 21 to Feb 8, 2020 | Du Zhanwei et al. 2020 | https://doi.org/10.3201/eid2606.200357 |
| % presymptomatic transmission | 157 | 6.4 | | | | | | | % | | | | | | Singapore | | | | | Data from Jan 23 to March 16, 2020 | Wei Wycliffe et al. 2020 | http://dx.doi.org/10.15585/mmwr.mm6914e1 |
| % presymptomatic transmission | 77 | 44 | | 30 | 57 | | | | % | | | | | | Guangzhou (China) | | | | | Data from Dec 18, 2019 to March 20 , 2020 | He Xi et al. 2020 | https://doi.org/10.1038/s41591-020-0869-5 |
| % presymptomatic transmission | 54 | 48 | | 32 | 67 | | | | % | | | | | | Singapore | | | | | Data from Jan 21 to Feb 6, 2020 | Ganyani et al. 2020 | https://www.eurosurveillance.org/content/10.2807/1560-7917.ES.2020.25.17.2000257 |
| % presymptomatic transmission | 114 | 62 | | 50 | 76 | | | | % | | | | | | Tianjin (China) | | | | | Data from Jan 14 to Feb 27,2020 | Ganyani et al. 2020 | https://www.eurosurveillance.org/content/10.2807/1560-7917.ES.2020.25.17.2000257 |
| **Presymptomatic transmission period (days)** | | | | | | | | | | | | | | | | | | | | | | - |
| Presymptomatic transmission | 10 | | 1 - 3 | | | | | | Days | | | | | | Singapore | | | | | Presymptomatic transmission exposure ocurred 1-3 days before the source patient developed symptoms Data from Jan 23 to March 16, 2020 | Wei Wycliffe et al. 2020 | http://dx.doi.org/10.15585/mmwr.mm6914e1 |

| | | | | | | | | | | | | | | | | | | | | | | |
|---|---|---|---|---|---|---|---|---|---|---|---|---|---|---|---|---|---|---|---|---|---|---|
| Presymptomatic transmission | 17 | | 1 - 2 | | | | | | Days | | | | | | Zhuhai (China) | | | | | “Viraemia might be high enough to trigger transmission for 1-2 days before onset of symptoms” January 2020 | Anderson et al. 2020 | https://doi.org/10.1016/S0140-6736(20)30567-5 |
| Presymptomatic transmission | | | 2 - 3 | | | | | | Days | | | | | | Guangzhou (China) | | 47 | | 50 | Data from Dec 18, 2019 to March 5,2020 | He Xi et al. 2020 | https://doi.org/10.1038/s41591-020-0869-5 |
| Presymptomatic transmission | 93 | 1.99 | | | | | | | Days | | | | | | Singapore | | | | | Data from Jan 18 to Feb 28,2020 | Tindale et al. 2020 | https://doi.org/10.7554/eLife.57149 |
| Presymptomatic transmission | 135 | 3.68 | | | | | | | Days | | | | | | Tianjin (China) | | | | | Data from Jan 5 to Feb 22, 2020 | Tindale et al. 2020 | https://doi.org/10.7554/eLife.57149 |
| **Asymptomatic patients (%)** | | | | | | | | | | | | | | | | | | | | | | - |
| Asymptomatic patients | 23 | 13.04 | | | | | | | % | | | | | | United States | | | | | Data from Feb 28 to March 13, 2020 | Kimball et al. 2020 | http://dx.doi.org/10.15585/mmwr.mm6913e1 |
| Asymptomatic patients | 328 | 3.05 | | | | | | | % | | | | | | Shangai | | | | | Data up to March 4, 2020 | Zhou X et al. 2020 | https://doi.org/10.1016/j.cmi.2020.03.024 |
| Asymptomatic patients * | 167 | 2 | | | | | | | % | | | | | | Chongqing (China) | | 46 | | | Data from Jan to March, 2020 | Tao et al. 2020 | https://doi.org/10.1101/2020.03.16.20037259 |
| Asymptomatic patients | 262 | 5 | | | | | | | % | | | | | | Beijing (China) | | | | | Data from Jan 20 to Feb 10, 2020 | Tian Sijia et al .2020 | https://doi.org/10.1016/j.jinf.2020.02.018 |
| Asymptomatic patients | 83 | 21.7 | | | | | | | % | | | | | | Tibet | | 31 | | 61.11 | Data from Jan 26 – to Feb 28 | Song et al. 2020 | https://doi.org/10.1101/2020.03.27.20043836 |
| Asymptomatic patients | 78 | 2.6 | | | | | | | % | | | | | | China | | | | | January 2020 | Mao et al. 2020 | https://doi.org/10.1016/j.ijid.2020.03.041 |
| Asymptomatic patients | 391 | 6.4 | | | | | | | % | | | | | | Shenzen (China) | | | | 47.8 | Data from Jan 14 to Feb 12, 2020 | Bi et al. 2020 | https://doi.org/10.1016/S1473-3099(20)30287-5 |
| Asymptomatic patients | 1012 | 1.4 | | | | | | | % | | | | | | Wuhan (China) | | 50 | | 51.8 | Patients admitted between Feb 7 and Feb 12, 2020 Clinical course recorded up to Feb 22, 2020 | Wang X et al. 2020 | https://doi.org/10.1016/j.cmi.2020.03.032 |
| Asymptomatic patients | 295 | 1.4 | | | | | | | % | | | | | | Guangzhou (China) | | | | | Data from Jan 23 to Feb 18, 2020 | Ling et al. 2020 | https://doi.org/10.1016/j.ejrad.2020.108956 |
| Asymptomatic patients | 6 | 16.7 | | | | | | | % | | | | | | Shenzen (China) | | | | | Data from January 2020 | Chan et al. 2020 | https://doi.org/10.1016/S0140-6736(20)30154-9 |
| Asymptomatic patients | 24 | 79.1 | | | | | | | % | | | | | | Nanjing (China) | | 32.5 | | | Data from Jan 28 to Feb 9 | Hu et al. 2020 | https://doi.org/10.1007/s11427-020-1661-4 |
| Asymptomatic patients | 3 | 66.7 | | | | | | | % | | | | | | Wuhan (China) | | 33 | | | Data from January 2020 | Pan et al. 2020 | https://doi.org/10.1016/S1473-3099(20)30114-6 |
| Asymptomatic patients | 634 | 17.9 | | 15.5 | 20.2 | | | | % | | | | | | Diamond Princess cruise ship | | | | 50.3 | Bayesian framework using hamiltonian Monte Carlo (HMC) algorithm. | Mizumoto et al. 2020 | https://doi.org/10.2807/1560-7917.ES.2020.25.10.2000180 |
| Asymptomatic patients | 565 | 30.8 | | 7.7 | 53.8 | | | | % | | | | | | Japanese evacuated from Wuhan (China) | | | | | binomial distribution | Nishiura et al. 2020 | https://doi.org/10.1016/j.ijid.2020.03.020 |
| Asymptomatic patients | 50 | 4 | | | | | | | % | | | | | | Hong Kong | 55.2 | | | 46 | Data from Jan 26 to Feb 28 | Leung et al. 2020 | https://doi.org/10.3201/eid2701.201543 |
| Asymptomatic patients | 24 | 29.17 | | | | | | | % | | | | | | Liaocheng city (China) | 48 | | | 37.5 | Period unknown, but at least one admission after Jan 9, 2020 | Tian Suochen et al .2020 | https://doi.org/10.1186/s12889-021-10713-z |
| Asymptomatic patients | 11 | 9.1 | | | | | | | % | | | | | | Thailand | | 61 | | 54.5 | Patients admitted from Jan 8-31, 2020 | Pongpirul et al. 2020 | https://doi.org/10.3201/eid2607.200598 |

| | | | | | | | | | | | | | | | | | | | | | | |
|---|---|---|---|---|---|---|---|---|---|---|---|---|---|---|---|---|---|---|---|---|---|---|
| Asymptomatic patients | 18 | 5.6 | | | | | | | % | | | | | | Guandong (China) | | 59 | | 50 | Data from Jan 7-26, 2020 | Zou et al. 2020 | https://doi.org/10.1056/NEJMc2001737 |
| Asymptomatic patients | 12 | 8.3 | | | | | | | % | | | | | | Vietnam | | 31.2 | | 25 | Data from Jan – February, 2020 | Le et al. 2020 | https://doi.org/10.3201/eid2607.200591 |
| Asymptomatic patients | 206 | 5.8 | | 3.4 | 9.9 | | | | % | | | | | | Wenzhou (China) | | 31 | | 50 | Period unknow | He Guiing et al. 2020 | https://doi.org/10.1002/jmv.25861 |
| Asymptomatic patients | 8 | 25 | | | | | | | % | | | | | | China | 46.6 | | | 37.5 | Data from Jan 19 to Feb 11, 2020 | Qian et al. 2020b | https://doi.org/10.1093/cid/ciaa316 |
| Asymptomatic patients | 46 | 4.3 | | | | | | | % | | | | | | Chongqing (China) | | | 10-35 | 52.2 | Data from Jan 25 to Feb 18, 2020. Outcomes until Feb 23, 2020 | Liao et al. 2020 | https://doi.org/10.1016/j.xinn.2020.04.001 |
| **ICU admissions vs all hospitalized patients with COVID-19 (%)** | | | | | | | | | | | | | | | | | | | | | | |
| ICU admissions vs all hospitalized patients with COVID-19 * | 1099 | 5 | | | | | | | % | | | | | | Mainland China | | 47 | | 58.1 | Data up to Jan 29, 2020 | Guan et al. 2020 | https://doi.org/10.1056/NEJMoa2002032 |
| ICU admissions vs all hospitalized patients with COVID-19 | 2217 | 16 | | | | | | | % | | | | | | Italy | | | | | | Grasselli et al. 2020 | https://doi.org/10.1001/jama.2020.4031 |
| ICU admissions vs all hospitalized patients with COVID-19 | 41 | 32 | | | | | | | % | | | | | | Patients from Wuhan Hospital (China) | | 49 | | 73 | Data from Dec 16 to Jan 2,2020 | Huang Chaolin et al. 2020 | https://doi.org/10.1016/S0140-6736(20)30183-5 |
| ICU admissions vs all hospitalized patients with COVID-19 * | 191 | 26 | | | | | | | % | | | | | | Patients from Wuhan Hospital (China) | | 51 | | 62 | Data from Dec 29 to Jan 31, 2020 | Zhou Fei et al. 2020 | https://doi.org/10.1016/S0140-6736(20)30566-3 |
| ICU admissions vs all hospitalized patients With COVID 19 | 201 | 26.4 | | | | | | | % | | | | | | Patients from Wuhan Hospital (China) | | 51 | | 63.7 | Data from Dec 25, 2019 to Jan 26, 2020 | Wu Chaomin et al. 2020 | https://doi.org/10.1001/jamainternmed.2020.0994 |
| ICU admissions vs all hospitalized patients with COVID-19 | 138 | 26.1 | | | | | | | % | | | | | | Wuhan (China) | | 56 | | 61.1 | Data from Jan 1 to Jan 28, 2020 | Wang Dawei et al. 2020 | https://doi.org/10.1001/jama.2020.1585 |
| ICU admissions vs all hospitalized patients with COVID-19 * | 267 | 19.9 | | | | | | | % | | | | | | Chongqing (China) | | 48 | | 55.8 | Data from Jan 19 to Feb 16, 2020 | Qi Di et al. 2020 | https://doi.org/10.1101/2020.03.01.20029397 |
| ICU admissions vs all hospitalized patients with COVID-19 | 62 | 2 | | | | | | | % | | | | | | Zhejiang (China) | | 41 | | 56 | Data from Jan 10 to Jan 26, 2020 | Xu Xiao-Wei et al. 2020 | http://dx.doi.org/10.1136/bmj.m606 |
| ICU admissions vs all hospitalized patients with COVID-19 | 100 | 15 | | | | | | | % | | | | | | Singapore | 42.5 | 41 | | 60 | Data from Jan 2 to Feb 29, 2020 | Ng el al. 2020 | http://dx.doi.org/10.15585/mmwr.mm6911e1 |
| ICU admissions vs all hospitalized patients with COVID-19 | 652 | 1.38 | | | | | | | % | | | | | | Zhejiang (China) | 41.15 | | <60 | 53.53 | Data from Jan 17 to Feb 12, 2020 | Lian et al. 2020 | https://doi.org/10.1093/cid/ciaa242 |
| ICU admissions vs all hospitalized patients with COVID-19 | 136 | 9.56 | | | | | | | % | | | | | | Zhejiang (China) | 68.27 | | >60 | 42.65 | Data from Jan 17 to Feb 12, 2020 | Lian et al. 2020 | https://doi.org/10.1093/cid/ciaa242 |
| ICU admissions vs all hospitalized patients with COVID-19 | 49 | 16 | | | | | | | % | | | | | | Hong Kong | | 64.5 | | | Patients admitted from Jan 22 to Feb 11, 2020 | Ling Lowell et al. 2020 | https://www.ncbi.nlm.nih.gov/pubmed/32248675 |
| ICU admissions vs all hospitalized patients with COVID-19 | | | 9-11.0 | | | | | | % | | | | | | Italy | | | | | reported daily in Italy from March 1 until March 11 | Remuzzi et al. 2020 | https://doi.org/10.1016/S0140-6736(20)30627-9 |
| ICU admissions vs all hospitalized patients with COVID-19 | 99 | 23 | | | | | | | % | | | | | | Wuhan (China) | 55.5 | | | 68 | Data from Jan 1 to Jan 20, 2020 | Chen N et al. 2020 | https://doi.org/10.1016/S0140-6736(20)30211-7 |
| ICU admissions vs all hospitalized patients with COVID-19 | 197 | 29.4 | | | | | | | % | | | | | | Hubei (China) | 55.94 | | | 50.3 | Patients discharged form Jan 17 to Feb 26, 2020 | Zhou Fating et al. 2020 | https://doi.org/10.1101/2020.03.26.20041426 |
| ICU admissions vs all hospitalized patients with COVID-19 * | 102 | 17.6 | | | | | | | % | | | | | | Wuhan (China) | | 54 | | 52 | Data from Jan 3 and Feb 1, 2020 | Cao Jianlei et al. 2020 | https://doi.org/10.1093/cid/ciaa243 |

| | | | | | | | | | | | | | | | | | | | | | | |
|---|---|---|---|---|---|---|---|---|---|---|---|---|---|---|---|---|---|---|---|---|---|---|
| ICU admissions vs all hospitalized patients with COVID-19 | 50 | 6 | | | | | | | % | | | | | | Hong Kong | 55.2 | | | 46 | Data from Jan 26 to Feb 28, 2020 | Leung et al. 2020 | https://doi.org/10.3201/eid2701.201543 |
| ICU admissions vs all hospitalized patients with COVID-19 | 297 | 41.1 | | | | | | | % | | | | | | USA | | 67.6 | | 97.3 | USA veterans | Rentsch et al. 2020 | https://doi.org/10.1101/2020.04.09.20059964 |
| ICU admissions vs all hospitalized patients with COVID-19 | 2669 | 16.5 | | | | | | | % | | | | | | Lombardia (Italy) | | | | | | Cereda et al. 2020 | https://arxiv.org/abs/2003.09320 |
| ICU admissions vs all hospitalized patients with COVID-19 | 9719 | 9.1 | | | | | | | % | | | | | | Lombardia. Veneto. Emilia-Romagna (Italy) | | | | | | Riccardo et al. 2020 | https://doi.org/10.1101/2020.04.08.20056861 |
| ICU admissions vs all hospitalized patients with COVID-19 * | 1590 | 6.23 | | | | | | | % | | | | | | Hubei + outside Hubei (China) | 48.9 | | | 57.3 | Cata cut-off Jan 31, 2020 | Liang et al. 2020 | https://doi.org/10.1183/13993003.00562-2020 |
| ICU admissions vs all hospitalized patients with COVID-19 | 125 | 15.2 | | | | | | | % | | | | | | Anhui (China) | 38.76 | | | 56.8 | Data from Jan 20 to Feb 9,2020 | Wang R et al. 2020 | https://doi.org/10.1016/j.ijid.2020.03.070 |
| ICU admissions vs all hospitalized patients with COVID-19 | 20 | 5 | | | | | | | % | | | | | | Guangzhou (China) | 43.2 | | | 50 | Patients admitted from Jan 22 to Feb 12, 2020. Outcomes follow-up Feb 18, 2020 | Lei et al. 2020 | https://doi.org/10.1016/j.tmaid.2020.101664 |
| ICU admissions vs all hospitalized patients with COVID-19 | 91 | 9.9 | | | | | | | % | | | | | | Zhejiang province (China) | | 50 | | 40.7 | Data colected from Jan 20 to Feb 11, 2020. Final folow-up until Feb 16, 2020 | Qian et al. 2020 | https://doi.org/10.1093/qjmed/hcaa089 |
| ICU admissions vs all hospitalized patients with COVID-19 | 18 | 11.1 | | | | | | | % | | | | | | Singapore | | 47 | | 50 | Data from Jan 23 to Feb 3, 2020. Final follow-up was Feb 25, 2020 | Young et al. 2020 | https://doi.org/10.1001/jama.2020.204 |
| ICU admissions vs all hospitalized patients with COVID-19 | 18 | 5.6 | | | | | | | % | | | | | | Zhengzou (China) | | 39 | | 55.6 | Data from Jan 21 to Feb 5. Final follo-up was Feb 7, 2020 | Wang Lei et al. 2020 | https://doi.org/10.1183/13993003.00398-2020 |
| ICU admissions vs all hospitalized patients with COVID-19 | 221 | 19,9 | | | | | | | % | | | | | | Wuhan (China) | | 55 | | 48.9 | Data from Jan 2 to Feb 10, 2020 | Zhang Guqin et al. 2020 | https://doi.org/10.1101/2020.03.02.20030452 https://doi.org/10.1016/j.jcv.2020.104364 |
| ICU admissions vs all hospitalized patients with COVID-19 | 82 | 17,1 | | | | | | | % | | | | | | Wuhan (China) | | 72.5 | | 65.9 | Data from Jan 11 to Feb 10, 2020 | Zhang Bicheng et al. 2020 | https://doi.org/10.1101/2020.02.26.20028191 https://doi.org/10.1371/journal.pone.0235458 |
| **Transferred from ICU to general wards (%)** | | | | | | | | | | | | | | | | | | | | | | |
| Transferred from ICU to general wards | 36 | 27.8 | | | | | | | % | | | | | | Wuhan (China) | | 66 | | 61.1 | Data from Jan 1 to Feb 3, 2020 | Wang Dawei et al. 2020 | https://doi.org/10.1001/jama.2020.1585 |
| Transferred from ICU to general wards | 24 | 16.6 | | | | | | | % | | | | | | Seattle region | | | | | | Bhatraju et al. 2020 | https://doi.org/10.1056/NEJMoa2004500 |
| **Death vs all hospitalized patients with COVID 19 (%)** | | | | | | | | | | | | | | | | | | | | | | |
| Death vs all hospitalized patients with COVID 19 | 191 | 28.3 | | | | | | | % | | | | | | Wuhan (China) | | | | | Data from Dec 29 to Jan 31, 2020 | Zhou Fei et al. 2020 | https://doi.org/10.1016/S0140-6736(20)30566-3 |
| Death vs all hospitalized patients with COVID 19 | 201 | 21.9 | | | | | | | % | | | | | | Wuhan (China) | | 51 | | 63.7 | Patients admitted from Dec 25, 2019 to Jan 26, 2020. Outcome follow-up to Feb 13, 2020 | Wu Chaomin et al. 2020 | https://doi.org/10.1001/jamainternmed.2020.0994 |
| Death vs all hospitalized patients with COVID 19 | 138 | 4.3 | | | | | | | % | | | | | | Wuhan (China) | | 56 | | 54.3 | Data from Jan 1 to Feb 3, 2020 | Wang Dawei et al. 2020 | https://doi.org/10.1001/jama.2020.1585 |

| | | | | | | | | | | | | | | | | | | | | | | |
|---|---|---|---|---|---|---|---|---|---|---|---|---|---|---|---|---|---|---|---|---|---|---|
| Death vs all hospitalized patients with COVID 19 | 41 | 15 | | | | | | | % | | | | | | Patients from Wuhan Hospital (China) | | 49 | | 73 | Data from Dec 16, 2019 to Jan 22, 2020 | Huang Chaolin et al. 2020 | https://doi.org/10.1016/S0140-6736(20)30183-5 |
| Death vs all hospitalized patients with COVID 19 | 73 | 31.8 | | | | | | | % | | | | | | Wuhan (China) | | 67 | | 61.1 | Data from December 24, 2019 to February 7,2020 | Tang et al. 2020 | https://doi.org/10.1016/j.chest.2020.03.032 |
| Death vs all hospitalized patients with COVID 19 | 67 | 7.5 | | | | | | | % | | | | | | Wuhan (China) | | 42 | | 46 | Patients hospitalized from Jan 16 to Jan 29, 2020. Outcome follow-up Feb 4,2020 | Wang Zhongliang et al. 2020 | https://doi.org/10.1093/cid/ciaa272 |
| Death vs all hospitalized patients with COVID 19 | 199 | 22.1 | | | | | | | % | | | | | | Wuhan (China) | | 58 | | 60.3 | Data from Jan 18 to Feb 3, 2020 | Cao Bin et al. 2020 | https://doi.org/10.1056/NEJMoa2001282 |
| Death vs all hospitalized patients with COVID 19 | 62 | 0 | | | | | | | % | | | | | | Zhejiang (China) | | 41 | | 56 | Data from Jan 10 to Jan 26, 020 | Xu Xiao-Wei et al. 2020 | http://dx.doi.org/10.1136/bmj.m606 |
| Death vs all hospitalized patients with COVID 19 | 99 | 11 | | | | | | | % | | | | | | Wuhan (China) | 55.5 | | | 68 | Data from Jan 1 to Jan 25, 2020 | Chen N et al. 2020 | https://doi.org/10.1016/S0140-6736(20)30211-7 |
| Death vs all hospitalized patients with COVID 19 | 168 | 3.6 | | | | | | | % | | | | | | Hainan (China) | | | | | Data from Jan 22 to March 13, 2020 | Yan et al. 2020 | https://doi.org/10.1101/2020.03.19.20038539 |
| Death vs all hospitalized patients with COVID 19 | 77 | 6.5 | | | | | | | % | | | | | | Beijing (China) | | | | | Patients admitted from Jan 21 to February 8, 2020 Outcomes follow-up to Feb 29,2020 | Zhao Wen et al. 2020 | https://doi.org/10.1101/2020.03.13.20035436 |
| Death vs all hospitalized patients with COVID 19 | 102 | 16.7 | | 9.4 | 23.9 | | | | % | | | | | | Wuhan (China) | | 72 | | | Patients admitted from Jan 3 to Feb 1, 2020. Outcomes follow up to Feb 15,2020 | Cao Jianlei et al. 2020 | https://doi.org/10.1093/cid/ciaa243 |
| Death vs all hospitalized patients with COVID 19 * | 221 | 5.4 | | | | | | | % | | | | | | Wuhan (China) | | 55 | | 48.9 | Data from Jan 2 to Feb 10, 2020 Outcomes follow up to Feb 15, 2020 | Zhang Guqin et al. 2020 | https://doi.org/10.1016/j.jcv.2020.104364 |
| Death vs all hospitalized patients with COVID 19 | 197 | 14.2 | | | | | | | % | | | | | | Hubei (China) | | | | | Patients discharged form Jan 17 to Feb 26, 2020 | Zhou Fating et al. 2020 | https://doi.org/10.1101/2020.03.26.20041426 |
| Death vs all hospitalized patients with COVID 19 | 24 | 50 | | | | | | | % | | | | | | Seattle region (USA) | | | | | | Bhatraju et al. 2020 | https://doi.org/10.1056/NEJMoa2004500 |
| Death vs all hospitalized patients with COVID 19 * | 267 | 1.5 | | | | | | | % | | | | | | Chongqing (China) | | 48 | | 44.2 | Data from Jan 19 to Feb 16, 2020 | Qi Di et al. 2020 | https://doi.org/10.1101/2020.03.01.20029397 |
| Death vs all hospitalized patients with COVID 19 * | 291 | 0.7 | | | | | | | % | | | | | | Hunan (China) | | 46 | | 49.8 | Data from Jan 23 to Feb 14, 2020. Outcomes until Feb 20, 2020 | Chen X et al. 2020 | https://doi.org/10.1101/2020.03.03.20030353 |
| Death vs all hospitalized patients with COVID 19 * | 1099 | 1.4 | | | | | | | % | | | | | | Mainland China | | 47 | | 58.1 | Data up to Jan 29, 2020 | Guan et al. 2020 | https://doi.org/10.1056/NEJMoa2002032 |
| Death vs all hospitalized patients with COVID 19 | 538 | 0.5 | | | | | | | % | | | | | | Sichuan (China) | | | | 53 | Data from January to March, 2020 | Wang Zhuo et al. 2020 | https://doi.org/10.1101/2020.04.07.20057299 |
| Death vs all hospitalized patients with COVID 19 | 1999 | 14.6 | | | | | | | % | | | | | | New York city (USA) | | 62 | | 62.6 | | Petrilli et al. 2020 | https://doi.org/10.1136/bmj.m1966 |
| Death vs all hospitalized patients with COVID 19 | 297 | 5.7 | | | | | | | % | | | | | | USA | | 67.6 | | 97.3 | USA veterans | Rentsch et al. 2020 | https://doi.org/10.1101/2020.04.09.20059964 |
| Death vs all hospitalized patients with COVID 19 | 799 | 14.1 | | | | | | | % | | | | | | Wuhan (China) | | | | | Data from Jan 13 to Feb 28, 2020 | Chen et al. 2020 | http://dx.doi.org/10.1136/bmj.m1091 |

| | | | | | | | | | | | | | | | | | | | | | | |
|---|---|---|---|---|---|---|---|---|---|---|---|---|---|---|---|---|---|---|---|---|---|---|
| Death vs all hospitalized patients with COVID 19 * | 1590 | 3.14 | | | | | | | % | | | | | | Hubei + outside Hubei (China) | 48.9 | | | | Cata cut-off Jan 31, 2020 | Liang et al. 2020 | https://doi.org/10.1183/13993003.00562-2020 |
| Death vs all hospitalized patients with COVID 19 | 137 | 11.7 | | | | | | | % | | | | | | Hubei (China) | | 57 | | 44.5 | Patients admitted from Dec 30, 2019 to Jan 24, 2020 | Liu et al. 2020 | https://doi.org/10.1097/CM9.0000000000000744 |
| Death vs all hospitalized patients with COVID 19 | 203 | 12.8 | | | | | | | % | | | | | | Wuhan (China) | | 54 | | 53.2 | Data from Jan 1 to Feb 20, 2020 | Chen TL et al. 2020 | https://doi.org/10.1093/gerona/glaa089 |
| Death vs all hospitalized patients with COVID 19 * | 262 | 0.9 | | | | | | | % | | | | | | Beijing (China) | | 47.5 | | 48.5 | Data from Jan 20 to Feb 10, 2020 (outcomes included) | Tian Sijia et al. 2020 | https://doi.org/10.1016/j.jinf.2020.02.018 |
| Death vs all hospitalized patients with COVID 19 * | 417 | 0.7 | | | | | | | % | | | | | | Shenzhen (China) | 45.4 | | | 47.2 | Cases from Jan 1 Feb 28, 2020 | Wen et al. 2020 | https://doi.org/10.1101/2020.03.22.20035246 |
| Death vs all hospitalized patients with COVID 19 * | 91 | 0 | | | | | | | % | | | | | | Zhejiang province (China) | | 50 | | 40.7 | Data colected from Jan 20 to Feb 11, 2020. Final folow-up until Feb 16, 2020 | Qian et al. 2020 | https://doi.org/10.1093/qjmed/hcaa089 |
| Death vs all hospitalized patients with COVID 19 * | 985 | 11 | | | | | | | % | | | | | | Wuhan (China) | | | | | Data from Jan 30 to Feb 25, 2020 (outcomes included) | Luo et al. 2020 | https://doi.org/10.1101/2020.03.19.20033175 |
| Death vs all hospitalized patients with COVID 19 | 135 | 2.2 | | | | | | | % | | | | | | Tianjin (China) | | | | | Data from Jan 21 to Feb 22, 2020 | Tindale et al. 2020 | https://doi.org/10.7554/eLife.57149 |
| **Deaths from ICU (%)** | | | | | | | | | | | | | | | | | | | | | | |
| Deaths from ICU | 36 | 16.7 | | | | | | | % | | | | | | Wuhan (China) | | 66 | | 61.1 | Data from Jan 1 to Feb 3, 2020 | Wang Dawei et al. 2020 | https://doi.org/10.1001/jama.2020.1585 |
| Deaths from ICU | 13 | 38 | | | | | | | % | | | | | | Patients from Wuhan Hospital (China) | | | | | Data from Dec 16, 2019 to Jan 22, 2020 | Huang Chaolin et al. 2020 | https://doi.org/10.1016/S0140-6736(20)30183-5/ |
| Deaths from ICU | 52 | 61.5 | | | | | | | % | | | | | | Wuhan (China) | | | | | Data from late Dec, 2019 to Feb 9, 2020 | Yang et al. 2020 | https://doi.org/10.1016/S2213-2600(20)30079-5 |
| Deaths from ICU | 21 | 67 | | | | | | | % | | | | | | Washington State | 70 | | | 52 | Patients admitted between Feb 20 and March 5, 2020. Outcomes by March 17, 2020 | Arentz et al. 2020 | https://doi.org/10.1001/jama.2020.4326 |
| Deaths from ICU | 8 | 12.5 | | | | | | | % | | | | | | Hong Kong | | 64.5 | | 50 | Patients admitted from Jan 22 to Feb 11, 2020 | Ling Lowell et al. 2020 | https://www.ncbi.nlm.nih.gov/pubmed/32248675 |
| Deaths from ICU | 344 | 38.7 | | | | | | | % | | | | | | Wuhan (China) | | 70 | | 55.6 | Patients enrolled from Jan 25 to Feb 25, 2020 | Wang Yang et al. 2020 | https://doi.org/10.1164/rccm.202003-0736LE |
| Deaths from ICU | 48 | 15 | | | | | | | % | | | | | | Vitoria (Spain) | | 63 | | 56.3 | Patients from March 4 to March 24, 2020 | Barrasa et al. 2020 | https://doi.org/doi:10.1016/j.accpm.2020.04.001 |
| Deaths from ICU | 33 | 3 | | | | | | | % | | | | | | Brescia (Italy) | | 64 | | 91 | Patients admitted between March 2 and March 13, 2020 | Piva et al. 2020 | https://doi.org/10.1016/j.jcrc.2020.04.004 |
| **Discharged from all hospitalized patients with COVID 19 (%)** | | | | | | | | | | | | | | | | | | | | | | |
| Discharged from all hospitalized patients with COVID 19 | 41 | 68 | | | | | | | % | | | | | | Patients from Wuhan Hospital (China) | | 49 | | 73 | Data from Dec 16, 2019 to Jan 22, 2020 | Huang Chaolin et al. 2020 | https://doi.org/10.1016/S0140-6736(20)30183-5/ |
| Discharged from all hospitalized patients with COVID 19 | 138 | 34.1 | | | | | | | % | | | | | | Wuhan (China) | | 56 | | 54.3 | Hospitalized January 1-28. Discharged on February 3 | Wang Dawei et al. 2020 | https://doi.org/10.1001/jama.2020.1585 |

| | | | | | | | | | | | | | | | | | | | | | | |
|---|---|---|---|---|---|---|---|---|---|---|---|---|---|---|---|---|---|---|---|---|---|---|
| Discharged from all hospitalized patients with COVID 19 | 191 | 71.7 | | | | | | | % | | | | | | Wuhan (China) | | | | | Data from Dec 29 to Jan 31, 2020 | Zhou Fei et al. 2020 | https://doi.org/10.1016/S0140-6736(20)30566-3 |
| Discharged from all hospitalized patients with COVID 19 | 73 | 35.6 | | | | | | | % | | | | | | Wuhan (China) | | 67 | | 61.6 | Data from December 24, 2019 to February 7,2020 | Tang et al. 2020 | https://doi.org/10.1016/j.chest.2020.03.032 |
| Discharged from all hospitalized patients with COVID 19 | 77 | 83.1 | | | | | | | % | | | | | | Beijing (China) | | | | | Patients admitted from Jan 21 to February 8, 2020 Outcomes follow-up to Feb 29,2020 | Zhao Wen et al. 2020 | https://doi.org/10.1101/2020.03.13.20035436 |
| Discharged from all hospitalized patients with COVID 19 | 102 | 83.3 | | | | | | | % | | | | | | Wuhan (China) | | 72 | | | Patients admitted from Jan 3 to Feb 1, 2020. Outcomes follow up to Feb 15,2020 | Cao Jianlei et al. 2020 | https://doi.org/10.1093/cid/ciaa243 |
| Discharged from all hospitalized patients with COVID 19 | 221 | 19 | | | | | | | % | | | | | | Wuhan (China) | | 55 | | 48.9 | Data from Jan 2 to Feb 10, 2020 Outcomes follow up to Feb 15, 2020 | Zhang Guqin et al. 2020 | https://doi.org/10.1016/j.jcv.2020.104364 |
| Discharged from all hospitalized patients with COVID 19 | 197 | 85.2 | | | | | | | % | | | | | | Hubei (China) | | | | | Patients discharged form Jan 17 to Feb 26, 2020 | Zhou Fating et al. 2020 | https://doi.org/10.1101/2020.03.26.20041426 |
| Discharged from all hospitalized patients with COVID 19 | 80 | 23.75 | | | | | | | % | | | | | | Jiangsu (China) | 46.1 | | | 48.75 | Patients enrolled from Jan 22 to Feb 14, 2020 | Wu Jian et al. 2020 | https://doi.org/10.1093/cid/ciaa199 |
| Discharged from all hospitalized patients with COVID 19 | 652 | 44.6 | | | | | | | % | | | | | | Zhejiang (China) | 41.15 | | <60 | 53.53 | Data from Jan 17 to Feb 12, 2020 (including outcomes) | Lian et al. 2020 | https://doi.org/10.1093/cid/ciaa242 |
| Discharged from all hospitalized patients with COVID 19 | 136 | 22.8 | | | | | | | % | | | | | | Zhejiang (China) | 68.27 | | >60 | 42.65 | Data from Jan 17 to Feb 12, 2020 (including outcomes) | Lian et al. 2020 | https://doi.org/10.1093/cid/ciaa242 |
| Discharged from all hospitalized patients with COVID 19 | 24 | 21 | | | | | | | % | | | | | | Seattle region | | | | | | Bhatraju et al. 2020 | https://doi.org/10.1056/NEJMoa2004500 |
| Discharged from all hospitalized patients with COVID 19 * | 291 | 54.6 | | | | | | | % | | | | | | Hunan (China) | | 46 | | 49.8 | Data from Jan 23 to Feb 14, 2020. Outcomes until Feb 20, 2020 | Chen X et al. 2020 | https://doi.org/10.1101/2020.03.03.20030353 |
| Discharged from all hospitalized patients with COVID 19 | 168 | 95.2 | | | | | | | % | | | | | | Hainan (China) | | | | | Data from Jan 22 to March 13, 2020 | Yan et al. 2020 | https://doi.org/10.1101/2020.03.19.20038539 |
| Discharged from all hospitalized patients with COVID 19 * | 267 | 38.6 | | | | | | | % | | | | | | Chongqing (China) | | 48 | | 55.8 | Data from Jan 19 to Feb 16, 2020 | Qi Di et al. 2020 | https://doi.org/10.1101/2020.03.01.20029397 |
| Discharged from all hospitalized patients with COVID 19 * | 1099 | 5.8 | | | | | | | % | | | | | | Mainland China | | 47 | | 58.1 | Data up to Jan 29, 2020 | Guan et al. 2020 | https://doi.org/10.1056/NEJMoa2002032 |
| Discharged from all hospitalized patients with COVID 19 | 99 | 31 | | | | | | | % | | | | | | Wuhan (China) | 55.5 | | | 68 | Data from Jan 1 to Jan 25, 2020 | Chen N et al. 2020 | https://doi.org/10.1016/S0140-6736(20)30211-7 |
| Discharged from all hospitalized patients with COVID 19 | 50 | 40 | | | | | | | % | | | | | | Hong Kong | 55.2 | | | 46 | Data from Jan 26 to Feb 28, 2020 | Leung et al. 2020 | https://doi.org/10.3201/eid2701.201543 |
| Discharged from all hospitalized patients with COVID 19 | 538 | 67.6 | | | | | | | % | | | | | | Sichuan (China) | | | | 53 | Data from January to March, 2020 | Wang Zhuo et al. 2020 | https://doi.org/10.1101/2020.04.07.20057299 |
| Discharged from all hospitalized patients with COVID 19 | 1999 | 49.1 | | | | | | | % | | | | | | New York City (USA) | | 62 | | 62.6 | | Petrilli et al. 2020 | https://doi.org/10.1101/2020.04.08.20057794 https://doi.org/10.1136/bmj.m1966 |
| Discharged from all hospitalized patients with COVID 19 | 799 | 20.2 | | | | | | | % | | | | | | Wuhan (China) | | | | | Data from Jan 13 to Feb 28, 2020 | Chen T et al. 2020 | http://dx.doi.org/10.1136/bmj.m1091 |
| Discharged from all hospitalized patients with COVID 19 | 125 | 37.6 | | | | | | | % | | | | | | Anhui (China) | 38.76 | | | 56.8 | Data from Jan 20 to Feb 18,2020 | Wang R et al. 2020 | https://doi.org/10.1016/j.ijid.2020.03.070 |

| | | | | | | | | | | | | | | | | | | | | | | |
|---|---|---|---|---|---|---|---|---|---|---|---|---|---|---|---|---|---|---|---|---|---|---|
| Discharged from all hospitalized patients with COVID 19 | 137 | 32.1 | | | | | | | % | | | | | | Hubei (China) | | 57 | | 44.5 | Patients admitted from Dec 30, 2019 to Jan 24, 2020 | Liu Kiu et al. 2020 | https://doi.org/10.1097/CM9.0000000000000744 |
| Discharged from all hospitalized patients with COVID 19 | 20 | 70 | | | | | | | % | | | | | | Guangzhou (China) | 43.2 | | | 50 | Patients admitted from Jan 22 to Feb 12, 2020. Outcomes follow-up Feb 18, 2020 | Lei et al. 2020 | https://doi.org/10.1016/j.tmaid.2020.101664 |
| Discharged from all hospitalized patients with COVID 19 | 4 | 50 | | | | | | | % | | | | | | Shangai (China) | 44.25 | | | 75 | Data from Jan 21 to Feb 4, 2020 | Wang Zhenwei et al. 2020 | https://doi.org/10.5582/bst.2020.01030 |
| Discharged from all hospitalized patients with COVID 19 | 203 | 87.2 | | | | | | | % | | | | | | Wuhan (China) | | 54 | | 53.2 | Data from Jan 1 to Feb 20, 2020 | Chen TL et al. 2020 | https://doi.org/10.1093/gerona/glaa089 |
| Discharged from all hospitalized patients with COVID 19 * | 262 | 17.2 | | | | | | | % | | | | | | Beijing (China) | | 47.5 | | 48.5 | Data from Jan 20 to Feb 10, 2020 | Tian Sijia et al. 2020 | https://doi.org/10.1016/j.jinf.2020.02.018 |
| Discharged from all hospitalized patients with COVID 19 | 8 | 50 | | | | | | | % | | | | | | Anhui (China) | | 22 | | 75 | Data from Jan 22 to Feb 20, 2020 | Huang Lei et al. 2020 | https://doi.org/10.1016/j.jinf.2020.03.006 |
| Discharged from all hospitalized patients with COVID 19 | 417 | 71.7 | | | | | | | % | | | | | | Shenzhen (China) | 45.4 | | | 47.2 | Cases from Jan 1 Feb 28, 2020 | Wen et al. 2020 | https://doi.org/10.1101/2020.03.22.20035246 |
| Discharged from all hospitalized patients with COVID 19 | 91 | 34.1 | | | | | | | % | | | | | | Zhejiang province (China) | | 50 | | 40.7 | Data colected from Jan 20 to Feb 11, 2020. Final folow-up until Feb 16, 2020 | Qian et al. 2020 | https://doi.org/10.1093/qjmed/hcaa089 |
| Discharged from all hospitalized patients with COVID 19 | 46 | 78.3 | | | | | | | % | | | | | | Chongqing (China) | | | 10-35 | 52.2 | Data from Jan 25 to Feb 18, 2020. Outcomes until Feb 23, 2020 | Liao et al. 2020 | https://doi.org/10.1016/j.xinn.2020.04.001 |
| Discharged from all hospitalized patients with COVID 19 | 18 | 44.4 | | | | | | | % | | | | | | Singapore | | 47 | | 50 | Data from Jan 23 to Feb 3, 2020. Final follow-up was Feb 25, 2020 | Young et al. 2020 | https://doi.org/10.1001/jama.2020.204 |
| Discharged from all hospitalized patients with COVID 19 | 18 | 33.3 | | | | | | | % | | | | | | Zhengzou (China) | | 39 | | 55.6 | Data from Jan 21 to Feb 5. Final follo-up was Feb 7, 2020 | Wang Lei et al. 2020 | https://doi.org/10.1183/13993003.00398-2020 |
| Discharged from all hospitalized patients with COVID 19 | 985 | 30.8 | | | | | | | % | | | | | | Wuhan (China) | | | | | Data from Jan 30 to Feb 25, 2020 (outcomes included) | Luo et al. 2020 | https://doi.org/10.1101/2020.03.19.20033175 |
| Discharged from all hospitalized patients with COVID 19 | 93 | 66.7 | | | | | | | % | | | | | | Singapore | | | | | Data from Jan 2 to Feb 26, 2020 | Tindale et al. 2020 | https://doi.org/10.7554/eLife.57149 |
| Discharged from all hospitalized patients with COVID 19 | 56 | 100 | | | | | | | % | | | | | | Jilin (China) | | | | | Data from Jan 21 to March 5, 2020 | Tian Suyan et al .2020 | https://doi.org/10.1007/s12250-020-00317-z |
| **Discharged from ICU (%)** | | | | | | | | | | | | | | | | | | | | | | |
| Discharged from ICU | 13 | 54 | | | | | | | % | | | | | | Patients from Wuhan Hospital | | | | | Data from Dec 16, 2019 to Jan 22, 2020 | Huang Chaolin et al. 2020 | https://doi.org/10.1016/S0140-6736(20)30183-5/ |
| Discharged from ICU | 36 | 25 | | | | | | | % | | | | | | Wuhan (China) | | 66 | | 61.1 | Data from Jan 1 to Feb 3, 2020 | Wang Dawei et al. 2020 | https://doi.org/10.1001/jama.2020.1585 |
| Discharged from ICU | 24 | 21 | | | | | | | % | | | | | | Seattle region (USA) | | | | | | Bhatraju et al. 2020 | https://doi.org/10.1056/NEJMoa2004500 |
| Discharged from ICU | 8 | 85.7 | | | | | | | % | | | | | | Hong Kong | | 64.5 | | 50 | Patients admitted from Jan 22 to Feb 11, 2020 | Ling Lowell et al. 2020 | https://www.ncbi.nlm.nih.gov/pubmed/32248675 |
| Discharged from ICU | 52 | 15.4 | | | | | | | % | | | | | | Wuhan (China) | | | | | Data from late Dec, 2019 to Feb 8, 2020 | Yang et al. 2020 | https://doi.org/10.1016/S2213-2600(20)30079-5 |
| Discharged from ICU | 21 | 9.5 | | | | | | | % | | | | | | Washington State (USA) | 70 | | | 52 | | Arentz et al. 2020 | https://doi.org/10.1001/jama.2020.4326 |
| Discharged from ICU | 34 | 58.8 | | | | | | | % | | | | | | Hangzhou (China) | | 66 | | 67.6 | Data from Jan 22 to March 5, 2020 | Zheng Xi et al. 2020 | https://doi.org/10.1631/jzus.b2000174 |

| | | | | | | | | | | | | | | | | | | | | | | |
|---|---|---|---|---|---|---|---|---|---|---|---|---|---|---|---|---|---|---|---|---|---|---|
| Discharged from ICU | 344 | 53.8 | | | | | | | % | | | | | | Wuhan (China) | | 71 | | 55.6 | Patients enrolled from Jan 25 to Feb 25, 2020 | Wang Yang et al. 2020 | https://doi.org/10.1164/rccm.202003-0736LE |
| Discharged from ICU | 33 | 27 | | | | | | | % | | | | | | Brescia (Italy) | 64 | | | 91 | | Piva et al. 2020 | https://doi.org/10.1016/j.jcrc.2020.04.004 |
| **In hospital (%)** | | | | | | | | | | | | | | | | | | | | | | |
| In hospital | 73 | 35.6 | | | | | | | % | | | | | | Wuhan (China) | | 67 | | 61.6 | Data from December 24, 2019 to February 7,2020 | Tang et al. 2020 | https://doi.org/10.1016/j.chest.2020.03.032 |
| In hospital | 67 | 65.7 | | | | | | | % | | | | | | Wuhan (China) | | 42 | | 46 | Patients hospitalized from Jan 16 to Jan 29, 2020. Outcome follow-up Feb 4,2020 | Wang Zhongliang et al. 2020 | https://doi.org/10.1093/cid/ciaa272 |
| In hospital | 62 | 98 | | | | | | | % | | | | | | Zhejiang (China) | | 41 | | 56 | Data from Jan 10 to Jan 26, 020 | Xu Xiao-Wei et al. 2020 | http://dx.doi.org/10.1136/bmj.m606 |
| In hospital | 99 | 58 | | | | | | | % | | | | | | Wuhan (China) | 55.5 | | | 68 | Data from Jan 1 to Jan 25, 2020 | Chen N et al. 2020 | https://doi.org/10.1016/S0140-6736(20)30211-7 |
| In hospital | 168 | 1.2 | | | | | | | % | | | | | | Hainan (China) | | | | | Data from Jan 22 to March 13, 2020 | Yan et al. 2020 | https://doi.org/10.1101/2020.03.19.20038539 |
| In hospital | 77 | 10.4 | | | | | | | % | | | | | | Beijing (China) | | | | | Patients admited from Jan 21 to February 8, 2020 Outcomes follow-up to Feb 29,2020 | Zhao Wen et al. 2020 | https://doi.org/10.1101/2020.03.13.20035436 |
| In hospital * | 221 | 76 | | | | | | | % | | | | | | Wuhan (China) | | 55 | | 48.9 | Data from Jan 2 to Feb 10, 2020 Outcomes follow up to Feb 15, 2020 | Zhang Guqin et al. 2020 | https://doi.org/10.1016/j.jcv.2020.104364 |
| In hospital | 138 | 61.1 | | | | | | | % | | | | | | Wuhan (China) | | 56 | | 54.3 | Data from Jan 1 to Feb 3, 2020 | Wang Dawei et al. 2020 | https://doi.org/10.1001/jama.2020.1585 |
| In hospital * | 267 | 59.9 | | | | | | | % | | | | | | Chongqing (China) | | 48 | | 55.8 | Data from Jan 19 to Feb 16, 2020 | Qi Di et al. 2020 | https://doi.org/10.1101/2020.03.01.20029397 |
| In hospital * | 291 | 44.7 | | | | | | | % | | | | | | Hunan (China) | | 46 | | 49.8 | Data from Jan 23 to Feb 14, 2020. Outcomes until Feb 20, 2020 | Chen X et al. 2020 | https://doi.org/10.1101/2020.03.03.20030353 |
| In hospital * | 1099 | 93.6 | | | | | | | % | | | | | | Mainland China | | | | 58.1 | Data up to Jan 29, 2020 | Guan et al. 2020 | https://doi.org/10.1056/NEJMoa2002032 |
| In hospital * | 41 | 17 | | | | | | | % | | | | | | Patients from Wuhan Hospital (China) | | 49 | | 73 | Data from Dec 16, 2019 to Jan 22, 2020 | Huang Chaolin et al. 2020 | https://doi.org/10.1016/S0140-6736(20)30183-5 |
| In hospital | 24 | 29.2 | | | | | | | % | | | | | | Seattle region (USA) | | | | | | Bhatraju et al. 2020 | https://doi.org/10.1056/NEJMoa2004500 |
| In hospital | 50 | 60 | | | | | | | % | | | | | | Hong Kong | 55.2 | | | 46 | Data from Jan 26 to Feb 28, 2020 | Leung et al. 2020 | https://doi.org/10.3201/eid2701.201543 |
| In hospital | 1999 | 36.3 | | | | | | | % | | | | | | New York city (USA) | | 62 | | 62.6 | | Petrilli et al. 2020 | https://doi.org/10.1101/2020.04.08.20057794 |
| In hospital | 799 | 65.7 | | | | | | | % | | | | | | Wuhan (China) | | | | | Data from Jan 13 to Feb 28, 2020 | Chen T et al. 2020 | http://dx.doi.org/10.1136/bmj.m1091 |
| In hospital | 125 | 62.4 | | | | | | | % | | | | | | Anhui (China) | 38.76 | | | 56.8 | Data from Jan 20 to Feb 18,2020 | Wang R et al. 2020 | https://doi.org/10.1016/j.ijid.2020.03.070 |
| In hospital | 137 | 11.7 | | | | | | | % | | | | | | Hubei (China) | | 57 | | 44.5 | Patients admitted from Dec 30, 2019 to Jan 24, 2020 | Liu Kiu et al. 2020 | https://doi.org/10.1097/CM9.0000000000000744 |
| In hospital | 20 | 30 | | | | | | | % | | | | | | Guangzhou (China) | 43.2 | | | 50 | Patients admitted from Jan 22 to Feb | Lei et al. 2020 | https://doi.org/10.1016/j.tmaid.2020.101664 |

| | | | | | | | | | | | | | | | | | | | | | | |
|---|---|---|---|---|---|---|---|---|---|---|---|---|---|---|---|---|---|---|---|---|---|---|
| | | | | | | | | | | | | | | | | | | | | 12, 2020. Outcomes follow-up Feb 18, 2020 | | |
| In hospital | 4 | 50 | | | | | | | % | | | | | | Shangai (China) | 44.25 | | | 75 | Data from Jan 21 to Feb 4, 2020 | Wang Zhenwei et al. 2020 | https://doi.org/10.5582/bst.2020.01030 |
| In hospital | 80 | 76.25 | | | | | | | % | | | | | | Jiangsu (China) | 46.1 | | | 48.75 | Patients enrolled from Jan 22 to Feb 14, 2020 | Wu Jian et al. 2020 | https://doi.org/10.1093/cid/ciaa199 |
| In hospital | 8 | 50 | | | | | | | % | | | | | | Anhui (China) | | 22 | | 75 | Data from Jan 22 to Feb 20, 2020 | Huang Lei et al. 2020 | https://doi.org/10.1016/j.jinf.2020.03.006 |
| In hospital * | 262 | 81.7 | | | | | | | % | | | | | | Beijing (China) | | 47.5 | | 48.5 | Data from Jan 20 to Feb 10, 2020 | Tian Sijia et al. 2020 | https://doi.org/10.1016/j.jinf.2020.02.018 |
| In hospital | 417 | 27.6 | | | | | | | % | | | | | | Shenzhen (China) | 45.4 | | | 47.2 | Cases from Jan 1 Feb 28, 2020 | Wen et al. 2020 | https://doi.org/10.1101/2020.03.22.20035246 |
| In hospital | 91 | 65.9 | | | | | | | % | | | | | | Zhejiang province (China) | | 50 | | 40.7 | Data colected from Jan 20 to Feb 11, 2020. Final folow-up until Feb 16, 2020 | Qian et al. 2020 | https://doi.org/10.1093/qjmed/hcaa089 |
| In hospital | 46 | 21.7 | | | | | | | % | | | | | | Chongqing (China) | | | 10-35 | 52.2 | Data from Jan 25 to Feb 18, 2020. Outcomes until Feb 23, 2020 | Liao et al. 2020 | https://doi.org/10.1016/j.xinn.2020.04.001 |
| In hospital | 18 | 55.6 | | | | | | | % | | | | | | Singapore | | 47 | | 50 | Data from Jan 23 to Feb 3, 2020. Final follow-up was Feb 25, 2020 | Young et al. 2020 | https://doi.org/10.1001/jama.2020.204 |
| In hospital | 18 | 66.6 | | | | | | | % | | | | | | Zhengzou (China) | | 39 | | 55.6 | Data from Jan 21 to Feb 5. Final follo-up was Feb 7, 2020 | Wang Lei et al. 2020 | https://doi.org/10.1183/13993003.00398-2020 |
| In hospital | 985 | 58.3 | | | | | | | % | | | | | | Wuhan (China) | | | | | Data from Jan 30 to Feb 25, 2020 (outcomes included) | Luo et al. 2020 | https://doi.org/10.1101/2020.03.19.20033175 |
| **In hospital (ICU) (%)** | | | | | | | | | | | | | | | | | | | | | | |
| In hospital (ICU) | 52 | 23.1 | | | | | | | % | | | | | | Wuhan (China) | | | | | Patients enrolled between late Dec 2019, and Jan 26, 2020. Primary outcome Feb 9, 2020 | Yang et al. 2020 | https://doi.org/10.1016/S2213-2600(20)30079-5 |
| In hospital (ICU) | 21 | 24 | | | | | | | % | | | | | | Washington State (USA) | 70 | | | 52 | Data up to March 17 | Arentz et al. 2020 | https://doi.org/10.1001/jama.2020.4326 |
| In hospital (ICU) | 8 | 14 | | | | | | | % | | | | | | Hong Kong | | 64.5 | | 50 | Patients admitted from Jan 22 to Feb 11, 2020 | Ling Lowell et al. 2020 | https://www.ncbi.nlm.nih.gov/pubmed/32248675 |
| In hospital (ICU) | 36 | 30.6 | | | | | | | % | | | | | | Wuhan (China) | | 66 | | 61.1 | Data from Jan 1 to Feb 3, 2020 | Wang Dawei et al. 2020 | https://doi.org/10.1001/jama.2020.1585 |
| In hospital (ICU) | 24 | 12.5 | | | | | | | % | | | | | | Seattle region (USA) | | | | | | Bhatraju et al. 2020 | https://doi.org/10.1056/NEJMoa2004500 |
| In hospital (ICU) | 13 | 8 | | | | | | | % | | | | | | Patients from Wuhan Hospital (China) | | 49 | | 85 | Data from Dec 16, 2019 to Jan 22, 2020 | Huang Chaolin et al. 2020 | https://doi.org/10.1016/S0140-6736(20)30183-5 |
| In hospital (ICU) | 50 | 6 | | | | | | | % | | | | | | Hong Kong | 55.2 | | | 46 | Data from Jan 26 to Feb 28, 2020 | Leung et al. 2020 | https://doi.org/10.3201/eid2701.201543 |
| In hospital (ICU) | 34 | 41.2 | | | | | | | % | | | | | | Hangzhou (China) | | 66 | | 67.6 | Data from Jan 22 to March 5, 2020 | Zheng Xi et al. 2020 | https://doi.org/10.1631/jzus.b2000174 |
| In hospital (ICU) | 344 | 7.5 | | | | | | | % | | | | | | Wuhan (China) | | 70 | | 55.6 | Patients enllored from Jan 25 to Feb 25, 2020 | Wang Yang et al. 2020 | https://doi.org/10.1164/rccm.202003-0736LE |

| | | | | | | | | | | | | | | | | | | | | | | |
|---|---|---|---|---|---|---|---|---|---|---|---|---|---|---|---|---|---|---|---|---|---|---|
| In hospital (ICU) | 33 | 70 | | | | | | | % | | | | | | Brescia (Italy) | | 64 | | 91 | | Piva et al. 2020 | https://doi.org/10.1016/j.jcrc.2020.04.004 |
| **CHILDREN** | | | | | | | | | | | | | | | | | | | | | | |
| **Incubation period (days)** | | | | | | | | | | | | | | | | | | | | | | |
| Incubation period | 7 | | | | | | | | Days | 5 | | | 3 | 12 | Shaanxi (China) | | 1.3 | | 57.1 | Data from Jan 31 to Feb 16, 2020 | Han et al. 2020 | https://doi.org/10.1002/jmv.25835 |
| Incubation period ** | 6 | 7.17 | 1-16 | | | 4.7 9 | | | Days | 7.5 | | | | | Changsha (China) | | 8 | | 33.3 | Data from Jan 8 to Feb 19, 2020. Outcomes follow-up to Feb 26 | Shen et al. 2020 | https://doi.org/10.1002/ppul.24762 |
| Incubation period | 12 | 8 | 1-13 | | | | | | Days | | | | | | Chongqing (China) | | 14.5 | | 50 | Patients recuited from Jan 28 to Feb 11,2020 | Chen et al. 2020 | https://doi.org/10.1016/j.gendis.2020.03.008 |
| Incubation period ** | 4 | 6.75 | | | | 2.3 6 | | | Days | | | | | | Wuhan (China) | | 4.6 | | 75 | Patients treated from Jan 24 to Feb 24, 2020 | Sun et al. 2020 | https://doi.org/10.1007/s12519-020-00354-4 |
| Incubation period ** | 8 | 6.5 | | | | 2.6 7 | | | Days | | | | | | China | | 6.5 | | 40 | | Cai et al. 2020 | https://doi.org/10.1093/cid/ciaa198 |
| **Onset of symptoms/illness onset to diagnosis (days)** | | | | | | | | | | | | | | | | | | | | | | |
| Onset of symptoms to diagnosis | 728 | | | | | | | | Days | 3 | | | 1 | 5 | China | | 10 | | 57.4 | Data from Jan 16 to Feb 8, 2020 | Dong et al. 2020 | https://doi.org/10.1542/peds.2020-0702 |
| Illness onset to diagnosis ** | 7 | 7.75 | | | | 5.1 2 | | | Days | | | | | | Wuhan (China) | | 4.9 | | 75 | Patients treated from Jan 24 to Feb 24, 2020 | Sun et al. 2020 | https://doi.org/10.1007/s12519-020-00354-4 |
| Illness onset to diagnosis ** | 9 | 1.56 | | | | 0.8 8 | | | Days | | | | | | China | | 0.5 | | 22.2 | | Wei Min et al. 2020 | https://doi.org/10.1001/jama.2020.2131 |
| **Onset of symptoms/illness onset to hospital admission (days)** | | | | | | | | | | | | | | | | | | | | | | |
| Onset of symptoms to hospital admission ** | 9 | 6.11 | 0-17 | | | 6.3 3 | | | Days | 3 | | | | | Changsha (China) | | 8 | | 33.3 | Data from Jan 8 to Feb 19, 2020. Outcomes follow-up to Feb 26 | Shen et al. 2020 | https://doi.org/10.1002/ppul.24762 |
| **Hospital length stay (days)** | | | | | | | | | | | | | | | | | | | | | | |
| Hospital stay | 10 | 17.2 | | | | 4.9 | | | Days | | | | | | Changsha (China) | | 7 | | 30 | Children recruited from Jan 27 to March 10, 2020 | Tan et al. 2020 | https://doi.org/10.1016/j.jcv.2020.104353 |
| Hospital stay ** | 6 | 8.33 | | | | 2.8 | | | Days | | | | | | Wuhan (China) | | 3 | | 33.3 | Patients enrolled from Jan 7 to Jan 15, 2020 | Liu Weiyong et al. 2020 | https://doi.org/10.1056/NEJMc2003717 |
| Hospital stay | 36 | 14 | | | | 3 | | | Days | | | | | | Zheijiang (China) | | 6 | | 64 | Data from Jan 17 to March 1, 2020 | Qiu et al. 2020 | https://doi.org/10.1016/S1473-3099(20)30198-5 |
| **Asymptomatic patients (%)** | | | | | | | | | | | | | | | | | | | | | | |
| Asymptomatic patients | 171 | 15.8 | | | | | | | % | | | | | | Wuhan (China) | | 6.7 | | 60.8 | Children assessed and tested from Jan 28 to Feb 26, 2020 | Lu et al. 2020 | https://doi.org/10.1056/NEJMc2005073 |
| Asymptomatic patients | 10 | 20 | | | | | | | % | | | | | | Changsha (China) | | 7 | | 30 | Children recruited from Jan 27 to March 10, 2020 | Tan et al. 2020 | https://doi.org/10.1016/j.jcv.2020.104353 |
| Asymptomatic patients * | 728 | 12.9 | | | | | | | % | | | | | | China | | 10 | | 57.4 | Data from Jan 16 to Feb 8, 2020 | Dong et al. 2020 | https://doi.org/10.1542/peds.2020-0702 |
| Asymptomatic patients | 36 | 28 | | | | | | | % | | | | | | Zheijiang (China) | | 8.3 | | 64 | Data from Jan 17 to March 1, 2020 | Qiu et al. 2020 | https://doi.org/10.1016/S1473-3099(20)30198-5 |

| | | | | | | | | | | | | | | | | | | | | | | |
|---|---|---|---|---|---|---|---|---|---|---|---|---|---|---|---|---|---|---|---|---|---|---|
| **ICU admissions vs all hospitalized patients with COVID-19 (%)** | | | | | | | | | | | | | | | | | | | | | | |
| ICU admissions vs all hospitalized patients with COVID 19 | 171 | 1.75 | | | | | | | % | | | | | | Wuhan (China) | | 6.7 | | 60.8 | Children assessed and tested from Jan 28 to Feb 26, 2020 | Lu et al. 2020 | https://doi.org/10.1056/NEJMc2005073 |
| ICU admissions vs all hospitalized patients with COVID 19 | 9 | 0 | | | | | | | % | | | | | | Changsha (China) | | 8 | | 33.3 | Data from Jan 8 to Feb 19, 2020. Outcomes follow-up to Feb 26 | Shen et al. 2020 | https://doi.org/10.1002/ppul.24762 |
| ICU admissions vs all hospitalized patients with COVID 19 | 25 | 8 | | | | | | | % | | | | | | Hubei (China) | | 3 | | 56 | | Zheng et al. 2020 | https://doi.org/10.1007/s11596-020-2172-6 |
| ICU admissions vs all hospitalized patients with COVID 19 | 6 | 16.7 | | | | | | | % | | | | | | Wuhan (China) | | 3 | | 33.3 | Patients enrolled from Jan 7 to Jan 15, 2020 | Liu Weiyong al. 2020 | https://doi.org/10.1056/NEJMc2003717 |
| **Death vs all hospitalized patients with COVID 19 (%)** | | | | | | | | | | | | | | | | | | | | | | |
| Death vs all hospitalized patients with COVID 19 | 171 | 0.6 | | | | | | | % | | | | | | Wuhan (China) | | 6.7 | | 60.8 | Children assessed and tested from Jan 28 to Feb 26, 2020 | Lu et al. 2020 | https://doi.org/10.1056/NEJMc2005073 |
| **Discharged from all hospitalized patients with COVID 19 (%)** | | | | | | | | | | | | | | | | | | | | | | |
| Discharged from all hospitalized patients with COVID 19 | 171 | 12.3 | | | | | | | % | | | | | | Wuhan (China) | | 6.7 | | 60.8 | Children assessed and tested from Jan 28 to Feb 26, 2020 | Lu et al. 2020 | https://doi.org/10.1056/NEJMc2005073 |
| Discharged from all hospitalized patients with COVID 19 | 9 | 66.66 | | | | | | | % | | | | | | Changsha (China) | | 8 | | 33.3 | Data from Jan 8 to Feb 19, 2020. Outcomes follow-up to Feb 26 | Shen et al. 2020 | https://doi.org/10.1002/ppul.24762 |
| Discharged from all hospitalized patients with COVID 19 | 25 | 4 | | | | | | | % | | | | | | Hubei (China) | | 3 | | 56 | Patients admitted between Feb 1 to Feb 10 | Zheng Fang et al. 2020 | https://doi.org/10.1007/s11596-020-2172-6 |
| **Discharged from ICU (%)** | | | | | | | | | | | | | | | | | | | | | | |
| Discharged from ICU | 8 | 62.5 | | | | | | | % | | | | | | Wuhan (China) | | 4.6 | | 75 | Patients treated from Jan 24 to Feb 24, 2020 | Sun et al. 2020 | https://doi.org/10.1007/s12519-020-00354-4 |
| **In hospital (%)** | | | | | | | | | | | | | | | | | | | | | | |
| In hospital | 171 | 87.1 | | | | | | | % | | | | | | Wuhan (China) | | 6.7 | | 60.8 | Children assessed and tested from Jan 28 to Feb 26, 2020 | Lu et al. 2020 | https://doi.org/10.1056/NEJMc2005073 |
| In hospital | 9 | 33.3 | | | | | | | % | | | | | | Changsha (China) | | 8 | | 33.3 | Data from Jan 8 to Feb 19, 2020. Outcomes follow-up to Feb 26 | Shen et al. 2020 | https://doi.org/10.1002/ppul.24762 |
| In hospital | 25 | 96 | | | | | | | % | | | | | | Hubei (China) | | 3 | | 56 | Patients admitted between Feb 1 to Feb 10 | Zheng Fang et al. 2020 | https://doi.org/10.1007/s11596-020-2172-6 |
| **In hospital (ICU) (%)** | | | | | | | | | | | | | | | | | | | | | | |
| In hospital (ICU) | 8 | 37.5 | | | | | | | % | | | | | | Wuhan (China) | | 4.6 | | 75 | Patients treated from Jan 24 to Feb 24, 2020 | Sun et al. 2020 | https://doi.org/10.1007/s12519-020-00354-4 |

Abbreviations: ICU, Intensive Care Unit.
* Papers include disaggregated data (by age, medical condition, etc.), apart from overall data.
** Parameters calculated from data shown in tables or supplementary material.
