## Supplementary material for "Scoping review and meta-analysis of COVID-19 epidemiological parameters for modeling from early Asian studies": Table S2

**Supplementary Table S2.** Quality assessment of selected studies.

| | Selection | Ascertainment | Causality | Reporting |
|---|---|---|---|---|
| Bi et al. 2020 | + | ? | + | + |
| Cai et al. 2020 | + | ? | + | + |
| Cao Jianlei et al. 2020 | + | + | + | + |
| Cao Bin et al. 2020 | - | + | + | + |
| Chan et al. 2020 | ? | + | + | + |
| Chen TL et al. 2020 | + | + | ? | + |
| Chen N et al. 2020 | + | + | - | + |
| Chen T et al. 2020 | + | + | + | + |
| Chen X et al. 2020 | + | + | ? | + |
| Deng et al. 2020 | + | + | + | + |
| Dong et al. 2020 | + | + | + | + |
| Dorigatti et al. 2020 | ? | + | + | + |
| Du Yingzhen et al. 2020 | + | + | + | + |
| Du Rong-Hui et al. 2020 | + | + | + | + |
| Du Zhanwei et al. 2020 | + | ? | + | + |
| Ganyani et al. 2020 | + | - | + | + |
| Guan et al. 2020 | + | + | - | + |
| Guo et al. 2020 | + | ? | + | + |
| Han et al. 2020 | + | ? | + | + |
| He Xi et al. 2020 | ? | + | + | + |
| He Guiqing et al. 2020 | + | + | ? | + |
| Hong et al. 2020 | + | + | + | + |
| Hu et al. 2020 | - | + | + | + |
| Huang Lei et al. 2020 | - | + | + | + |
| Huang Chaolin et al. 2020 | + | + | + | + |
| Huang Guoqan et al. 2020 | ? | + | + | + |
| Jiang et al. 2020 | + | + | + | + |
| Jung et al. 2020 | + | ? | + | + |
| Lauer et al. 2020 | + | ? | + | + |
| Le et al. 2020 | + | + | + | + |
| Lei et al. 2020 | + | + | ? | + |
| Leung et al. 2020 | + | + | - | + |
| Li et al. 2020 | + | + | + | + |
| Lian et al. 2020 | + | + | + | + |
| Liang et al. 2020 | + | + | + | + |
| Liao et al. 2020 | - | + | ? | + |
| Ling Lowell et al. 2020 | ? | + | + | + |
| Ling Zhoukun et al. 2020 | + | ? | ? | ? |
| Liu Tao et al. 2020 | + | + | + | + |
| Liu Lei et al. 2020 | + | + | + | + |
| Liu Kiu et al. 2020 | + | + | ? | + |
| Liu Weiyong et al. 2020 | + | + | + | + |
| Lo et al. 2020 | + | + | + | + |
| Lu et al. 2020 | + | + | ? | + |
| Luo et al. 2020 | + | + | - | + |
| Mao et al. 2020 | ? | + | + | + |
| Men et al. 2020 | ? | + | + | + |
| Meng et al. 2020 | - | + | + | + |
| Miao et al. 2020 | + | + | + | + |

| | | | | |
|---|---|---|---|---|
| Mizumoto et al. 2020 | ? | + | + | + |
| Mo et al. 2020 | + | + | + | + |
| Ng et al. 2020 | + | ? | + | + |
| Pan et al. 2020 | - | ? | ? | ? |
| Pongpirul et al. 2020 | + | ? | + | + |
| Pung et al. 2020 | ? | + | + | + |
| Qi Di et al. 2020 | + | + | - | + |
| Qi Shi et al. 2020 | + | ? | + | + |
| Qian et al. 2020 | + | + | - | + |
| Qian et al. 2020b | - | + | + | + |
| Qiu et al. 2020 | + | + | + | + |
| Wang R et al. 2020 | + | + | ? | + |
| Shen et al. 2020 | + | + | + | + |
| Song et al. 2020 | ? | + | + | + |
| Sun et al. 2020 | ? | + | + | + |
| Tan et al. 2020 | + | + | + | + |
| Tang et al. 2020 | + | + | ? | + |
| Tao et al. 2020 | + | + | + | + |
| Tian Suochen et al. 2020b | ? | + | + | + |
| Tian Sijia et al. 2020 | + | + | - | + |
| Tian Suyan et al. 2020 | ? | + | + | + |
| Tian Suochen et al. 2020 | ? | + | + | + |
| Tindale et al. 2020 | + | + | + | + |
| Wang X et al. 2020 | ? | + | + | + |
| Wang Zhuo et al. 2020 | ? | ? | ? | ? |
| Wang Dawei et al. 2020 | + | + | - | + |
| Wang Zhongliang et al. 2020 | + | + | ? | + |
| Wang Lei et al. 2020 | + | + | - | + |
| Wang Zhenwei et al. 2020 | ? | + | ? | + |
| Wang Lang et al. 2020 | - | + | + | + |
| Wang Pei et al. 2020 | + | ? | + | + |
| Wang Yang et al. 2020 | - | + | + | + |
| Wei Wycliffe et al. 2020 | + | + | + | + |
| Wei Min et al. 2020 | + | ? | + | + |
| Wen et al. 2020 | + | + | - | + |
| Wu Joseph et al. 2020 | ? | + | + | + |
| Wu Jian et al. 2020 | + | + | - | + |
| Wu Chaomin et al. 2020 | + | + | + | + |
| Xia et al. 2020 | + | + | + | + |
| Xu Tianmin et al. 2020 | + | + | + | + |
| Xu Xiao-Wei et al. 2020 | + | + | - | + |
| Yan et al. 2020 | + | + | + | + |
| Yang et al. 2020 | + | + | + | + |
| Young et al. 2020 | + | + | + | + |
| Yuan et al. 2020 | + | + | + | + |
| Zhang Bicheng et al. 2020 | - | + | + | + |
| Zhang Lin et al. 2020 | - | + | + | + |
| Zhang Juanjuan et al. 2020 | ? | + | + | + |
| Zhang Guqin et al. 2020 | + | + | ? | + |
| Zhao Wen et al. 2020 | + | + | + | + |
| Zhao Shi et al. 2020 | ? | + | + | + |
| Zheng Xi et al. 2020 | + | + | ? | + |
| Zheng Fang et al. 2020 | + | + | - | + |

| | | | | |
|---|---|---|---|---|
| Zhou Fei et al. 2020 | + | + | + | + |
| Zhou Fating et al. 2020 | + | + | + | + |
| Zhou X et al. 2020 | + | + | + | + |
| Zou et al. 2020 | ? | + | + | + |

| | | | | | |
|---|---|---|---|---|---|
| + | Low risk of bias | ? | Unclear risk of bias | - | High risk of bias |

The selection domain evaluates whether the selection criteria is clear (e.g. a study that explicitly describes all the cases who have presented to a medical center over a certain period of time would satisfy this domain). The ascertainment assesses whether the outcome is adequately ascertained (e.g. use of clinical records is more reliable than self-report). The causality domain takes into account the follow-up time and if it was long enough for the results to be reliable (e.g. ≤ 5 days of follow-up was considered high risk of bias for death/discharged/In hospital outcomes). The reporting domain examines whether the study is described in sufficient details to allow other investigators to replicate the research or to make useful inferences.
