## Supplementary material for "Scoping review and meta-analysis of COVID-19 epidemiological parameters for modeling from early Asian studies": Text S1

**Supplementary Text S1.** Examples of electronic search strategy in PubMed

Example 1: Incubation period

((2020/1/1:2020/4/15[Date - Publication] AND (((((("covid 19"[All Fields] OR "covid 2019"[All Fields]) OR "severe acute respiratory syndrome coronavirus 2"[All Fields]) OR "2019 ncov"[All Fields]) OR "sars cov 2"[All Fields]) OR "2019ncov"[All Fields]) OR "novel coronavirus"[All Fields]))) AND ("incubation period"[All Fields])

Example 2: Hospital stay length

((2020/1/1:2020/4/15[Date - Publication] AND (((((("covid 19"[All Fields] OR "covid 2019"[All Fields]) OR "severe acute respiratory syndrome coronavirus 2"[All Fields]) OR "2019 ncov"[All Fields]) OR "sars cov 2"[All Fields]) OR "2019ncov"[All Fields]) OR "novel coronavirus"[All Fields]))) AND (((("length"[All Fields] AND "stay"[All Fields]) OR "length of stay"[All Fields] OR "hospital stay length" [All Fields]) OR ("hospital"[All Fields] AND "stay"[All Fields])) OR "hospital stay"[All Fields])
