## Supplementary material for "Scoping review and meta-analysis of COVID-19 epidemiological parameters for modeling from early Asian studies": Text S2

**Supplementary text 2.** Parameters definitions and reclassifications.

### ***Definitions***

- *Pre-symptomatic transmission*: transmission of SARS-COV-2 from an infected person –source patient- to a secondary patient –infectee- before the source patient developed symptoms.
- *Serial interval*: time duration between an infector having symptom onset and infectee having symptom onset.
- *Incubation period:* time from exposure to the development of symptoms.

### ***Reclassifications***

- *Hospital admission to death*: classified as hospital stay length of non-survivor cases.
- *Hospital admission to discharge*: classified as hospital stay length of survivor cases.
- *Onset of symptoms/illness onset to diagnosis* and *onset of symptoms/illness onset to hospital admission:* they have been pooled in a unique parameter because once a patient was diagnosed as a COVID-19 case usually the patient is admitted immediately.
