## Supplementary material for "Scoping review and meta-analysis of COVID-19 epidemiological parameters for modeling from early Asian studies": Figures S1-31

### Supplementary Figures S1-31. Forest plots of epidemiological parameters

We performed a meta-analysis for all epidemiological parameters considering all available studies that achieved the criteria detailed in the main text. Depending on the dates considered on the studies, we have distinguish two periods (A = Before February 12, 2020, B= whole period, December 2019 – March 2020), and in order to explore its impact in the meta-analysis, we have fitted to all studies a meta-regression model considering the period as a categorical moderator, where we allowed the amount of residual heterogeneity to be different in each subset. This has been done for each of the parameters, and for those where the period was a significant moderator we also present the meta-analysis for the two subsets defined by the period separately. In all cases, we also diagnosed for outliers and influencers. When outliers and/or influencers were identified, we performed separated meta-analysis to provide the results without those studies. All forest plots for all parameters and details from all meta-analysis results are provided herein. Notice that the high heterogeneity present in most cases when all studies are considered might reduce the use of the pulled effect, but when outliers were removed, the heterogeneity was significantly reduced. Thus, any conclusion from the meta-analysis could be considered reliable when it is robust to the meta-analysis when outliers and influencers are removed.

(Please note that only forest plots with all studies are shown when influencers or outliers were not detected).

i)

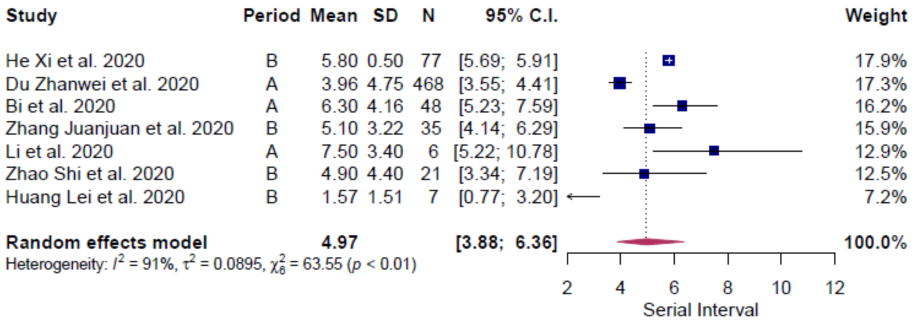

ii)

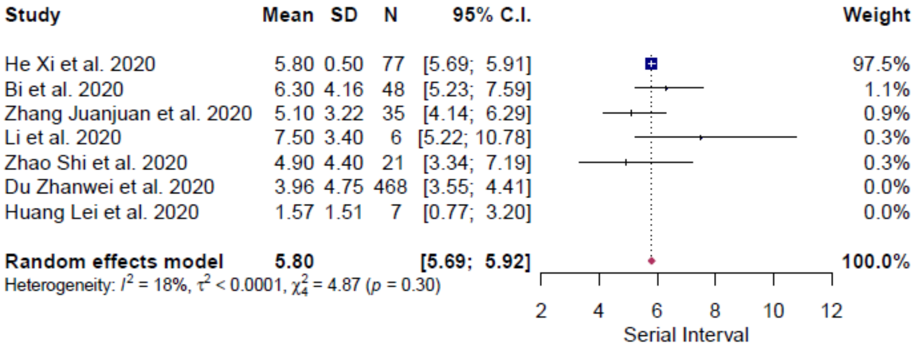

iii)

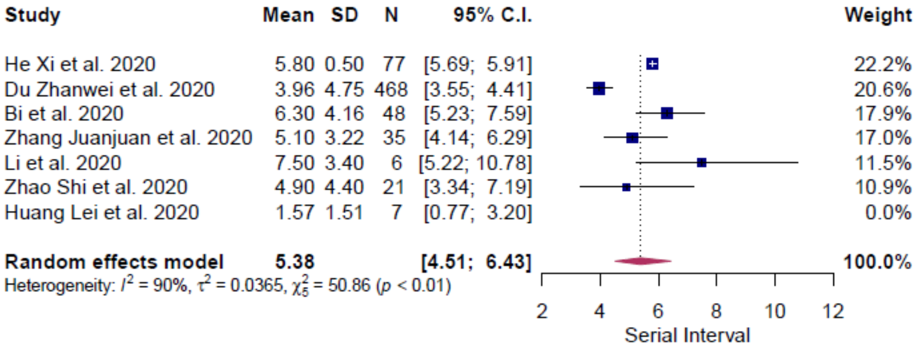

**Supplementary Figure S1.** Serial interval (general population). i) Results with all studies considered, ii) results with outliers removed and iii) results with influencers removed. Period: A, includes patients admitted before Feb 12, 2020; B, includes patients admitted before and after Feb 12, 2020.

i)

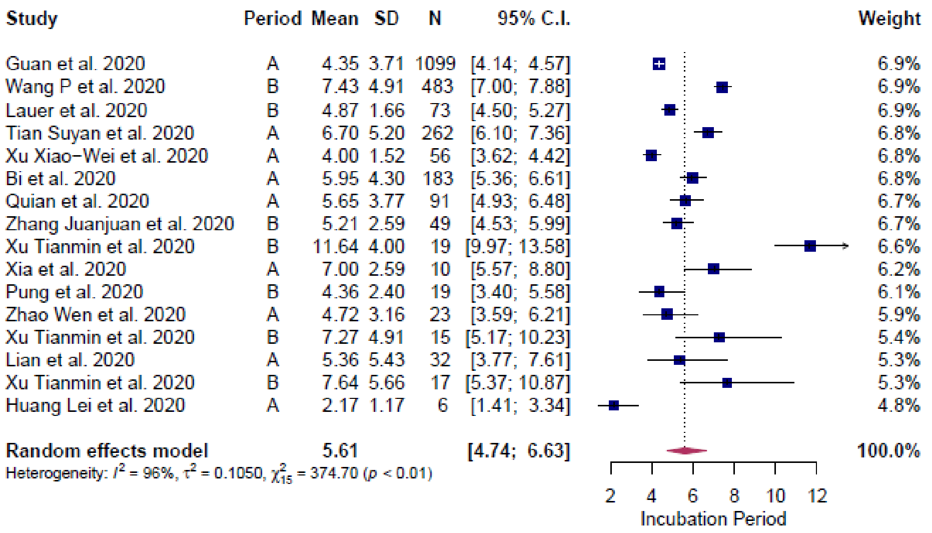
Study Period Mean SD N 95% C.I. Weight
Guan et al. 2020 A 4.35 3.71 1099 [4.14; 4.57] 6.9%
Wang P et al. 2020 B 7.43 4.91 483 [7.00; 7.88] 6.9%
Lauer et al. 2020 B 4.87 1.66 73 [4.50; 5.27] 6.9%
Tian Suyan et al. 2020 A 6.70 5.20 262 [6.10; 7.36] 6.8%
Xu Xiao−Wei et al. 2020 A 4.00 1.52 56 [3.62; 4.42] 6.8%
Bi et al. 2020 A 5.95 4.30 183 [5.36; 6.61] 6.8%
Quian et al. 2020 A 5.65 3.77 91 [4.93; 6.48] 6.7%
Zhang Juanjuan et al. 2020 B 5.21 2.59 49 [4.53; 5.99] 6.7%
Xu Tianmin et al. 2020 B 11.64 4.00 19 [9.97; 13.58] 6.6%
Xia et al. 2020 A 7.00 2.59 10 [5.57; 8.80] 6.2%
Pung et al. 2020 B 4.36 2.40 19 [3.40; 5.58] 6.1%
Zhao Wen et al. 2020 A 4.72 3.16 23 [3.59; 6.21] 5.9%
Xu Tianmin et al. 2020 B 7.27 4.91 15 [5.17; 10.23] 5.4%
Lian et al. 2020 A 5.36 5.43 32 [3.77; 7.61] 5.3%
Xu Tianmin et al. 2020 B 7.64 5.66 17 [5.37; 10.87] 5.3%
Huang Lei et al. 2020 A 2.17 1.17 6 [1.41; 3.34] 4.8%
Random effects model 5.61 [4.74; 6.63] 100.0%
Heterogeneity: $I^2$ = 96%, $\tau^2$ = 0.1050, $\chi^2_{15}$ = 374.70 ($p$ < 0.01)
2 4 6 8 10 12
Incubation Period

ii)

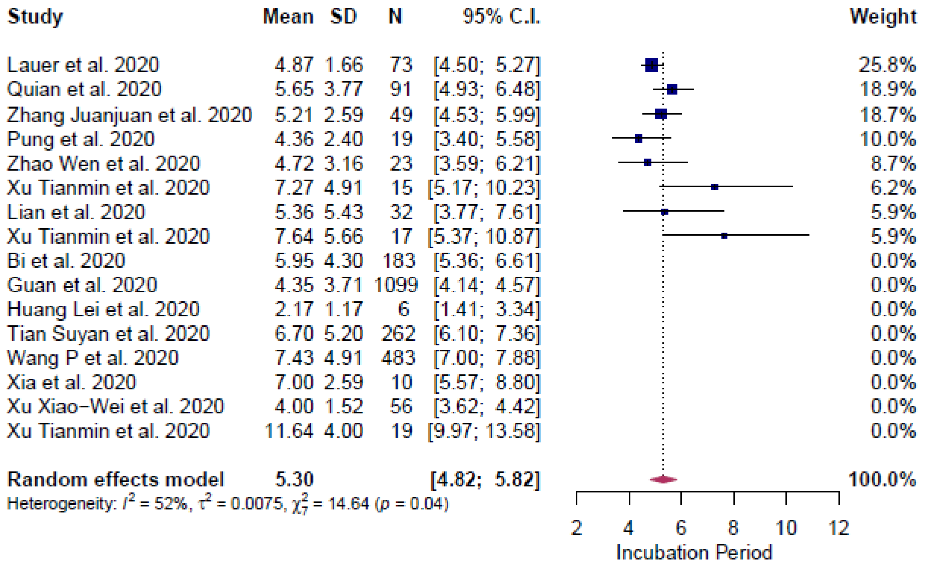
Study Mean SD N 95% C.I. Weight
Lauer et al. 2020 4.87 1.66 73 [4.50; 5.27] 25.8%
Quian et al. 2020 5.65 3.77 91 [4.93; 6.48] 18.9%
Zhang Juanjuan et al. 2020 5.21 2.59 49 [4.53; 5.99] 18.7%
Pung et al. 2020 4.36 2.40 19 [3.40; 5.58] 10.0%
Zhao Wen et al. 2020 4.72 3.16 23 [3.59; 6.21] 8.7%
Xu Tianmin et al. 2020 7.27 4.91 15 [5.17; 10.23] 6.2%
Lian et al. 2020 5.36 5.43 32 [3.77; 7.61] 5.9%
Xu Tianmin et al. 2020 7.64 5.66 17 [5.37; 10.87] 5.9%
Bi et al. 2020 5.95 4.30 183 [5.36; 6.61] 0.0%
Guan et al. 2020 4.35 3.71 1099 [4.14; 4.57] 0.0%
Huang Lei et al. 2020 2.17 1.17 6 [1.41; 3.34] 0.0%
Tian Suyan et al. 2020 6.70 5.20 262 [6.10; 7.36] 0.0%
Wang P et al. 2020 7.43 4.91 483 [7.00; 7.88] 0.0%
Xia et al. 2020 7.00 2.59 10 [5.57; 8.80] 0.0%
Xu Xiao−Wei et al. 2020 4.00 1.52 56 [3.62; 4.42] 0.0%
Xu Tianmin et al. 2020 11.64 4.00 19 [9.97; 13.58] 0.0%
Random effects model 5.30 [4.82; 5.82] 100.0%
Heterogeneity: $I^2$ = 52%, $\tau^2$ = 0.0075, $\chi^2_7$ = 14.64 ($p$ = 0.04)
2 4 6 8 10 12
Incubation Period

iii)

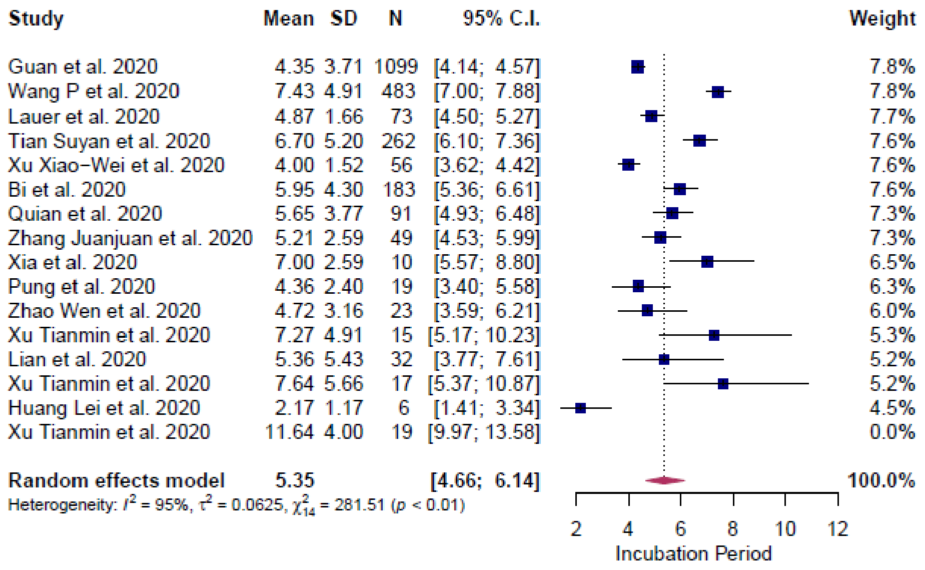

**Supplementary Figure S2.** Incubation period (general population). i) Results with all studies considered, ii) results with outliers removed and iii) results with influencers removed. Period: A, includes patients admitted before Feb 12, 2020; B, includes patients admitted before and after Feb 12, 2020.

i)

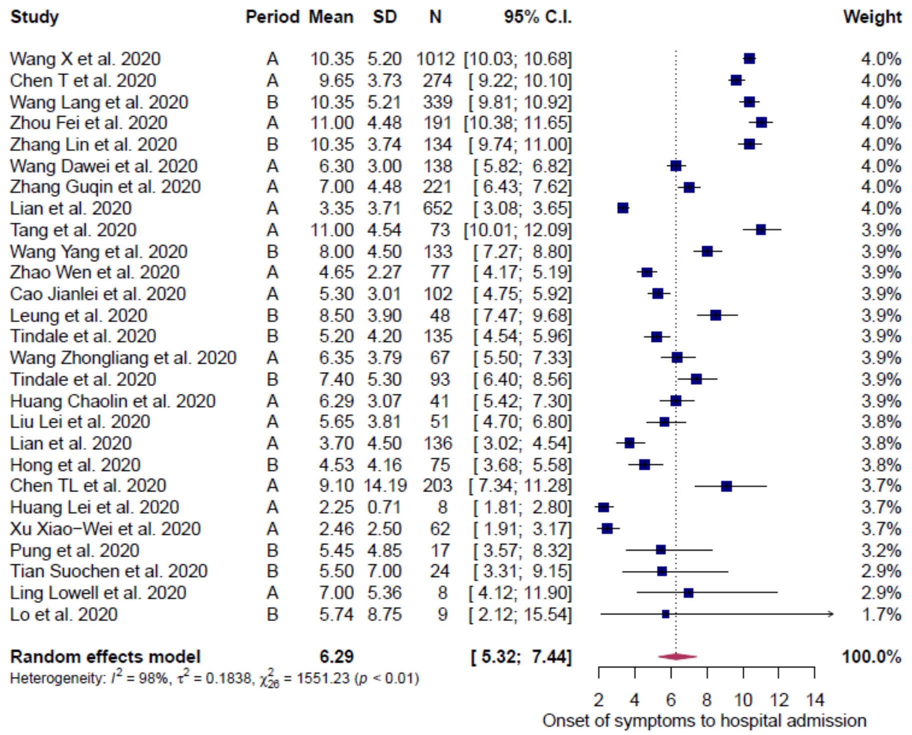

ii)

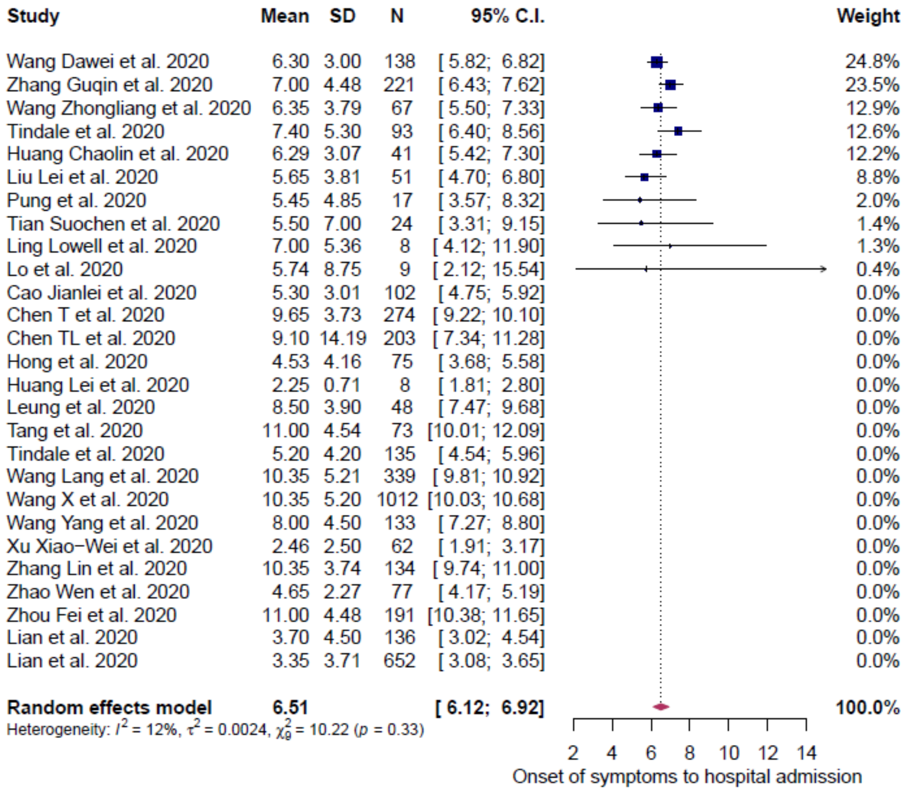

**Supplementary Figure S3.** Onset of symptoms to hospital admission – all (general population). i) Results with all studies considered and ii) results with outliers removed. Period: A, includes patients admitted before Feb 12, 2020; B, includes patients admitted before and after Feb 12, 2020.

i)

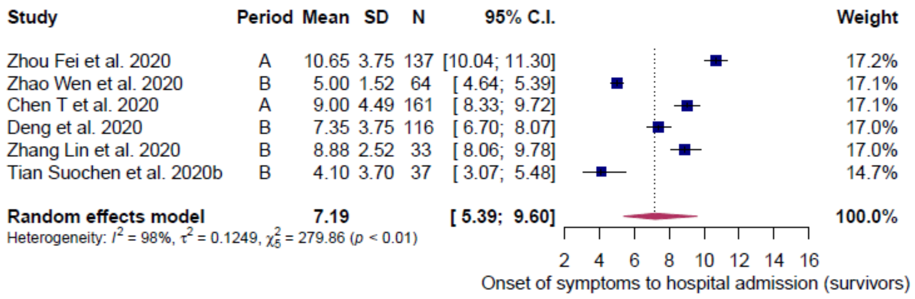

ii)

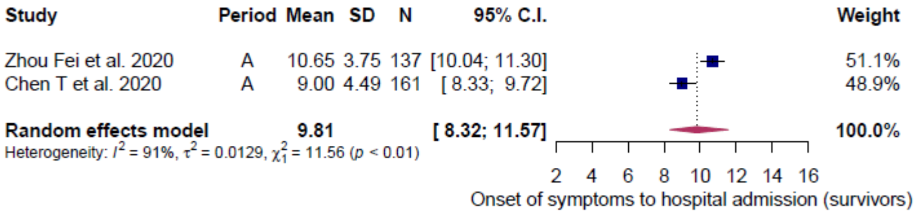

iii)

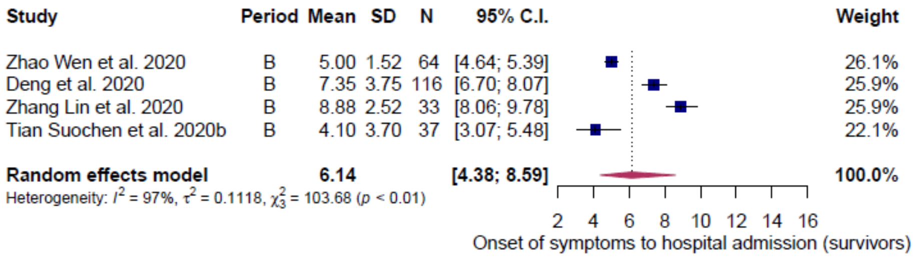

**Supplementary Figure S4.** Onset of symptoms to hospital admission –survivors (general population). i) Results with all studies considered, ii) results of period A (all studies considered) and iii) results of period B (all studies considered). Period: A, includes patients admitted before Feb 12, 2020; B, includes patients admitted before and after Feb 12, 2020.

i)

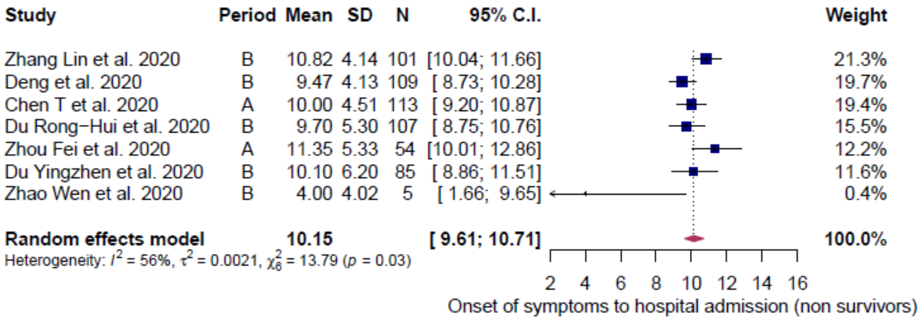

ii)

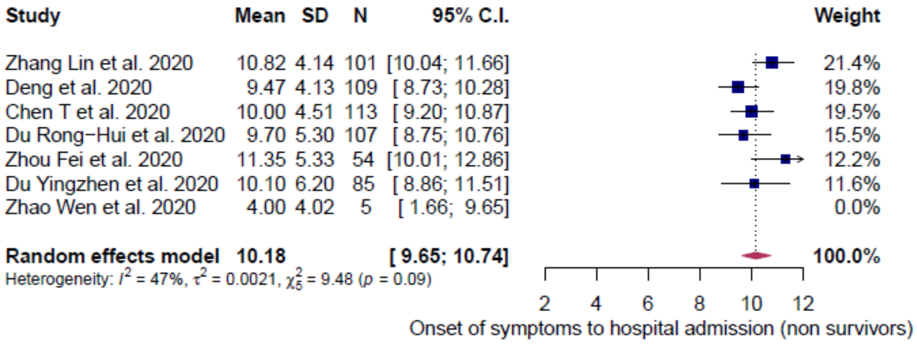

**Supplementary Figure S5.** Onset of symptoms to hospital admission – non survivors (general population). i) Results with all studies considered and ii) results with outliers removed. Period: A, includes patients admitted before Feb 12, 2020; B, includes patients admitted before and after Feb 12, 2020.

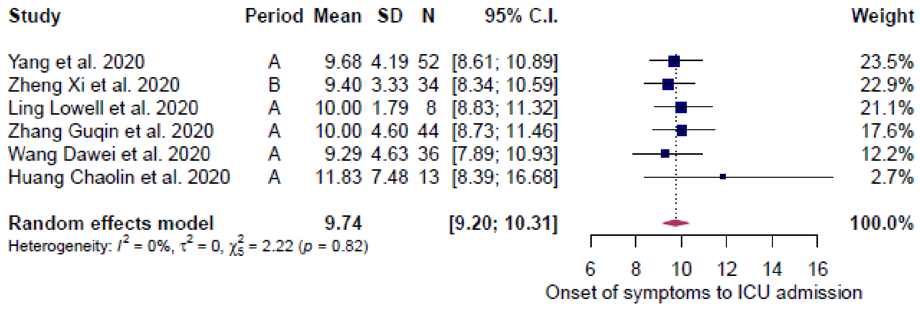

**Supplementary Figure S6**. Onset of symptoms to hospital ICU admission (general population). Results with all studies considered. Period: A, includes patients admitted before Feb 12, 2020; B, includes patients admitted before and after Feb 12, 2020.

i)

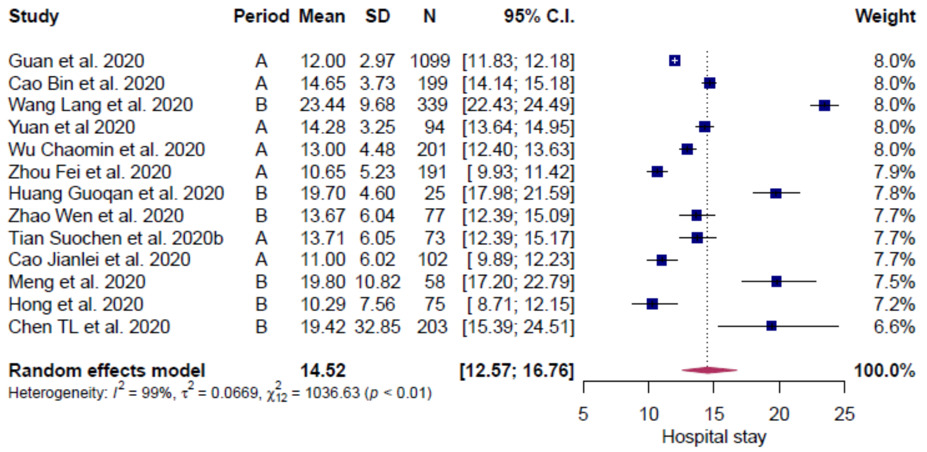
Study Period Mean SD N 95% C.I. Weight
Guan et al. 2020 A 12.00 2.97 1099 [11.83; 12.18] 8.0%
Cao Bin et al. 2020 A 14.65 3.73 199 [14.14; 15.18] 8.0%
Wang Lang et al. 2020 B 23.44 9.68 339 [22.43; 24.49] 8.0%
Yuan et al 2020 A 14.28 3.25 94 [13.64; 14.95] 8.0%
Wu Chaomin et al. 2020 A 13.00 4.48 201 [12.40; 13.63] 8.0%
Zhou Fei et al. 2020 A 10.65 5.23 191 [ 9.93; 11.42] 7.9%
Huang Guoqan et al. 2020 B 19.70 4.60 25 [17.98; 21.59] 7.8%
Zhao Wen et al. 2020 B 13.67 6.04 77 [12.39; 15.09] 7.7%
Tian Suochen et al. 2020b A 13.71 6.05 73 [12.39; 15.17] 7.7%
Cao Jianlei et al. 2020 A 11.00 6.02 102 [ 9.89; 12.23] 7.7%
Meng et al. 2020 B 19.80 10.82 58 [17.20; 22.79] 7.5%
Hong et al. 2020 B 10.29 7.56 75 [ 8.71; 12.15] 7.2%
Chen TL et al. 2020 B 19.42 32.85 203 [15.39; 24.51] 6.6%
Random effects model 14.52 [12.57; 16.76] 100.0%
Heterogeneity: $I^2$ = 99%, $\tau^2$ = 0.0669, $\chi^2_{12}$ = 1036.63 ($p$ < 0.01)
5 10 15 20 25
Hospital stay

ii)

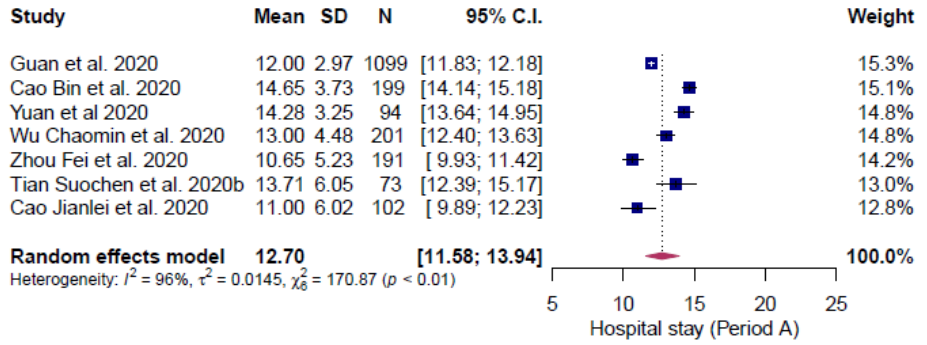
Study Mean SD N 95% C.I. Weight
Guan et al. 2020 12.00 2.97 1099 [11.83; 12.18] 15.3%
Cao Bin et al. 2020 14.65 3.73 199 [14.14; 15.18] 15.1%
Yuan et al 2020 14.28 3.25 94 [13.64; 14.95] 14.8%
Wu Chaomin et al. 2020 13.00 4.48 201 [12.40; 13.63] 14.8%
Zhou Fei et al. 2020 10.65 5.23 191 [ 9.93; 11.42] 14.2%
Tian Suochen et al. 2020b 13.71 6.05 73 [12.39; 15.17] 13.0%
Cao Jianlei et al. 2020 11.00 6.02 102 [ 9.89; 12.23] 12.8%
Random effects model 12.70 [11.58; 13.94] 100.0%
Heterogeneity: $I^2$ = 96%, $\tau^2$ = 0.0145, $\chi^2_6$ = 170.87 ($p$ < 0.01)
5 10 15 20 25
Hospital stay (Period A)

iii)

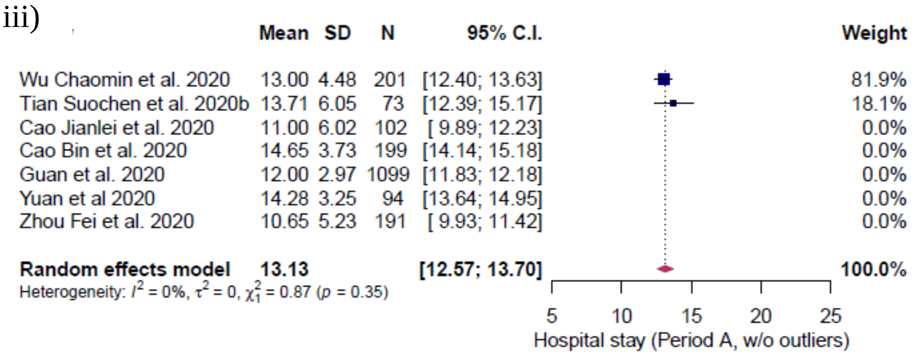
Mean SD N 95% C.I. Weight
Wu Chaomin et al. 2020 13.00 4.48 201 [12.40; 13.63] 81.9%
Tian Suochen et al. 2020b 13.71 6.05 73 [12.39; 15.17] 18.1%
Cao Jianlei et al. 2020 11.00 6.02 102 [ 9.89; 12.23] 0.0%
Cao Bin et al. 2020 14.65 3.73 199 [14.14; 15.18] 0.0%
Guan et al. 2020 12.00 2.97 1099 [11.83; 12.18] 0.0%
Yuan et al 2020 14.28 3.25 94 [13.64; 14.95] 0.0%
Zhou Fei et al. 2020 10.65 5.23 191 [ 9.93; 11.42] 0.0%
Random effects model 13.13 [12.57; 13.70] 100.0%
Heterogeneity: $I^2$ = 0%, $\tau^2$ = 0, $\chi^2_1$ = 0.87 ($p$ = 0.35)
5 10 15 20 25
Hospital stay (Period A, w/o outliers)

iv)

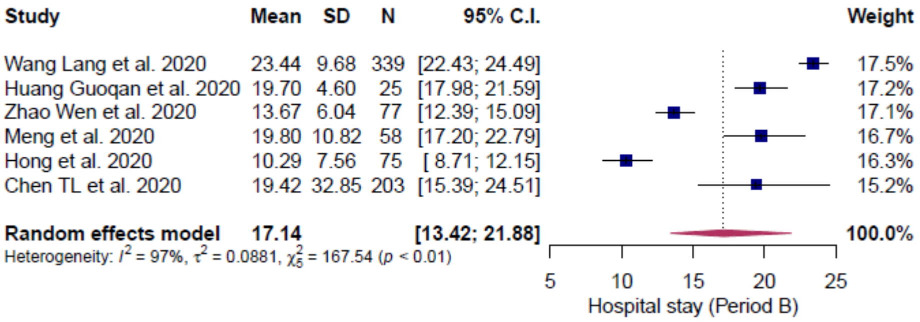

v)

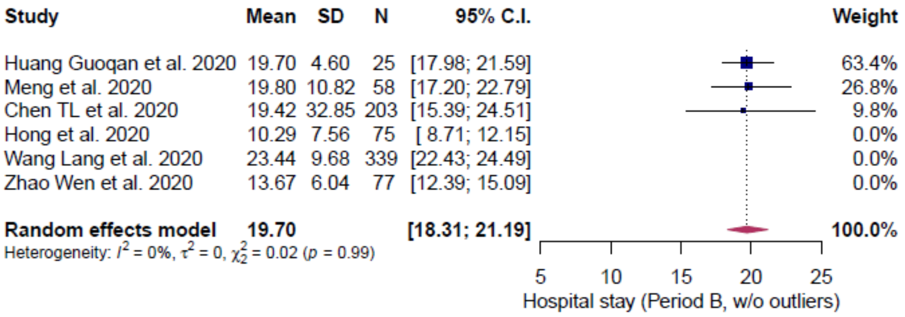

**Supplementary Figure S7.** Hospital stay length – all (general population). i) Results with all studies considered (both periods), ii) results of period A (all studies considered), iii) results of period A (outliers removed), iv) results of period B (all studies considered), v) results of period B (outliers removed). Period: A, includes patients admitted before Feb 12, 2020; B, includes patients admitted before and after Feb 12, 2020.

i)

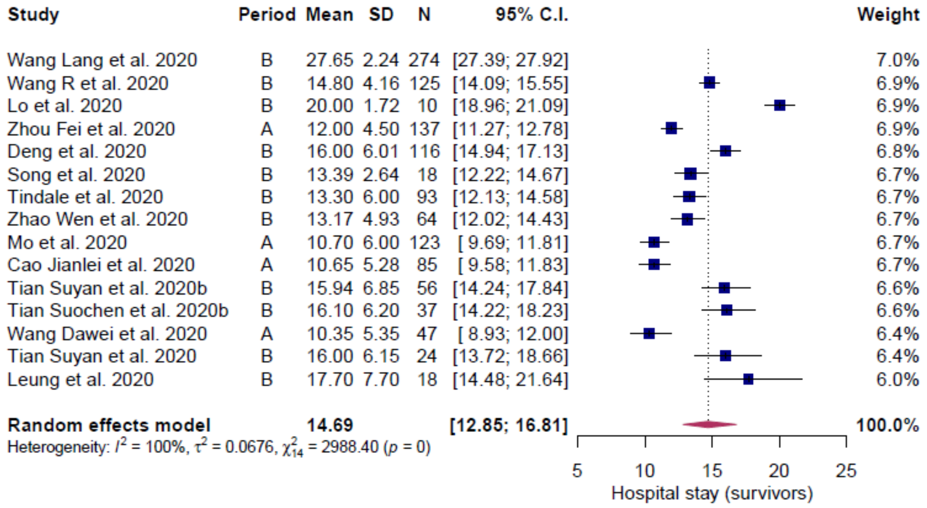

Study Period Mean SD N 95% C.I. Weight
Wang Lang et al. 2020 B 27.65 2.24 274 [27.39; 27.92] 7.0%
Wang R et al. 2020 B 14.80 4.16 125 [14.09; 15.55] 6.9%
Lo et al. 2020 B 20.00 1.72 10 [18.96; 21.09] 6.9%
Zhou Fei et al. 2020 A 12.00 4.50 137 [11.27; 12.78] 6.9%
Deng et al. 2020 B 16.00 6.01 116 [14.94; 17.13] 6.8%
Song et al. 2020 B 13.39 2.64 18 [12.22; 14.67] 6.7%
Tindale et al. 2020 B 13.30 6.00 93 [12.13; 14.58] 6.7%
Zhao Wen et al. 2020 B 13.17 4.93 64 [12.02; 14.43] 6.7%
Mo et al. 2020 A 10.70 6.00 123 [ 9.69; 11.81] 6.7%
Cao Jianlei et al. 2020 A 10.65 5.28 85 [ 9.58; 11.83] 6.7%
Tian Suyan et al. 2020b B 15.94 6.85 56 [14.24; 17.84] 6.6%
Tian Suochen et al. 2020b B 16.10 6.20 37 [14.22; 18.23] 6.6%
Wang Dawei et al. 2020 A 10.35 5.35 47 [ 8.93; 12.00] 6.4%
Tian Suyan et al. 2020 B 16.00 6.15 24 [13.72; 18.66] 6.4%
Leung et al. 2020 B 17.70 7.70 18 [14.48; 21.64] 6.0%
Random effects model 14.69 [12.85; 16.81] 100.0%
Heterogeneity: $I^2$ = 100%, $\tau^2$ = 0.0676, $\chi^2_{14}$ = 2988.40 ($p$ = 0)
5 10 15 20 25
Hospital stay (survivors)

ii)

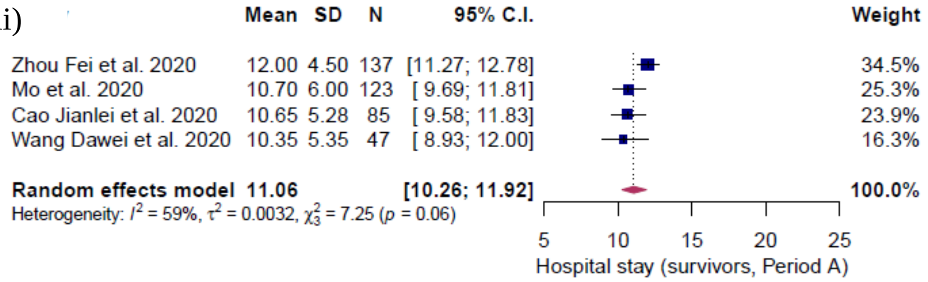

Mean SD N 95% C.I. Weight
Zhou Fei et al. 2020 12.00 4.50 137 [11.27; 12.78] 34.5%
Mo et al. 2020 10.70 6.00 123 [ 9.69; 11.81] 25.3%
Cao Jianlei et al. 2020 10.65 5.28 85 [ 9.58; 11.83] 23.9%
Wang Dawei et al. 2020 10.35 5.35 47 [ 8.93; 12.00] 16.3%
Random effects model 11.06 [10.26; 11.92] 100.0%
Heterogeneity: $I^2$ = 59%, $\tau^2$ = 0.0032, $\chi^2_3$ = 7.25 ($p$ = 0.06)
5 10 15 20 25
Hospital stay (survivors, Period A)

iii)

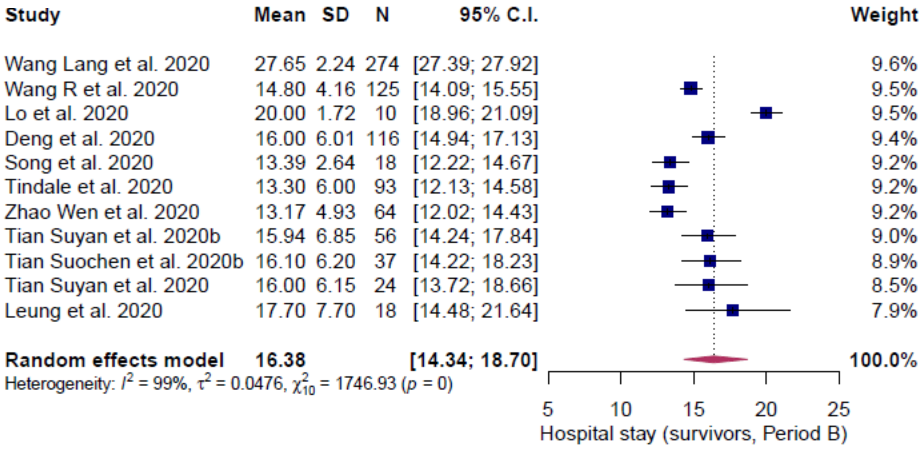

Study Mean SD N 95% C.I. Weight
Wang Lang et al. 2020 27.65 2.24 274 [27.39; 27.92] 9.6%
Wang R et al. 2020 14.80 4.16 125 [14.09; 15.55] 9.5%
Lo et al. 2020 20.00 1.72 10 [18.96; 21.09] 9.5%
Deng et al. 2020 16.00 6.01 116 [14.94; 17.13] 9.4%
Song et al. 2020 13.39 2.64 18 [12.22; 14.67] 9.2%
Tindale et al. 2020 13.30 6.00 93 [12.13; 14.58] 9.2%
Zhao Wen et al. 2020 13.17 4.93 64 [12.02; 14.43] 9.2%
Tian Suyan et al. 2020b 15.94 6.85 56 [14.24; 17.84] 9.0%
Tian Suochen et al. 2020b 16.10 6.20 37 [14.22; 18.23] 8.9%
Tian Suyan et al. 2020 16.00 6.15 24 [13.72; 18.66] 8.5%
Leung et al. 2020 17.70 7.70 18 [14.48; 21.64] 7.9%
Random effects model 16.38 [14.34; 18.70] 100.0%
Heterogeneity: $I^2$ = 99%, $\tau^2$ = 0.0476, $\chi^2_{10}$ = 1746.93 ($p$ = 0)
5 10 15 20 25
Hospital stay (survivors, Period B)

iv)

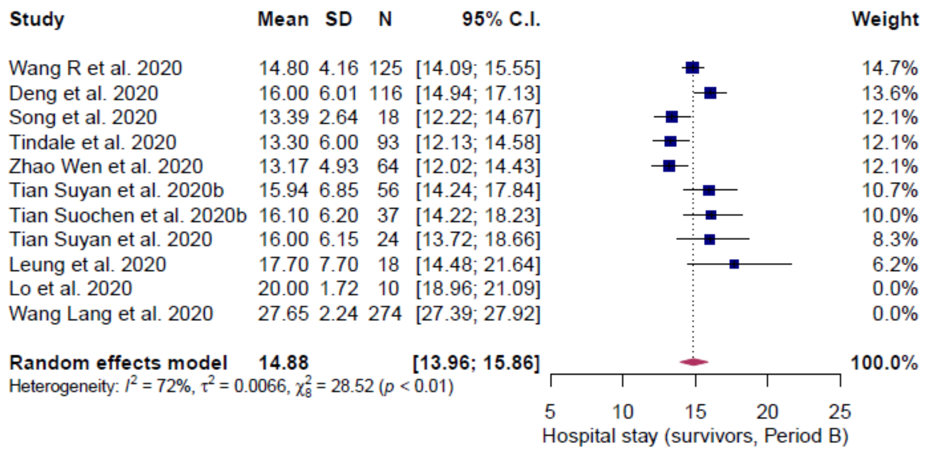

v)

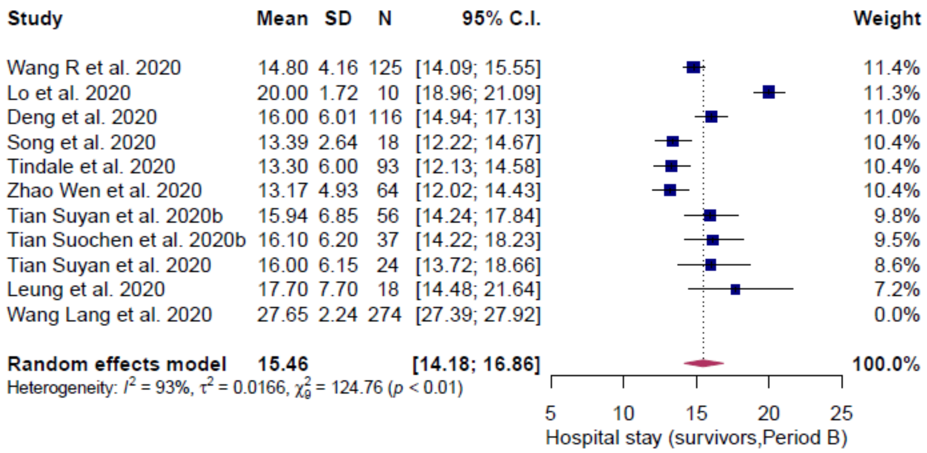

**Supplementary Figure S8.** Hospital stay – survivors (general population). i) Results with all studies considered (both periods), ii) results of period A (all studies considered), iii) results of period B (all studies considered), iv) results of period B (outliers removed) and v) results of period B (influencers removed). Period: A, includes patients admitted before Feb 12, 2020; B, includes patients admitted before and after Feb 12, 2020.

i)

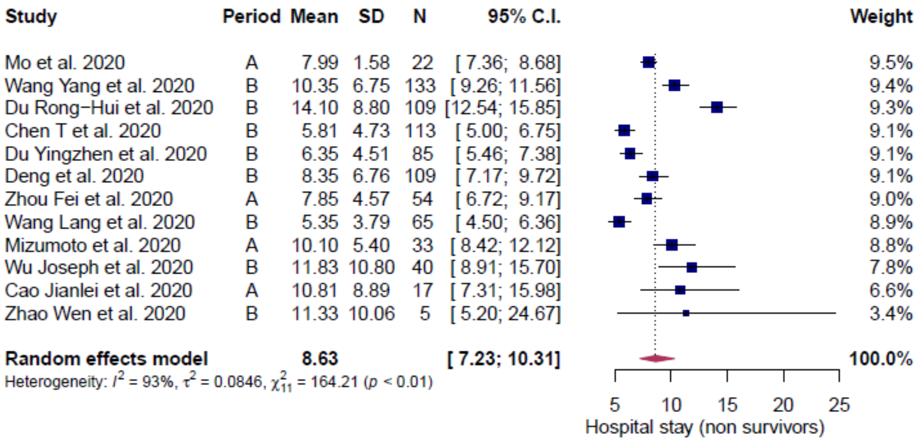
Study Period Mean SD N 95% C.I. Weight
Mo et al. 2020 A 7.99 1.58 22 [ 7.36; 8.68] 9.5%
Wang Yang et al. 2020 B 10.35 6.75 133 [ 9.26; 11.56] 9.4%
Du Rong−Hui et al. 2020 B 14.10 8.80 109 [12.54; 15.85] 9.3%
Chen T et al. 2020 B 5.81 4.73 113 [ 5.00; 6.75] 9.1%
Du Yingzhen et al. 2020 B 6.35 4.51 85 [ 5.46; 7.38] 9.1%
Deng et al. 2020 B 8.35 6.76 109 [ 7.17; 9.72] 9.1%
Zhou Fei et al. 2020 A 7.85 4.57 54 [ 6.72; 9.17] 9.0%
Wang Lang et al. 2020 B 5.35 3.79 65 [ 4.50; 6.36] 8.9%
Mizumoto et al. 2020 A 10.10 5.40 33 [ 8.42; 12.12] 8.8%
Wu Joseph et al. 2020 B 11.83 10.80 40 [ 8.91; 15.70] 7.8%
Cao Jianlei et al. 2020 A 10.81 8.89 17 [ 7.31; 15.98] 6.6%
Zhao Wen et al. 2020 B 11.33 10.06 5 [ 5.20; 24.67] 3.4%
Random effects model 8.63 [ 7.23; 10.31] 100.0%
Heterogeneity: $I^2 = 93\%$, $\tau^2 = 0.0846$, $\chi^2_{11} = 164.21$ ($p < 0.01$)
5 10 15 20 25
Hospital stay (non survivors)

ii)

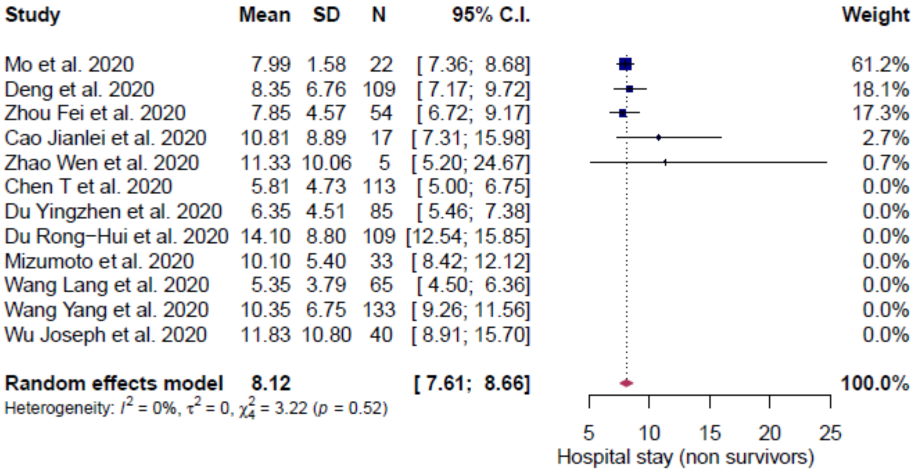
Study Mean SD N 95% C.I. Weight
Mo et al. 2020 7.99 1.58 22 [ 7.36; 8.68] 61.2%
Deng et al. 2020 8.35 6.76 109 [ 7.17; 9.72] 18.1%
Zhou Fei et al. 2020 7.85 4.57 54 [ 6.72; 9.17] 17.3%
Cao Jianlei et al. 2020 10.81 8.89 17 [ 7.31; 15.98] 2.7%
Zhao Wen et al. 2020 11.33 10.06 5 [ 5.20; 24.67] 0.7%
Chen T et al. 2020 5.81 4.73 113 [ 5.00; 6.75] 0.0%
Du Yingzhen et al. 2020 6.35 4.51 85 [ 5.46; 7.38] 0.0%
Du Rong−Hui et al. 2020 14.10 8.80 109 [12.54; 15.85] 0.0%
Mizumoto et al. 2020 10.10 5.40 33 [ 8.42; 12.12] 0.0%
Wang Lang et al. 2020 5.35 3.79 65 [ 4.50; 6.36] 0.0%
Wang Yang et al. 2020 10.35 6.75 133 [ 9.26; 11.56] 0.0%
Wu Joseph et al. 2020 11.83 10.80 40 [ 8.91; 15.70] 0.0%
Random effects model 8.12 [ 7.61; 8.66] 100.0%
Heterogeneity: $I^2 = 0\%$, $\tau^2 = 0$, $\chi^2_4 = 3.22$ ($p = 0.52$)
5 10 15 20 25
Hospital stay (non survivors)

iii)

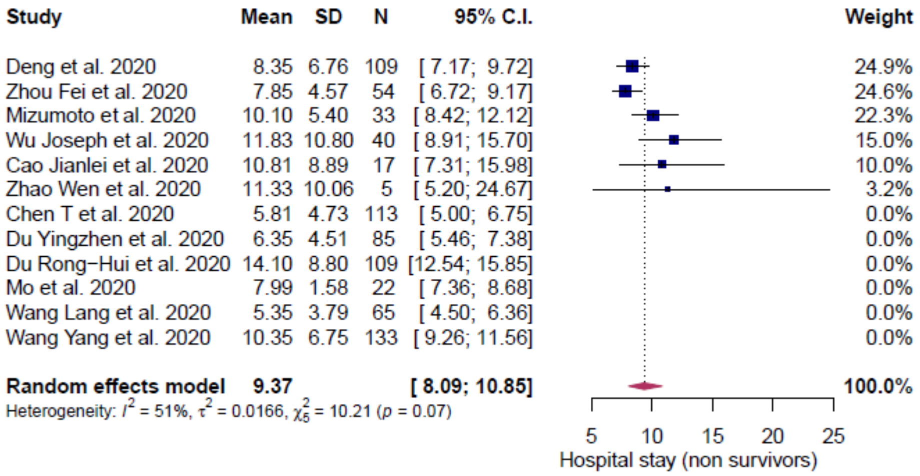

**Supplementary Figure S9.** Hospital stay length – non survivors (general population).i) Results with all studies considered, ii) results with outliers removed and iii) results with influencer removed. Period: A, includes patients admitted before Feb 12, 2020; B, includes patients admitted before and after Feb 12, 2020.

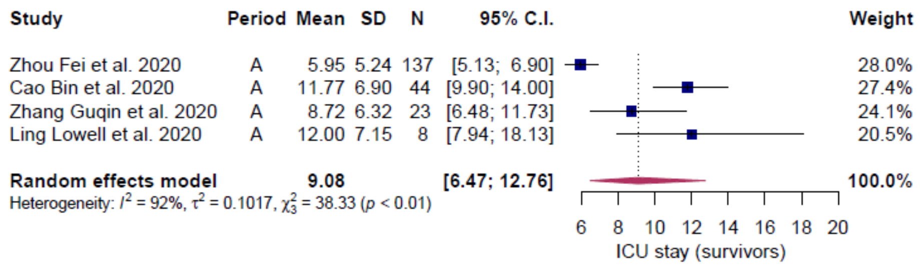

**Supplementary Figure S10.** ICU stay length – survivors (general population). Results with all studies considered. Period: A, includes patients admitted before Feb 12, 2020.

i)

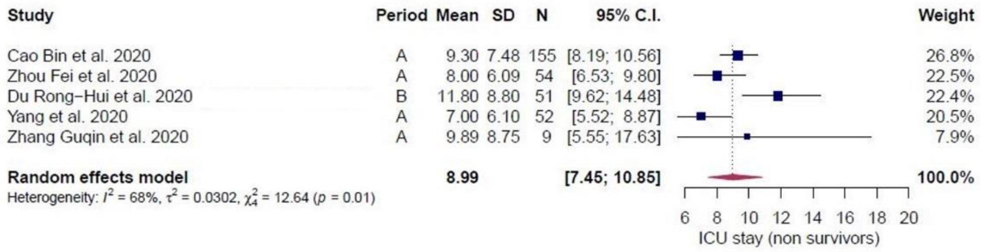

ii)

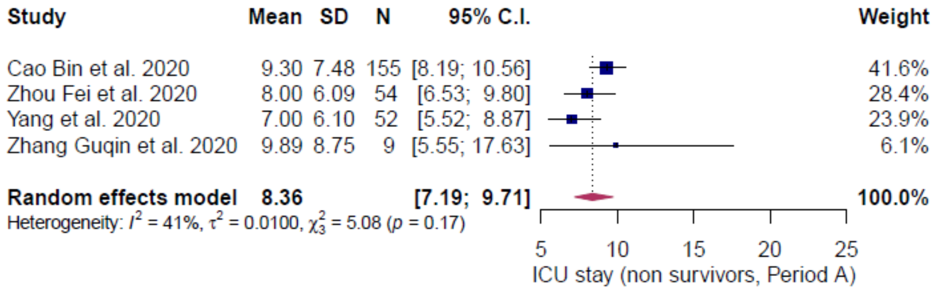

**Supplementary Figure S11.** ICU stay length – non survivors (general population). i) Results with all studies considered (both periods) and ii) results of results of period A (all studies considered). Period: A, includes patients admitted before Feb 12, 2020; B, includes patients admitted before and after Feb 12, 2020.

i)

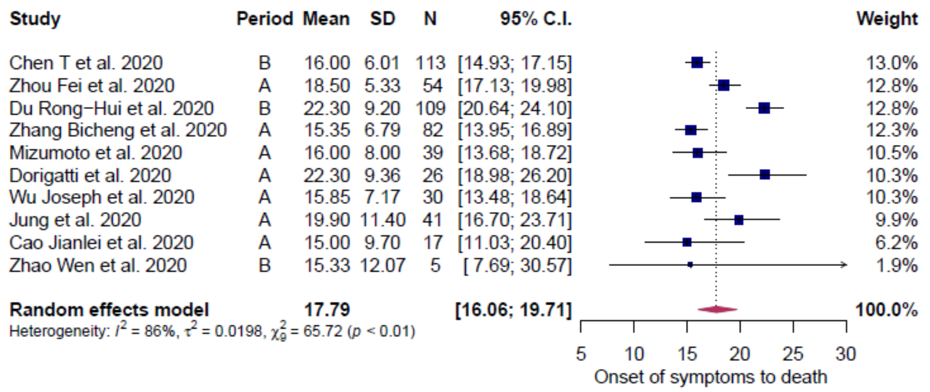

ii)

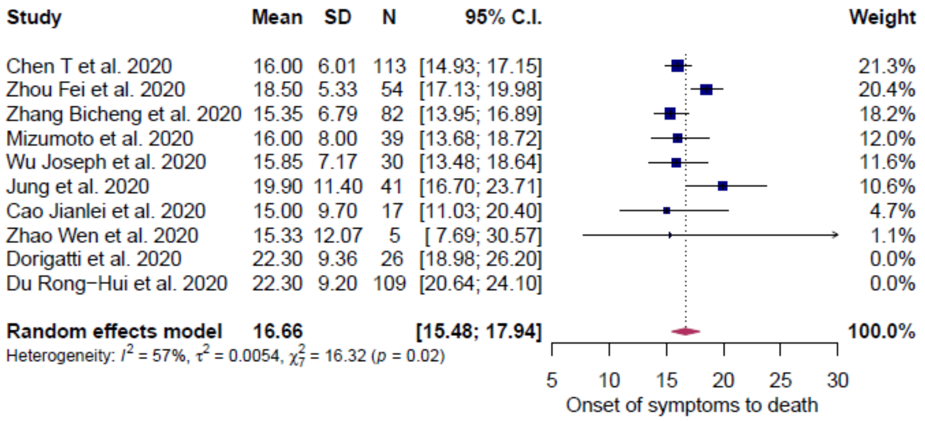

iii)

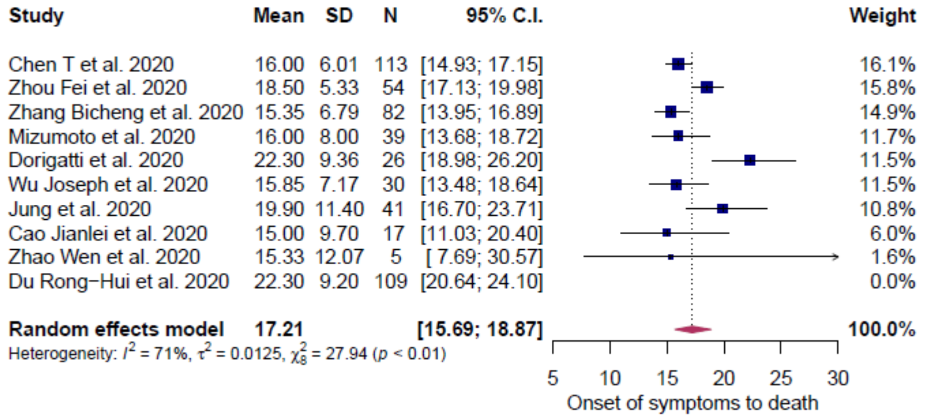

**Supplementary Figure S12.** Onset of symptoms to death (general population). i) Results with all studies considered, ii) results with outliers removed and iii) results with influencers removed. Period: A, includes patients admitted before Feb 12, 2020; B, includes patients admitted before and after Feb 12, 2020.

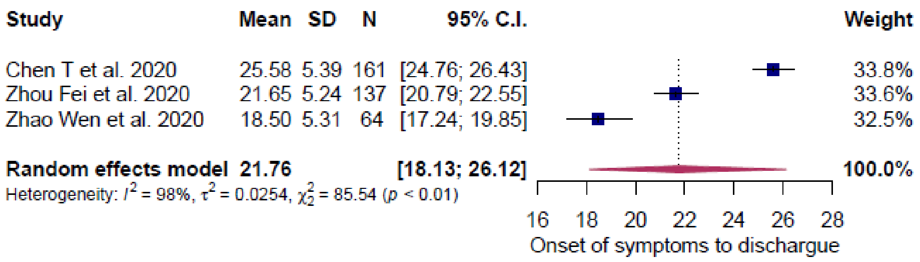

**Supplementary Figure S13.** Onset of symptoms to discharge (general population). Results with all studies considered.

i)

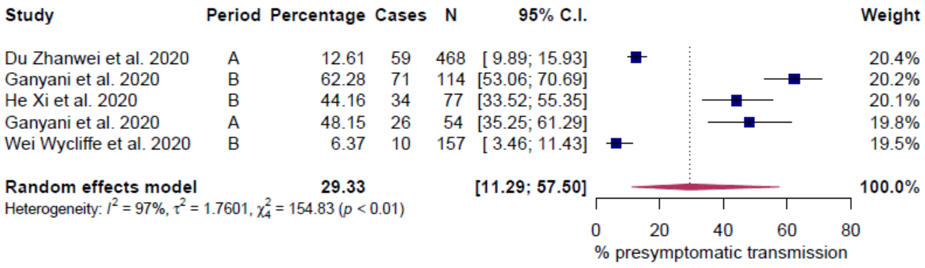

ii)

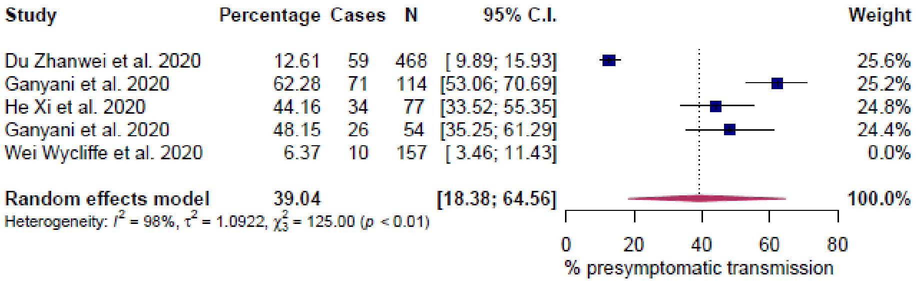

**Supplementary Figure S14.** Percentage of pre-symptomatic transmission (general population). i) Results with all studies considered and ii) results with influencers removed. Period: A, includes patients admitted before Feb 12, 2020; B, includes patients admitted before and after Feb 12, 2020.

i)

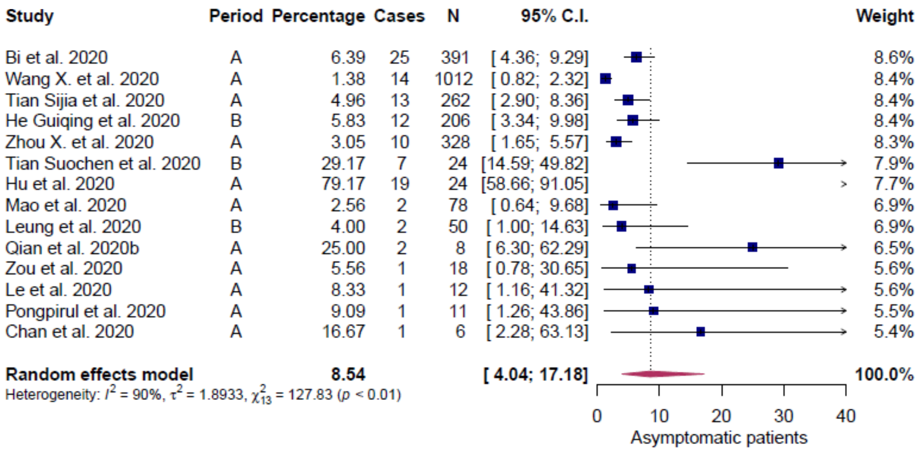
Study Period Percentage Cases N 95% C.I. Weight
Bi et al. 2020 A 6.39 25 391 [ 4.36; 9.29] 8.6%
Wang X. et al. 2020 A 1.38 14 1012 [ 0.82; 2.32] 8.4%
Tian Sijia et al. 2020 A 4.96 13 262 [ 2.90; 8.36] 8.4%
He Guiqing et al. 2020 B 5.83 12 206 [ 3.34; 9.98] 8.4%
Zhou X. et al. 2020 A 3.05 10 328 [ 1.65; 5.57] 8.3%
Tian Suochen et al. 2020 B 29.17 7 24 [14.59; 49.82] 7.9%
Hu et al. 2020 A 79.17 19 24 [58.66; 91.05] 7.7%
Mao et al. 2020 A 2.56 2 78 [ 0.64; 9.68] 6.9%
Leung et al. 2020 B 4.00 2 50 [ 1.00; 14.63] 6.9%
Qian et al. 2020b A 25.00 2 8 [ 6.30; 62.29] 6.5%
Zou et al. 2020 A 5.56 1 18 [ 0.78; 30.65] 5.6%
Le et al. 2020 A 8.33 1 12 [ 1.16; 41.32] 5.6%
Pongpirul et al. 2020 A 9.09 1 11 [ 1.26; 43.86] 5.5%
Chan et al. 2020 A 16.67 1 6 [ 2.28; 63.13] 5.4%
Random effects model 8.54 [ 4.04; 17.18] 100.0%
Heterogeneity: $I^2$ = 90%, $\tau^2$ = 1.8933, $\chi^2_{13}$ = 127.83 ($p$ < 0.01)
0 10 20 30 40
Asymptomatic patients

ii)

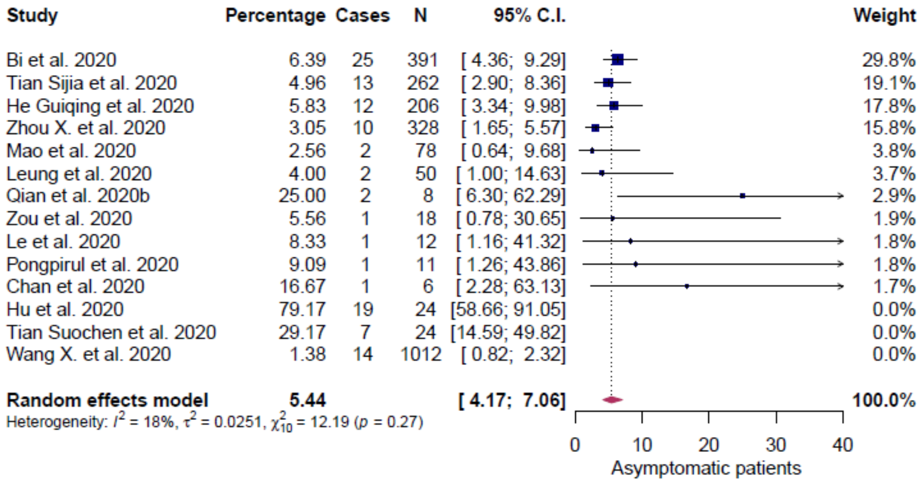
Study Percentage Cases N 95% C.I. Weight
Bi et al. 2020 6.39 25 391 [ 4.36; 9.29] 29.8%
Tian Sijia et al. 2020 4.96 13 262 [ 2.90; 8.36] 19.1%
He Guiqing et al. 2020 5.83 12 206 [ 3.34; 9.98] 17.8%
Zhou X. et al. 2020 3.05 10 328 [ 1.65; 5.57] 15.8%
Mao et al. 2020 2.56 2 78 [ 0.64; 9.68] 3.8%
Leung et al. 2020 4.00 2 50 [ 1.00; 14.63] 3.7%
Qian et al. 2020b 25.00 2 8 [ 6.30; 62.29] 2.9%
Zou et al. 2020 5.56 1 18 [ 0.78; 30.65] 1.9%
Le et al. 2020 8.33 1 12 [ 1.16; 41.32] 1.8%
Pongpirul et al. 2020 9.09 1 11 [ 1.26; 43.86] 1.8%
Chan et al. 2020 16.67 1 6 [ 2.28; 63.13] 1.7%
Hu et al. 2020 79.17 19 24 [58.66; 91.05] 0.0%
Tian Suochen et al. 2020 29.17 7 24 [14.59; 49.82] 0.0%
Wang X. et al. 2020 1.38 14 1012 [ 0.82; 2.32] 0.0%
Random effects model 5.44 [ 4.17; 7.06] 100.0%
Heterogeneity: $I^2$ = 18%, $\tau^2$ = 0.0251, $\chi^2_{10}$ = 12.19 ($p$ = 0.27)
0 10 20 30 40
Asymptomatic patients

iii)

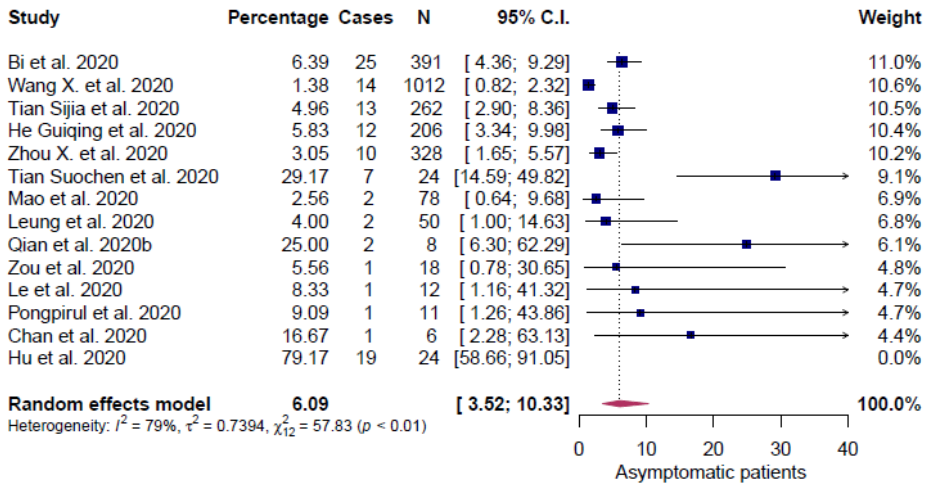

**Supplementary Figure S15.** Asymptomatic patients (general population). i) Results with all studies considered, ii) results with outliers removed and iii) results with influencers removed. Period: A, includes patients admitted before Feb 12, 2020; B, includes patients admitted before and after Jan Feb 12, 2020.

i)

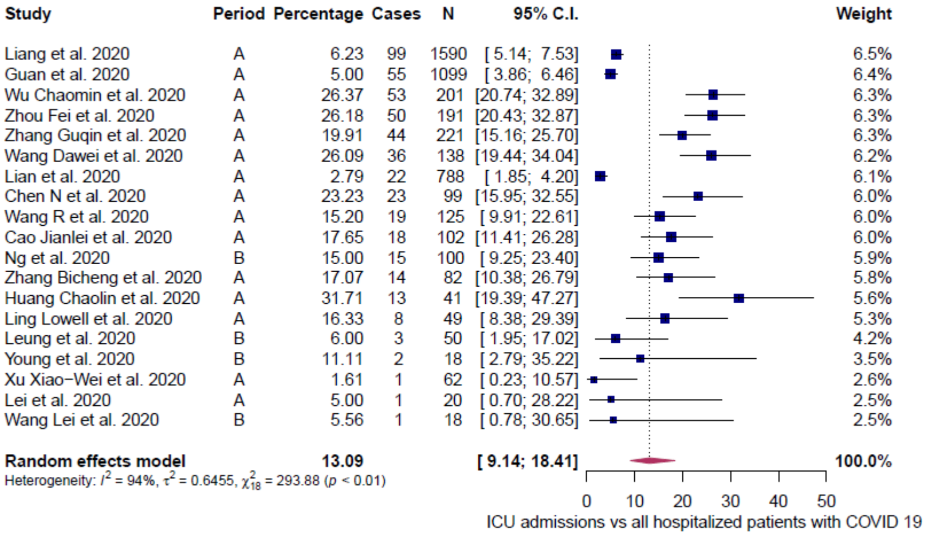

Study Period Percentage Cases N 95% C.I. Weight
Liang et al. 2020 A 6.23 99 1590 [ 5.14; 7.53] 6.5%
Guan et al. 2020 A 5.00 55 1099 [ 3.86; 6.46] 6.4%
Wu Chaomin et al. 2020 A 26.37 53 201 [20.74; 32.89] 6.3%
Zhou Fei et al. 2020 A 26.18 50 191 [20.43; 32.87] 6.3%
Zhang Guqin et al. 2020 A 19.91 44 221 [15.16; 25.70] 6.3%
Wang Dawei et al. 2020 A 26.09 36 138 [19.44; 34.04] 6.2%
Lian et al. 2020 A 2.79 22 788 [ 1.85; 4.20] 6.1%
Chen N et al. 2020 A 23.23 23 99 [15.95; 32.55] 6.0%
Wang R et al. 2020 A 15.20 19 125 [ 9.91; 22.61] 6.0%
Cao Jianlei et al. 2020 A 17.65 18 102 [11.41; 26.28] 6.0%
Ng et al. 2020 B 15.00 15 100 [ 9.25; 23.40] 5.9%
Zhang Bicheng et al. 2020 A 17.07 14 82 [10.38; 26.79] 5.8%
Huang Chaolin et al. 2020 A 31.71 13 41 [19.39; 47.27] 5.6%
Ling Lowell et al. 2020 A 16.33 8 49 [ 8.38; 29.39] 5.3%
Leung et al. 2020 B 6.00 3 50 [ 1.95; 17.02] 4.2%
Young et al. 2020 B 11.11 2 18 [ 2.79; 35.22] 3.5%
Xu Xiao−Wei et al. 2020 A 1.61 1 62 [ 0.23; 10.57] 2.6%
Lei et al. 2020 A 5.00 1 20 [ 0.70; 28.22] 2.5%
Wang Lei et al. 2020 B 5.56 1 18 [ 0.78; 30.65] 2.5%
Random effects model 13.09 [ 9.14; 18.41] 100.0%
Heterogeneity: $I^2 = 94\%$, $\tau^2 = 0.6455$, $\chi^2_{18} = 293.88$ ($p < 0.01$)
0 10 20 30 40 50
ICU admissions vs all hospitalized patients with COVID 19

ii)

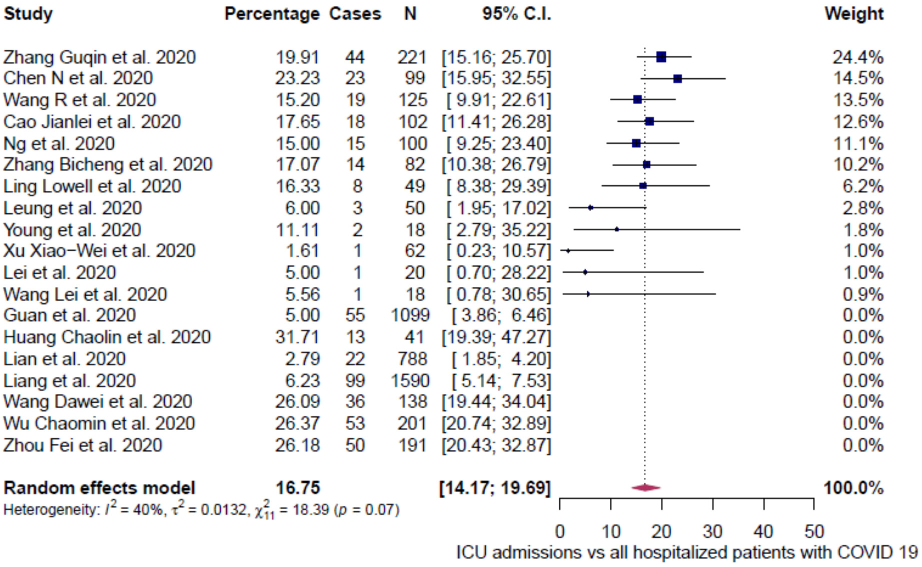

Study Percentage Cases N 95% C.I. Weight
Zhang Guqin et al. 2020 19.91 44 221 [15.16; 25.70] 24.4%
Chen N et al. 2020 23.23 23 99 [15.95; 32.55] 14.5%
Wang R et al. 2020 15.20 19 125 [ 9.91; 22.61] 13.5%
Cao Jianlei et al. 2020 17.65 18 102 [11.41; 26.28] 12.6%
Ng et al. 2020 15.00 15 100 [ 9.25; 23.40] 11.1%
Zhang Bicheng et al. 2020 17.07 14 82 [10.38; 26.79] 10.2%
Ling Lowell et al. 2020 16.33 8 49 [ 8.38; 29.39] 6.2%
Leung et al. 2020 6.00 3 50 [ 1.95; 17.02] 2.8%
Young et al. 2020 11.11 2 18 [ 2.79; 35.22] 1.8%
Xu Xiao−Wei et al. 2020 1.61 1 62 [ 0.23; 10.57] 1.0%
Lei et al. 2020 5.00 1 20 [ 0.70; 28.22] 1.0%
Wang Lei et al. 2020 5.56 1 18 [ 0.78; 30.65] 0.9%
Guan et al. 2020 5.00 55 1099 [ 3.86; 6.46] 0.0%
Huang Chaolin et al. 2020 31.71 13 41 [19.39; 47.27] 0.0%
Lian et al. 2020 2.79 22 788 [ 1.85; 4.20] 0.0%
Liang et al. 2020 6.23 99 1590 [ 5.14; 7.53] 0.0%
Wang Dawei et al. 2020 26.09 36 138 [19.44; 34.04] 0.0%
Wu Chaomin et al. 2020 26.37 53 201 [20.74; 32.89] 0.0%
Zhou Fei et al. 2020 26.18 50 191 [20.43; 32.87] 0.0%
Random effects model 16.75 [14.17; 19.69] 100.0%
Heterogeneity: $I^2 = 40\%$, $\tau^2 = 0.0132$, $\chi^2_{11} = 18.39$ ($p = 0.07$)
0 10 20 30 40 50
ICU admissions vs all hospitalized patients with COVID 19

iii)

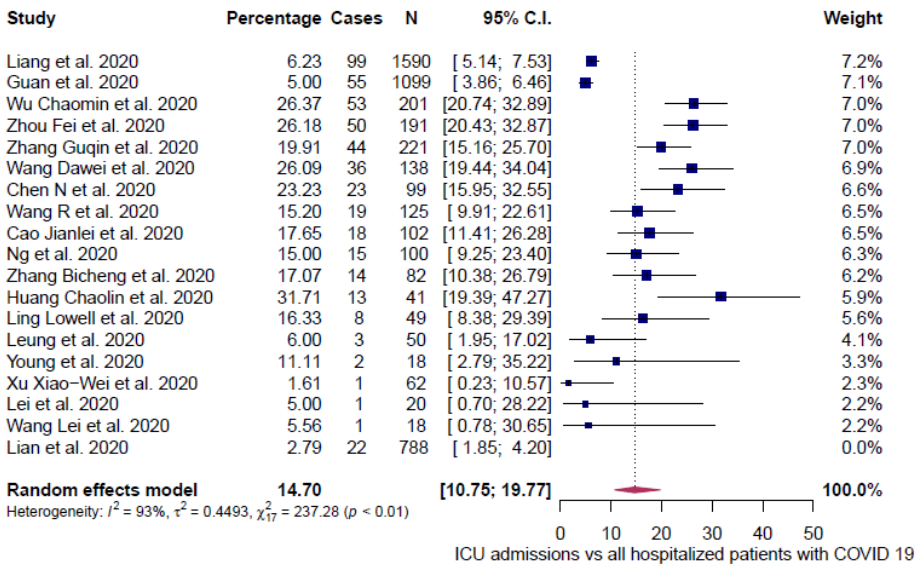

**Supplementary Figure S16.** ICU Admissions vs all hospital admissions (general population). i) Results with all studies considered, ii) results with outliers removed and iii) results with influencers removed. Period: A, includes patients admitted before Feb 12, 2020; B, includes patients admitted before and after Feb 12, 2020.

i)

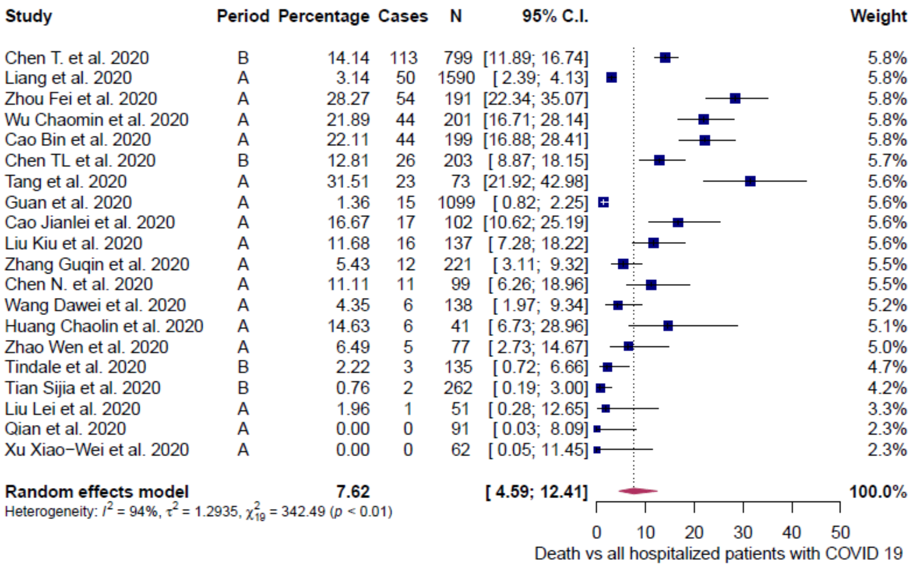

ii)

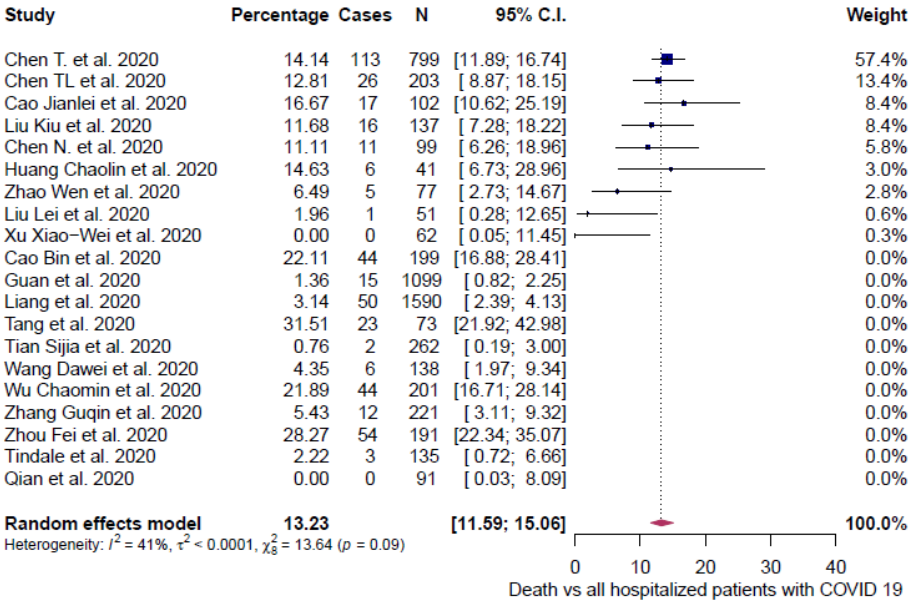

**Supplementary Figure S17.** Death vs all hospitalized patients (general population). i) Results with all studies considered and ii) results with outliers removed. Period: A, includes patients admitted before Feb 12, 2020; B, includes patients admitted before and after Feb 12, 2020.

i)

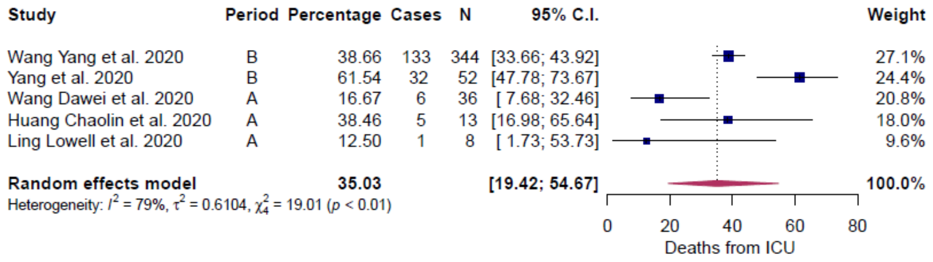

ii)

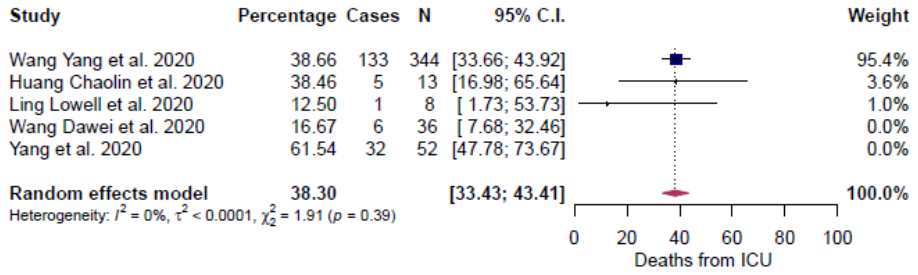

iii)

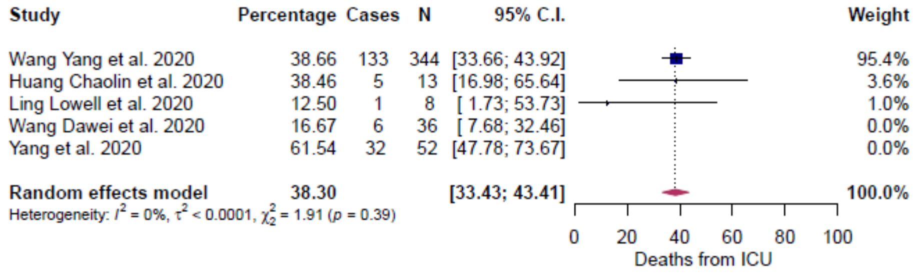

**Supplementary Figure S18.** Deaths from ICU (general population). i) Results with all studies considered ii) results with outliers removed and iii) results with influencers removed. Period: A, includes patients admitted before Feb 12, 2020; B, includes patients admitted before and after Feb 12, 2020.

i)

| Study | Period | Percentage | Cases | N | 95% C.I. | Weight |
|---|---|---|---|---|---|---|
| Lian et al. 2020 | A | 48.48 | 382 | 788 | [45.00; 51.97] | 4.3% |
| Chen T. et al. 2020 | B | 20.15 | 161 | 799 | [17.51; 23.08] | 4.3% |
| Guan et al. 2020 | A | 5.82 | 64 | 1099 | [ 4.58; 7.37] | 4.3% |
| Zhou Fei et al. 2020 | A | 71.73 | 137 | 191 | [64.93; 77.66] | 4.3% |
| Tian Sijia et al. 2020 | A | 17.18 | 45 | 262 | [13.08; 22.23] | 4.3% |
| Zhang Guqin et al. 2020 | A | 19.00 | 42 | 221 | [14.36; 24.72] | 4.3% |
| Wang Dawei et al. 2020 | A | 34.06 | 47 | 138 | [26.64; 42.34] | 4.3% |
| Liu Kiu et al. 2021 | A | 32.12 | 44 | 137 | [24.84; 40.38] | 4.3% |
| Wang R et al. 2020 | B | 37.60 | 47 | 125 | [29.56; 46.39] | 4.3% |
| Chen TL et al. 2020 | B | 87.19 | 177 | 203 | [81.85; 91.13] | 4.2% |
| Chen N. et al. 2020 | A | 31.31 | 31 | 99 | [22.97; 41.08] | 4.2% |
| Tindale et al. 2020 | B | 66.67 | 62 | 93 | [56.51; 75.48] | 4.2% |
| Qian et al. 2020 | A | 34.07 | 31 | 91 | [25.09; 44.35] | 4.2% |
| Tang et al. 2020 | A | 35.62 | 26 | 73 | [25.52; 47.18] | 4.2% |
| Wu Jian et al. 2020 | B | 23.75 | 19 | 80 | [15.69; 34.27] | 4.2% |
| Cao Jianlei et al. 2020 | A | 83.33 | 85 | 102 | [74.81; 89.38] | 4.2% |
| Leung et al. 2020 | B | 40.00 | 20 | 50 | [27.46; 54.00] | 4.1% |
| Zhao Wen et al. 2020 | A | 83.12 | 64 | 77 | [73.06; 89.94] | 4.1% |
| Huang Chaolin et al. 2020 | A | 68.29 | 28 | 41 | [52.73; 80.61] | 4.1% |
| Young et al. 2020 | B | 44.44 | 8 | 18 | [24.00; 66.96] | 3.9% |
| Lei et al. 2020 | A | 70.00 | 14 | 20 | [47.28; 85.86] | 3.8% |
| Wang Lei et al. 2020 | B | 33.33 | 6 | 18 | [15.80; 57.12] | 3.8% |
| Huang Lei et al. 2020 | B | 50.00 | 4 | 8 | [20.01; 79.99] | 3.4% |
| Liu Lei et al. 2020 | A | 98.04 | 50 | 51 | [87.35; 99.72] | 2.8% |
| Tian Suchen et al. 2020 | B | 100.00 | 56 | 56 | [87.47; 99.95] | 2.0% |
| **Random effects model** | | **49.34** | | | **[36.01; 62.76]** | **100.0%** |

Heterogeneity: $I^2 = 97\%$, $\tau^2 = 1.8064$, $\chi^2_{24} = 939.39$ ($p < 0.01$)

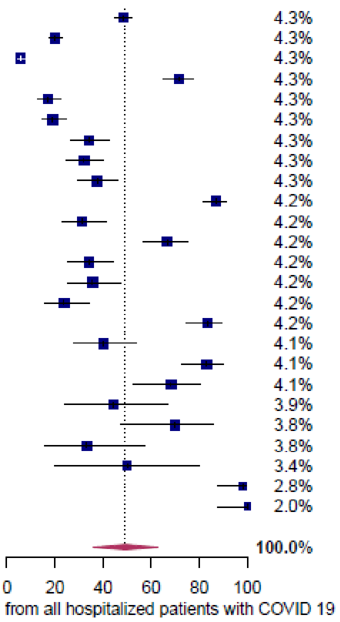

ii)

| Study | Percentage | Cases | N | 95% C.I. | Weight |
|---|---|---|---|---|---|
| Wang Dawei et al. 2020 | 34.06 | 47 | 138 | [26.64; 42.34] | 18.1% |
| Liu Kiu et al. 2021 | 32.12 | 44 | 137 | [24.84; 40.38] | 17.5% |
| Wang R et al. 2020 | 37.60 | 47 | 125 | [29.56; 46.39] | 17.1% |
| Chen N. et al. 2020 | 31.31 | 31 | 99 | [22.97; 41.08] | 12.4% |
| Qian et al. 2020 | 34.07 | 31 | 91 | [25.09; 44.35] | 11.9% |
| Tang et al. 2020 | 35.62 | 26 | 73 | [25.52; 47.18] | 9.8% |
| Leung et al. 2020 | 40.00 | 20 | 50 | [27.46; 54.00] | 7.0% |
| Young et al. 2020 | 44.44 | 8 | 18 | [24.00; 66.96] | 2.6% |
| Wang Lei et al. 2020 | 33.33 | 6 | 18 | [15.80; 57.12] | 2.3% |
| Huang Lei et al. 2020 | 50.00 | 4 | 8 | [20.01; 79.99] | 1.2% |
| Cao Jianlei et al. 2020 | 83.33 | 85 | 102 | [74.81; 89.38] | 0.0% |
| Chen TL et al. 2020 | 87.19 | 177 | 203 | [81.85; 91.13] | 0.0% |
| Chen T. et al. 2020 | 20.15 | 161 | 799 | [17.51; 23.08] | 0.0% |
| Guan et al. 2020 | 5.82 | 64 | 1099 | [ 4.58; 7.37] | 0.0% |
| Huang Chaolin et al. 2020 | 68.29 | 28 | 41 | [52.73; 80.61] | 0.0% |
| Lei et al. 2020 | 70.00 | 14 | 20 | [47.28; 85.86] | 0.0% |
| Lian et al. 2020 | 48.48 | 382 | 788 | [45.00; 51.97] | 0.0% |
| Tian Sijia et al. 2020 | 17.18 | 45 | 262 | [13.08; 22.23] | 0.0% |
| Tindale et al. 2020 | 66.67 | 62 | 93 | [56.51; 75.48] | 0.0% |
| Wu Jian et al. 2020 | 23.75 | 19 | 80 | [15.69; 34.27] | 0.0% |
| Zhang Guqin et al. 2020 | 19.00 | 42 | 221 | [14.36; 24.72] | 0.0% |
| Zhao Wen et al. 2020 | 83.12 | 64 | 77 | [73.06; 89.94] | 0.0% |
| Zhou Fei et al. 2020 | 71.73 | 137 | 191 | [64.93; 77.66] | 0.0% |
| Liu Lei et al. 2020 | 98.04 | 50 | 51 | [87.35; 99.72] | 0.0% |
| Tian Suchen et al. 2020 | 100.00 | 56 | 56 | [87.47; 99.95] | 0.0% |
| **Random effects model** | **34.93** | | | **[31.61; 38.41]** | **100.0%** |

Heterogeneity: $I^2 = 0\%$, $\tau^2 = 0$, $\chi^2_9 = 3.59$ ($p = 0.94$)

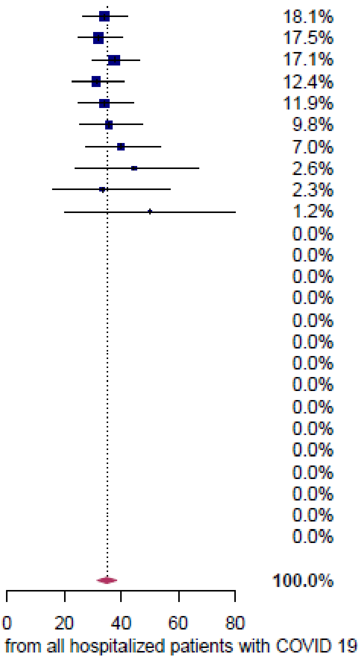

**Supplementary Figure S19.** Discharged vs all hospitalized patients (general population). i) Results with all studies considered and ii) results with outliers removed. Period: A, includes patients admitted before Feb 12, 2020; B, includes patients admitted before and after Feb 12, 2020.

i)

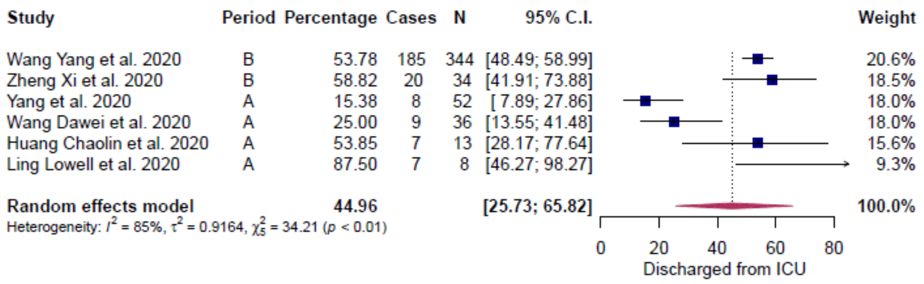

ii)

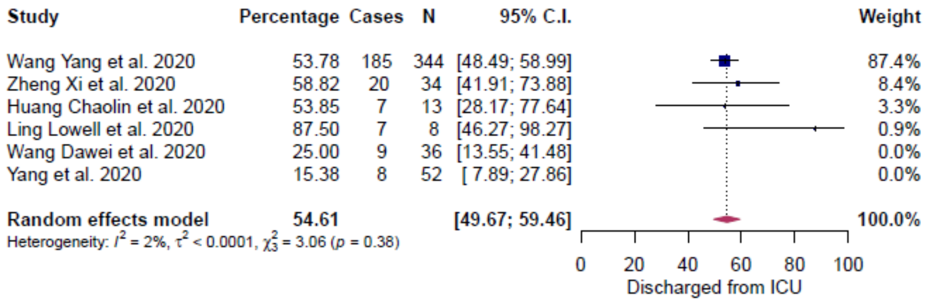

**Supplementary Figure S20.** Discharged from ICU (general population). i) Results with all studies considered and ii) results with outliers removed. Period: A, includes patients admitted before Feb 12, 2020; B, includes patients admitted before and after Jan Feb 12, 2020.

i)

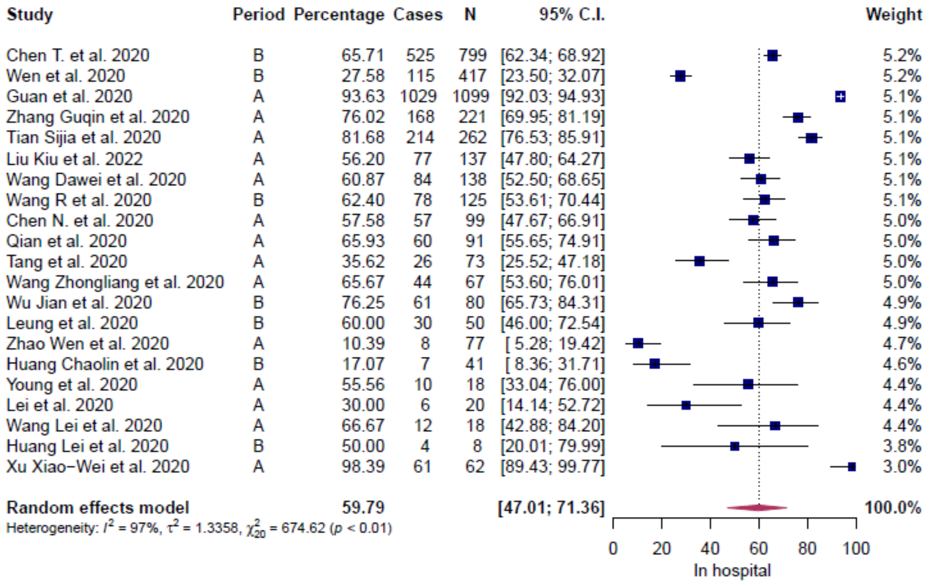

ii)

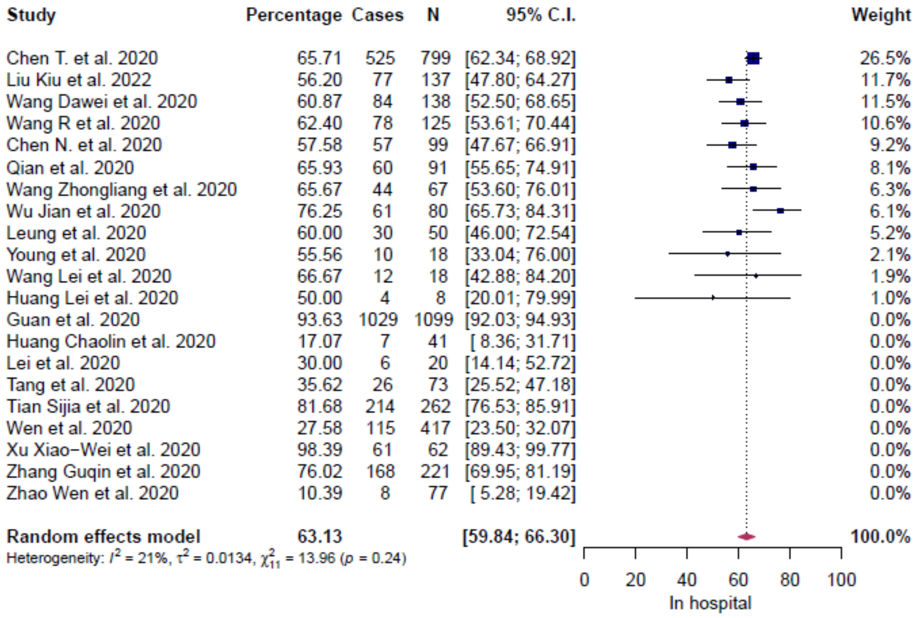

**Supplementary Figure S21.** In hospital (general population). i) Results with all studies considered and ii) results with outliers removed. Period: A, includes patients admitted before Feb 12, 2020; B, includes patients admitted before and after Feb 12. 2020.

i)

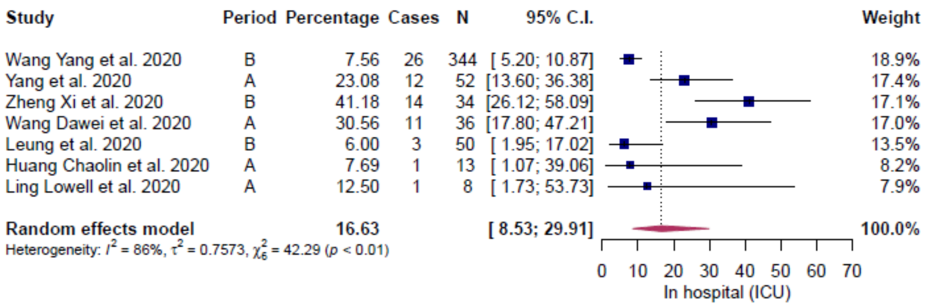

ii)

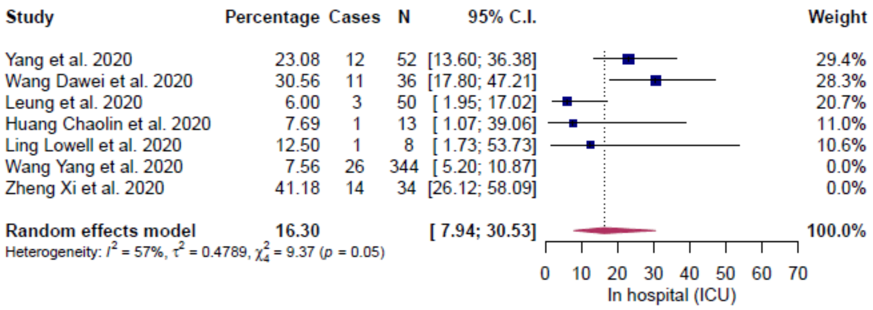

**Supplementary Figure S22.** In hospital (ICU) (general population). i) Results with all studies considered and ii) results with outliers removed. Period: A, includes patients admitted before Feb 12, 2020; B, includes patients admitted before and after Feb 12, 2020.

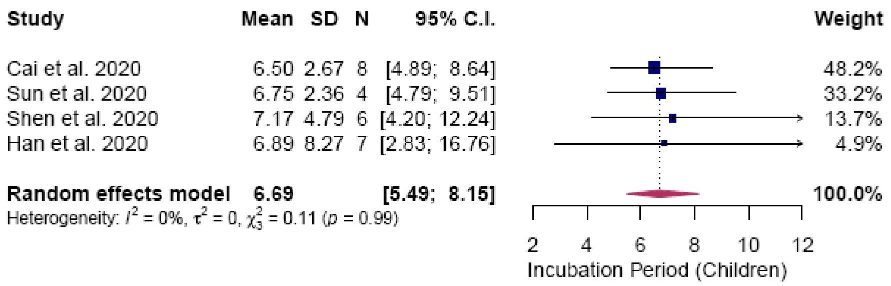

**Supplementary Figure S23.** Incubation period (children). Results with all studies considered.

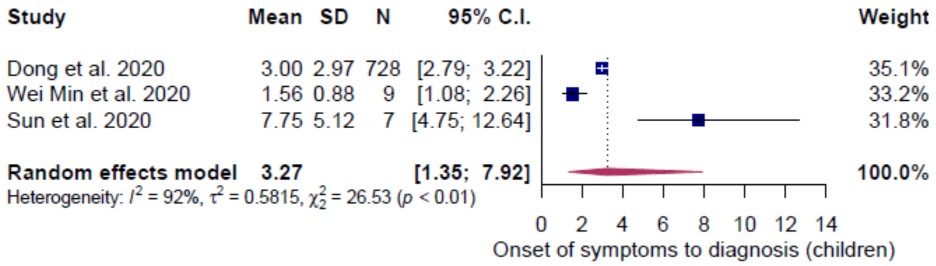

**Supplementary Figure S24.** Onset of symptom to diagnosis (children). Results with all studies considered.

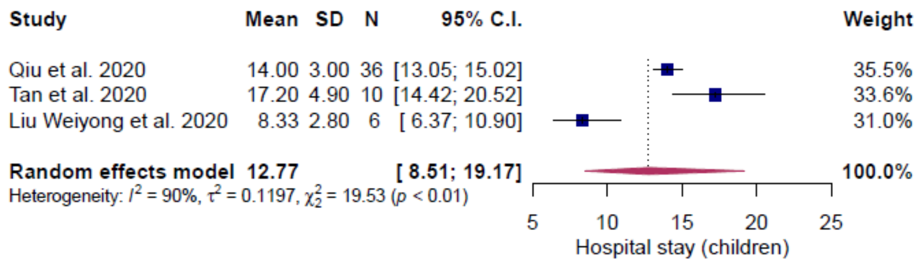

**Supplementary Figure S25.** Hospital stay (children). Results with all studies considered.

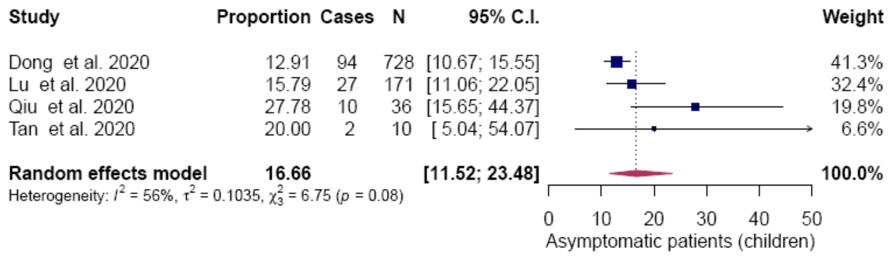

**Supplementary Figure S26.** Asymptomatic patients (children). Results with all studies considered.

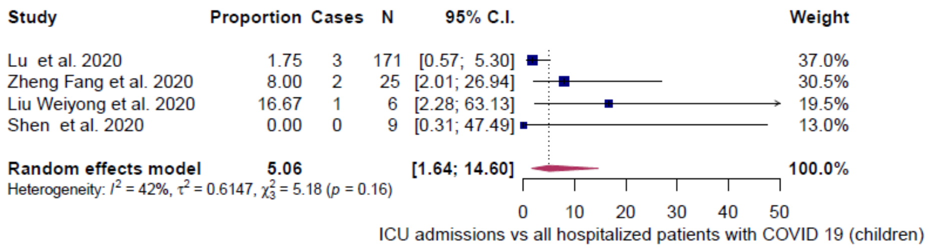

**Supplementary Figure S27.** ICU admissions vs all hospital admissions (children). Results with all studies considered.

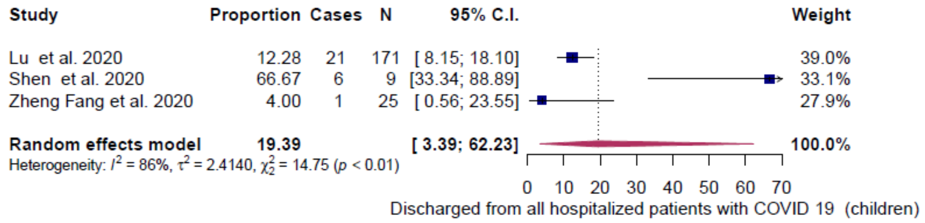

**Supplementary Figure S28.** Discharged vs all hospitalized patients (children). Results with all studies considered.

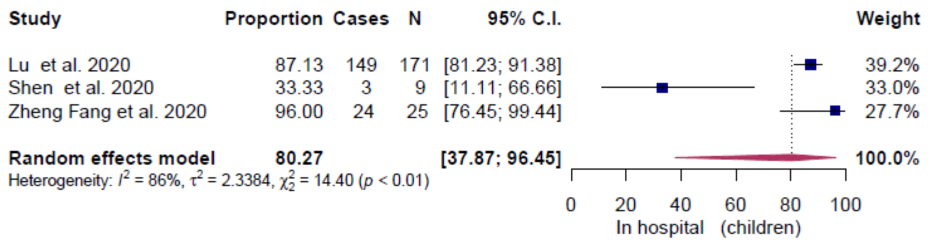

**Supplementary Figure S29.** In hospital (children). Results with all studies considered.
