## Supplementary material for "Scoping review and meta-analysis of COVID-19 epidemiological parameters for modeling from early Asian studies": Meta-analysis report

### META-ANALYSIS REPORT
### (WITH OUTLIERS AND INFLUENCERS DIAGNOSIS)

Details on meta-analytical method (common to all):
- Inverse variance method
- Restricted maximum-likelihood estimator for tau^2
- Jackson method for confidence interval of tau^2 and tau
- Logit transformation

#### GENERAL POPULATION: Meta-analysis of single means

---------------------------
##### Serial Interval
---------------------------

Number of studies combined: k = 7

| | mean | 95%-CI |
|---|---|---|
| Fixed effect model | 5.7299 | [5.6236; 5.8383] |
| **Random effects model** | **4.9658** | **[3.8751; 6.3635]** |

Quantifying heterogeneity:
tau^2 = 0.0895 [0.0304; 1.1795]; tau = 0.2991 [0.1743; 1.0860];
I^2 = 90.6% [83.1%; 94.7%]; H = 3.25 [2.43; 4.35]

Test of heterogeneity:

| Q | d.f. | p-value |
|---|---|---|
| 63.55 | 6 | < 0.0001 |

**Meta-regression with Period as a Categorical Moderator:**

Multivariate Meta-Analysis Model (k = 7; method: REML)

Variance Components:

| | estim | sqrt | nlvls | fixed | factor |
|---|---|---|---|---|---|
| sigma^2 | 1.816 | 1.347 | 2 | no | period |

Test for Residual Heterogeneity:
QE(df = 5) = 75.426, p-val < .001

Test of Moderators (coefficient 2):
**QM(df = 1) = 0.565, p-val = 0.452**

Model Results:

| | estimate | se | zval | pval | ci.lb | ci.ub | |
|---|---|---|---|---|---|---|---|
| intrcpt | 4.306 | 1.363 | 3.160 | 0.002 | 1.635 | 6.978 | ** |
| periodB | 1.442 | 1.917 | 0.752 | 0.452 | -2.316 | 5.200 | |

---
Signif. codes: 0 '***' 0.001 '**' 0.01 '*' 0.05 '.' 0.1 ' ' 1

➔ There are no significant differences between the two subsets.

**Outliers :**
"2", "4"

**Results with outliers removed :**

Number of studies combined: k = 5

| | mean | 95%-CI |
|---|---|---|
| Fixed effect model | 5.8005 | [5.6912; 5.9119] |

**Random effects model 5.8005 [5.6879; 5.9154]**

Quantifying heterogeneity:

tau^2 < 0.0001 [0.0000; 0.2126]; tau = 0.0025 [0.0000; 0.4611];
I^2 = 17.9% [0.0%; 82.9%]; H = 1.10 [1.00; 2.42]

Test of heterogeneity:

```
  Q d.f.  p-value
4.87    4  0.3004
```

**Influencers:**

"4"

**Results with influencers removed:**

Number of studies combined: k = 6

| | mean | 95%-CI |
|---|---|---|
| Fixed effect model | 5.7350 | [5.6286; 5.8435] |
| **Random effects model** | **5.3846** | **[4.5120; 6.4259]** |

Quantifying heterogeneity:

tau^2 = 0.0365 [0.0087; 0.2766]; tau = 0.1911 [0.0933; 0.5260];
I^2 = 90.2% [81.3%; 94.8%]; H = 3.19 [2.31; 4.40]

Test of heterogeneity:

```
  Q d.f.  p-value
50.86   5 < 0.0001
```

---------------------------

#### Incubation Period

---------------------------

Number of studies combined: k = 16

| | mean | 95%-CI |
|---|---|---|
| Fixed effect model | 5.5017 | [5.3554; 5.6521] |
| Random effects model | 5.6085 | [4.7425; 6.6328] |

Quantifying heterogeneity:

tau^2 = 0.1050 [0.0536; 0.3035];
tau = 0.3241 [0.2315; 0.5509];
I^2 = 96.0% [94.7%; 97.0%];
H = 5.00 [4.33; 5.77]

Test of heterogeneity:

```
   Q      d.f.  p-value
374.70   15   < 0.0001
```

**Meta-regression with period as a categorical moderator**

Multivariate Meta-Analysis Model (k = 16; method: REML)

Variance Components:

| | estim | sqrt | nlvls | fixed | factor |
|---|---|---|---|---|---|
| sigma^2 | 1.208 | 1.099 | 2 | no | period |

Test for Residual Heterogeneity:

QE(df = 14) = 244.819, p-val < .001

Test of Moderators (coefficient 2):

QM(df = 1) = 0.726, p-val = 0.394

Model Results:

| | estimate | se | zval | pval | ci.lb | ci.ub | |
|---|---|---|---|---|---|---|---|
| intrcpt | 4.600 | 1.102 | 4.173 | <.001 | 2.439 | 6.760 | *** |
| periodB | 1.331 | 1.562 | 0.852 | 0.394 | -1.730 | 4.392 | |

```
---
Signif. codes:  0 '***' 0.001 '**' 0.01 '*' 0.05 '.' 0.1 ' ' 1
```

Identified outliers

-------------------------

"1", "2", "3", "8", "9", "10", "11", "14"

Results with outliers removed

-----------------------------

Number of studies combined: k = 8

```
                      mean      95%-CI
Fixed effect model    5.1334 [4.8555; 5.4272]
Random effects model 5.2957 [4.8170; 5.8220]

Quantifying heterogeneity:
 tau^2 = 0.0075 [0.0000; 0.1419]; tau = 0.0864 [0.0000; 0.3768];
 I^2 = 52.2% [0.0%; 78.5%]; H = 1.45 [1.00; 2.16]

Test of heterogeneity:
    Q d.f. p-value
 14.64    7  0.0409
```

Identified influencers

-------------------------

"14"

Results with influencers removed

-----------------------------

Number of studies combined: k = 15

```
                      mean      95%-CI
Fixed effect model     5.3738 [5.2286; 5.5229]
Random effects model   5.3453 [4.6568; 6.1356]

Quantifying heterogeneity:
        tau^2 = 0.0625 [0.0309; 0.2301]; tau = 0.2499 [0.1759; 0.4797];
        I^2 = 95.0% [93.2%; 96.4%]; H = 4.48 [3.83; 5.26]
Test of heterogeneity:
          Q    d.f.    p-value
        281.51   14    < 0.0001
```

---------------------------

#### Onset of symptoms to hospital admission

---------------------------

Number of studies combined: k = 27

```
                     mean      95%-CI
Fixed effect model    8.5825   [8.4419; 8.7255]
Random effects model 6.2879   [5.3162; 7.4372]

Quantifying heterogeneity:
        tau^2 = 0.1838 [0.1071; 0.3401]; tau = 0.4287 [0.3272; 0.5832];
        I^2 = 98.3% [98.0%; 98.6%]; H = 7.72 [7.11; 8.39]
Test of heterogeneity:
          Q d.f.  p-value
        1551.23   26 < 0.0001
```

**Meta-regression with period as a categorical moderator**

```
Multivariate Meta-Analysis Model (k = 27; method: REML)

Variance Components:
              estim   sqrt  nlvls  fixed  factor
   sigma^2    2.331  1.527      2     no  period

Test for Residual Heterogeneity: QE(df = 25) = 2295.747, p-val < .001
Test of Moderators (coefficient 2): QM(df = 1) = 0.834, p-val = 0.361

Model Results:
             estimate     se   zval   pval    ci.lb   ci.ub
   intrcpt      6.263  1.528  4.098  <.001    3.268   9.259  ***
   periodB      1.977  2.165  0.913  0.361   -2.266   6.221

---
Signif. codes:  0 '***' 0.001 '**' 0.01 '*' 0.05 '.' 0.1 ' ' 1
```

Identified outliers
-------------------------
"1", "2", "3", "4", "5", "7", "12", "14", "17", "18", "19", "21", "23", "24", "25", "26", "27"

Results with outliers removed
-----------------------------
Number of studies combined: k = 10

```
                         mean          95%-CI
Fixed effect model     6.5457 [6.2545; 6.8504]
Random effects model 6.5079 [6.1175; 6.9233]

Quantifying heterogeneity:
   tau^2 = 0.0024 [0.0000; 0.0180]; tau = 0.0488 [0.0000; 0.1343];
   I^2 = 11.9% [0.0%; 52.9%]; H = 1.07 [1.00; 1.46]

Test of heterogeneity:
       Q d.f. p-value
   10.22    9  0.3329
```

No influencers detected
-----------------

---------------------------

#### Onset of symptoms to hospital admission (survivors)

---------------------------

Number of studies combined: k = 6

Number of studies combined: k = 6

```
                         mean          95%-CI
Fixed effect model     8.0998 [7.8300; 8.3790]
Random effects model 7.1948 [5.3940; 9.5968]

Quantifying heterogeneity:
   tau^2 = 0.1249 [0.0453; 0.8275]; tau = 0.3534 [0.2129; 0.9097];
   I^2 = 98.2% [97.4%; 98.8%]; H = 7.48 [6.18; 9.06]

Test of heterogeneity:
        Q d.f.  p-value
   279.86    5 < 0.0001
```

**Meta-regression with period as a categorical moderator**

Multivariate Meta-Analysis Model (k = 6; method: REML)

```
Variance Components:
            estim   sqrt  nlvls  fixed  factor
sigma^2    1.039  1.019      2     no  period

Test for Residual Heterogeneity: QE(df = 4) = 106.507, p-val < .001
Test of Moderators (coefficient 2): QM(df = 1) = 7.641, p-val = 0.006

Model Results:
         estimate    se    zval   pval   ci.lb   ci.ub
intrcpt    9.907  1.047   9.464  <.001   7.855  11.958  ***
periodB   -4.060  1.469  -2.764  0.006  -6.939  -1.181   **

---
Signif. codes:  0 '***' 0.001 '**' 0.01 '*' 0.05 '.' 0.1 ' ' 1
```

**OSHA_S  Onset of symptom to HOSPITAL ADMISSION (survivors, Period A)**

```
Number of studies combined: k = 2

                        mean           95%-CI
Fixed effect model      10.0082 [9.5503; 10.4880]
Random effects model  9.8090 [8.3176; 11.5677]

Quantifying heterogeneity:
 tau^2 = 0.0129; tau = 0.1138;
 I^2 = 91.4% [69.5%; 97.5%]; H = 3.40 [1.81; 6.38]
Test of heterogeneity:
     Q    d.f.  p-value
 11.56    1    0.0007
```

**OSHA_S  Onset of symptom to HOSPITAL ADMISSION (survivors, Period B)**

```
Number of studies combined: k = 4

                       mean          95%-CI
Fixed effect model      6.4189 [6.1113; 6.7419]
Random effects model 6.1356 [4.3805; 8.5938]

Quantifying heterogeneity:
 tau^2 = 0.1118 [0.0311; 1.7176]; tau = 0.3343 [0.1763; 1.3106];
 I^2 = 97.1% [94.9%; 98.4%]; H = 5.88 [4.42; 7.82]
Test of heterogeneity:
      Q d.f.  p-value
 103.68    3 < 0.0001
```

No identified outliers
-------------------------
No identified influencers
-------------------------

---------------------------

#### Onset of symptoms to hospital admission (non survivor)

---------------------------

```
Number of studies combined: k = 7

                       mean          95%-CI
Fixed effect model   10.1556 [9.7764; 10.5496]
Random effects model 10.1471 [9.6133; 10.7106]

Quantifying heterogeneity:
 tau^2 = 0.0021 [0.0000; 0.4618]; tau = 0.0461 [0.0000; 0.6796];
 I^2 = 56.5% [0.0%; 81.3%]; H = 1.52 [1.00; 2.31]
```

```
Test of heterogeneity:
    Q d.f. p-value
 13.79    6  0.0321
```

**Meta-regression with period as a categorical moderator**

```
Multivariate Meta-Analysis Model (k = 7; method: REML)

Variance Components:

         estim   sqrt  nlvls  fixed  factor
sigma^2    1.796  1.340      2     no  period

Test for Residual Heterogeneity: QE(df = 5) = 19.746, p-val = 0.001
Test of Moderators (coefficient 2): QM(df = 1) = 0.046, p-val = 0.831

Model Results:
                         estimate    se    zval   pval   ci.lb   ci.ub
            intrcpt     10.344  1.389   7.445  <.001   7.621  13.067  ***
            periodB     -0.416  1.944  -0.214  0.831  -4.227   3.395
---
Signif. codes:  0 '***' 0.001 '**' 0.01 '*' 0.05 '.' 0.1 ' ' 1
```

Identified outliers
-------------------------
"7"

Results with outliers removed
-----------------------------
Number of studies combined: k = 6

```
                           mean           95%-CI
Fixed effect model   10.1733 [9.7931; 10.5684]
Random effects model 10.1822 [9.6489; 10.7450]

Quantifying heterogeneity:
             tau^2 = 0.0021 [0.0000; 0.0253]; tau = 0.0455 [0.0000; 0.1589];
             I^2 = 47.3% [0.0%; 79.1%]; H = 1.38 [1.00; 2.19]

Test of heterogeneity:
                Q d.f. p-value
             9.48    5  0.0912
```

No influencers detected
-----------------

---------------------------

#### Onset of symptoms to ICU admission

---------------------------

Number of studies combined: k = 6

```
                   mean           95%-CI
Fixed effect model   9.7414 [9.2019; 10.3125]
```

**Random effects model 9.7414 [9.2019; 10.3125]**

```
Quantifying heterogeneity:
 tau^2 = 0 [0.0000; 0.0239]; tau = 0 [0.0000; 0.1546];
 I^2 = 0.0% [0.0%; 43.0%]; H = 1.00 [1.00; 1.32]

Test of heterogeneity:
   Q d.f. p-value
 2.22    5  0.8173
```

**Meta-regression with Period as a Categorical Moderator:**

```
Multivariate Meta-Analysis Model (k = 6; method: REML)

Variance Components:

         estim   sqrt  nlvls  fixed  factor
sigma^2  0.102  0.319      2     no  period

Test for Residual Heterogeneity:
QE(df = 4) = 1.618, p-val = 0.806

Test of Moderators (coefficient 2):
QM(df = 1) = 0.275, p-val = 0.600

Model Results:

         estimate     se     zval    pval   ci.lb   ci.ub
intrcpt     9.818  0.456   21.540  <.001   8.925  10.712  ***
periodB    -0.418  0.797   -0.525  0.600  -1.981   1.144

---
Signif. codes:  0 '***' 0.001 '**' 0.01 '*' 0.05 '.' 0.1 ' ' 1
```

➔ There are no significant differences between the two subsets.

**Outliers:**

Not detected

**Influencers:**

Not detected

--------------------------------

#### Hospital stay

--------------------------------

Number of studies combined: k = 13

```
                        mean            95%-CI
Fixed effect model   13.1637 [13.0137; 13.3153]
Random effects model 14.5163 [12.5749; 16.7574]

Quantifying heterogeneity:
 tau^2 = 0.0669 [0.0330; 0.1895]; tau = 0.2586 [0.1818; 0.4353];
 I^2 = 98.8% [98.6%; 99.1%]; H = 9.29 [8.35; 10.35]

Test of heterogeneity:
       Q d.f.  p-value
 1036.63   12 < 0.0001
```

**Meta-regression with period as a categorical moderator**

```
Multivariate Meta-Analysis Model (k = 13; method: REML)

Variance Components:

         estim   sqrt  nlvls  fixed  factor
sigma^2  3.705  1.925      2     no  period

Test for Residual Heterogeneity: QE(df = 11) = 385.899, p-val < .001
```

Test of Moderators (coefficient 2): QM(df = 1) = 4.867, p-val = 0.027

Model Results:

```
         estimate     se    zval   pval   ci.lb    ci.ub
intrcpt    12.350  1.926  6.411  <.001  8.574  16.126  ***
periodB     6.053  2.744  2.206  0.027  0.675  11.431    *
```

---

Signif. codes: 0 ‘***’ 0.001 ‘**’ 0.01 ‘*’ 0.05 ‘.’ 0.1 ‘ ’ 1

➔ Period is a significant moderator, we study the two subsets.

---------------------------

**Hospital stay ( Period A)**

---------------------------

Number of studies combined: k = 7

```
                        mean        95%-CI
Fixed effect model      12.4650   [12.3145; 12.6174]
Random effects model    12.7035.  [11.5800; 13.9361]

Quantifying heterogeneity:
      tau^2 = 0.0145 [0.0054; 0.0757]; tau = 0.1204 [0.0734; 0.2752];
      I^2 = 96.5% [94.6%; 97.7%]; H = 5.34 [4.30; 6.63]

Test of heterogeneity:
         Q d.f.  p-value
      170.87    6 < 0.0001
```

Identified outliers

-------------------------

"1", "2", "3", "6", "7"

Results with outliers removed

-----------------------------

Number of studies combined: k = 2

```
                        mean        95%-CI
Fixed effect model      13.1260   [12.5722; 13.7041]
Random effects model    13.1260   [12.5722; 13.7041]

Quantifying heterogeneity:
 tau^2 = 0; tau = 0; I^2 = 0.0%; H = 1.00

Test of heterogeneity:
   Q d.f. p-value
 0.87    1  0.3516
```

No influencers detected

-----------------

---------------------------

**Hospital stay (Period B)**

---------------------------

Number of studies combined: k = 6

```
                        mean        95%-CI
Fixed effect model      20.3864     [19.6969; 21.1000]
Random effects model    17.1384     [13.4245; 21.8797]

Quantifying heterogeneity:
      tau^2 = 0.0881 [0.0314; 0.5546];
      tau = 0.2968 [0.1771; 0.7447];
      I^2 = 97.0% [95.3%; 98.1%];
      H = 5.79 [4.62; 7.25]
```

Test of heterogeneity:
Q d.f. p-value
167.54 5 < 0.0001

Identified outliers
-------------------------
"9", "12", "13"

Results with outliers removed
-----------------------------
Number of studies combined: k = 3

| | mean | 95%-CI |
|---|---|---|
| Fixed effect model | 19.6991 | [18.3149; 21.1879] |
| Random effects model | 19.6991 | [18.3149; 21.1879] |

Quantifying heterogeneity:
tau^2 = 0 [<0.0000; <0.0000]; tau = 0 [<0.0000; <0.0000];
I^2 = 0.0% [0.0%; 0.0%]; H = 1.00 [1.00; 1.00]

Test of heterogeneity:
Q d.f. p-value
0.02 2 0.9903

No influencers detected
-----------------

---------------------------

#### Hospital stay (survivors)

---------------------------

Number of studies combined: k = 15

| | mean | 95%-CI |
|---|---|---|
| Fixed effect model | 24.9945 | [24.7754; 25.2155] |
| Random effects model | 14.6950 | [12.8474; 16.8082] |

Quantifying heterogeneity:
tau^2 = 0.0676 [0.0348; 0.1681]; tau = 0.2599 [0.1866; 0.4101];
I^2 = 99.5% [99.5%; 99.6%]; H = 14.61 [13.58; 15.71]

Test of heterogeneity:
Q d.f. p-value
2988.40 14 0

**Meta-regression with period as a categorical moderator**

Multivariate Meta-Analysis Model (k = 15; method: REML)

Variance Components:

| | estim | sqrt | nlvls | fixed | factor |
|---|---|---|---|---|---|
| sigma^2 | 7.291 | 2.700 | 2 | no | period |

Test for Residual Heterogeneity:
QE(df = 13) = 2688.857, p-val < .001
Test of Moderators (coefficient 2):
QM(df = 1) = 10.793, p-val = 0.001

Model Results:

| | estimate | se | zval | pval | ci.lb | ci.ub | |
|---|---|---|---|---|---|---|---|
| intrcpt | 11.242. | 2.713 | 4.145 | <.001 | 5.926 | 16.559 | *** |

```
periodB    12.580    3.829    3.285    0.001  5.075  20.084   **
---
Signif. codes:  0 ‘***’ 0.001 ‘**’ 0.01 ‘*’ 0.05 ‘.’ 0.1 ‘ ’ 1
```

➔ Period is a significant moderator, we study the two subsets.

---------------------------

**Hospital stay (survivors, Period A)**

---------------------------

Number of studies combined: k = 4

```
                         mean          95%-CI
Fixed effect model       11.3077   [10.8088; 11.8295]
Random effects model 11.0589   [10.2642; 11.9152]

Quantifying heterogeneity:
         tau^2 = 0.0032 [0.0000; 0.0572]; tau = 0.0563 [0.0000; 0.2391];
         I^2 = 58.6% [0.0%; 86.2%]; H = 1.55 [1.00; 2.70]
Test of heterogeneity:
            Q d.f. p-value
         7.25    3  0.0643
```

No outliers detected

-----------------

Identified influencers

-------------------------

" Zhou Fei et al. 2020"

Results with influencers removed

-----------------------------

Number of studies combined: k = 3

```
                         mean          95%-CI
Fixed effect model       10.6128 [9.9462; 11.3241]
Random effects model 10.6128 [9.9462; 11.3241]

Quantifying heterogeneity:
         tau^2 = 0 [0.0000; 0.0082]; tau = 0 [0.0000; 0.0904];
         I^2 = 0.0% [0.0%; 0.0%]; H = 1.00 [1.00; 1.00]
Test of heterogeneity:
            Q d.f. p-value
         0.14    2  0.9319
```

---------------------------

**Hospital stay (survivors, Period B)**

---------------------------

Number of studies combined: k = 11

```
                          mean           95%-CI
Fixed effect model        25.7914   [25.5609; 26.0239]
Random effects model 16.3765    [14.3411; 18.7008]


Quantifying heterogeneity:
         tau^2 = 0.0476 [0.0219; 0.1420];
         tau = 0.2181 [0.1479; 0.3769];
         I^2 = 99.4% [99.3%; 99.5%];
         H = 13.22 [12.05; 14.50]
Test of heterogeneity:
              Q        d.f. p-value
         1746.93   10      0
```

Identified outliers (REML)
-------------------------
"12",”4”

Results with outliers removed
-----------------------------
Number of studies combined: k = 9

| | mean | 95%-CI |
|---|---|---|
| Fixed effect model | 14.7639. | [14.3430; 15.1972] |
| Random effects model | 14.8800 | [13.9608; 15.8598] |

Quantifying heterogeneity:
tau^2 = 0.0066 [0.0015; 0.0361];
tau = 0.0810 [0.0389; 0.1899];
I^2 = 71.9% [44.8%; 85.7%];
H = 1.89 [1.35; 2.65]

Test of heterogeneity:

| Q | d.f. | p-value |
|---|---|---|
| 28.52 | 8 | 0.0004 |

Identified influencers
-------------------------
" Wang Lang et al. 2020 "

Results with influencers removed
-----------------------------
Number of studies combined: k = 10

| | mean | 95%-CI |
|---|---|---|
| Fixed effect model | 15.8195 | [15.4224; 16.2268] |
| Random effects model | 15.4613 | [14.1770; 16.8620] |

Quantifying heterogeneity:
tau^2 = 0.0166 [0.0065; 0.0570];
tau = 0.1289 [0.0804; 0.2387];
I^2 = 92.8% [88.8%; 95.4%];
H = 3.72 [2.99; 4.64]

Test of heterogeneity:

| Q | d.f. | p-value |
|---|---|---|
| 124.76 | 9 | < 0.0001 |

---------------------------

#### Hospital stay (non survivors)

---------------------------

Number of studies combined: k = 12

| | mean | 95%-CI |
|---|---|---|
| Fixed effect model | 8.5894 | [8.2334; 8.9607] |
| Random effects model | 8.6307 | [7.2257; 10.3088] |

Quantifying heterogeneity:
tau^2 = 0.0846 [0.0359; 0.2507]; tau = 0.2908 [0.1895; 0.5007];
I^2 = 93.3% [90.1%; 95.5%]; H = 3.86 [3.18; 4.70]

Test of heterogeneity:

| Q | d.f. | p-value |
|---|---|---|
| 164.21 | 11 | < 0.0001 |

**Meta-regression with period as a categorical moderator**

```
Multivariate Meta-Analysis Model (k = 12; method: REML)

Variance Components:
           estim   sqrt  nlvls  fixed  factor
sigma^2    2.313  1.521      2     no  period

Test for Residual Heterogeneity: QE(df = 10) = 140.877, p-val < .001
Test of Moderators (coefficient 2): QM(df = 1) = 0.128, p-val = 0.720

Model Results:
          estimate     se    zval    pval    ci.lb   ci.ub
intrcpt      8.197  1.546   5.301  <.001   5.166  11.227  ***
periodB     -0.781  2.180  -0.358  0.720  -5.053   3.491

---
Signif. codes:  0 '***' 0.001 '**' 0.01 '*' 0.05 '.' 0.1 ' ' 1
```

Identified outliers
-------------------------
"2", "4", "5", "6", "8", "9", "10"

Results with outliers removed
-----------------------------
Number of studies combined: k = 5

```
                        mean      95%-CI
Fixed effect model    8.1154  [7.6075; 8.6572]
Random effects model  8.1154  [7.6075; 8.6572]

Quantifying heterogeneity:
    tau^2 = 0 [0.0000; 0.1947]; tau = 0 [0.0000; 0.4413];
    I^2 = 0.0% [0.0%; 74.2%]; H = 1.00 [1.00; 1.97]

Test of heterogeneity:
       Q d.f. p-value
    3.22    4  0.5217
```

Identified influencers
-------------------------
"2", "4", "5", "7", "8", "9"

Results with influencers removed
-----------------------------
Number of studies combined: k = 6

```
                       mean       95%-CI
Fixed effect model   8.9698 [8.2314;  9.7744]
Random effects model 9.3691 [8.0886; 10.8522]

Quantifying heterogeneity:
    tau^2 = 0.0166 [0.0000; 0.1459]; tau = 0.1287 [0.0000; 0.3819];
    I^2 = 51.0% [0.0%; 80.5%]; H = 1.43 [1.00; 2.27]

Test of heterogeneity:
        Q d.f. p-value
    10.21    5  0.0695
```

---------------------------

#### ICU stay (survivors)

---------------------------

Number of studies combined: k = 4

mean 95%-CI
Fixed effect model 8.2197 [7.4244; 9.1001]
**Random effects model 9.0844 [6.4692; 12.7568]**

Quantifying heterogeneity:
tau^2 = 0.1017 [0.0236; 1.4870]; tau = 0.3189 [0.1535; 1.2194];
I^2 = 92.2% [83.2%; 96.4%]; H = 3.57 [2.44; 5.24]

Test of heterogeneity:
Q d.f. p-value
38.33 3 < 0.0001

---------------------------

#### ICU stay (non survivors)

---------------------------

Review: ICU stay (non survivors)
Number of studies combined: k = 5

mean 95%-CI
Fixed effect model 9.1004 [8.3399; 9.9302]
**Random effects model 8.9886 [7.4482; 10.8475]**

Quantifying heterogeneity:
tau^2 = 0.0302 [0.0016; 0.3152]; tau = 0.1738 [0.0398; 0.5614];
I^2 = 68.4% [18.5%; 87.7%]; H = 1.78 [1.11; 2.85]

Test of heterogeneity:
Q d.f. p-value
12.64 4 0.0132

**Meta-regression with Period as a Categorical Moderator:**

Multivariate Meta-Analysis Model (k = 5; method: REML)

Variance Components:

| | estim | sqrt | nlvls | fixed | factor |
|---|---|---|---|---|---|
| sigma^2 | 0.163 | 0.403 | 2 | no | period |

Test for Residual Heterogeneity:
QE(df = 3) = 5.474, p-val = 0.140

Test of Moderators (coefficient 2):
**QM(df = 1) = 5.654, p-val = 0.017**

Model Results:

| | estimate | se | zval | pval | ci.lb | ci.ub | |
|---|---|---|---|---|---|---|---|
| intrcpt | 8.423 | 0.580 | 14.515 | <.001 | 7.285 | 9.560 | *** |
| periodB | 3.377 | 1.420 | 2.378 | 0.017 | 0.593 | 6.161 | * |

---
Signif. codes: 0 ‘***’ 0.001 ‘**’ 0.01 ‘*’ 0.05 ‘.’ 0.1 ‘ ’ 1

➔ Period is a significant moderator, we study the two subsets.

---------------------------

##### ICU stay (non survivors, Period A)

---------------------------

Number of studies combined: k = 4

```
                      mean         95%-CI
Fixed effect model      8.5900 [7.8001; 9.4600]
```
**Random effects model 8.3558 [7.1891; 9.7117]**

```
Quantifying heterogeneity:
    tau^2 = 0.0100 [0.0000; 0.3007]; tau = 0.0999 [0.0000; 0.5484];
    I^2 = 40.9% [0.0%; 80.0%]; H = 1.30 [1.00; 2.24]

Test of heterogeneity:
       Q  d.f.   p-value
    5.08   3     0.1660
```

---------------------------

**ICU stay (non survivors, Period B)**

----------------------------

```
Review:     ICU stay (non survivors, Period B)
    mean         95%-CI
 11.8000     [9.6160; 14.4801]
```

---------------------------

#### Onset of symptoms to death

---------------------------

Number of studies combined: k = 10

```
                         mean            95%-CI
Fixed effect model       17.9562   [17.3385; 18.5959]
Random effects model  17.7899   [16.0556; 19.7114]

Quantifying heterogeneity:
    tau^2 = 0.0198 [0.0060; 0.0716]; tau = 0.1407 [0.0773; 0.2675];
    I^2 = 86.3% [76.7%; 91.9%]; H = 2.70 [2.07; 3.52]

Test of heterogeneity:
        Q d.f.  p-value
    65.72    9 < 0.0001
```

**Meta-regression with period as a categorical moderator**

```
Multivariate Meta-Analysis Model (k = 10; method: REML)

Variance Components:
            estim   sqrt  nlvls  fixed  factor
    sigma^2   2.466  1.570     2     no  period

Test for Residual Heterogeneity: QE(df = 8) = 58.532, p-val < .001
Test of Moderators (coefficient 2): QM(df = 1) = 0.088, p-val = 0.767

Model Results:
            estimate    se    zval   pval   ci.lb   ci.ub
    intrcpt   17.135  1.626  10.541  <.001  13.949  20.321  ***
    periodB    0.683  2.309   0.296  0.767  -3.843   5.209

---
Signif. codes:  0 ‘***’ 0.001 ‘**’ 0.01 ‘*’ 0.05 ‘.’ 0.1 ‘ ’ 1
```

Identified outliers

-------------------------

"3", "4"

Results with outliers removed
-----------------------------
Number of studies combined: k = 8

```
                          mean          95%-CI
Fixed effect model       16.6963    [16.0343; 17.3857]
Random effects model 16.6627   [15.4759; 17.9406]

Quantifying heterogeneity:
         tau^2 = 0.0054 [0.0001; 0.0323]; tau = 0.0736 [0.0078; 0.1798];
         I^2 = 57.1% [5.7%; 80.5%]; H = 1.53 [1.03; 2.26]
Test of heterogeneity:
           Q d.f. p-value
         16.32    7  0.0224
```

Identified influencers
-------------------------
"4"
Results with influencers removed
-----------------------------
Number of studies combined: k = 9

```
                          mean          95%-CI
Fixed effect model       16.9847   [16.3310; 17.6645]
Random effects model 17.2107   [15.6942; 18.8738]

Quantifying heterogeneity:
         tau^2 = 0.0125 [0.0022; 0.0598]; tau = 0.1117 [0.0465; 0.2446];
         I^2 = 71.4% [43.5%; 85.5%]; H = 1.87 [1.33; 2.63]
Test of heterogeneity:
           Q d.f. p-value
         27.94    8  0.0005
```

---------------------------
#### Onset of symptoms to discharge
---------------------------
Number of studies combined: k = 3.  --ONLY 3 STUDIES--

```
                          mean          95%-CI
Fixed effect model       23.2570  [22.7084; 23.8189]
Random effects model 21.7641  [18.1316; 26.1243]

Quantifying heterogeneity:
         tau^2 = 0.0254 [0.0064; 1.0364]; tau = 0.1593 [0.0797; 1.0180];
         I^2 = 97.7% [95.5%; 98.8%]; H = 6.54 [4.73; 9.05]

Test of heterogeneity:
           Q d.f.  p-value
85.54    2 < 0.0001
```

#### GENERAL POPULATION: Meta-analysis of single proportions

------------------------------------------------------------------------------------------------------
#### PT - % presymptomatic transmission
------------------------------------------------------------------------------------------------------

Number of studies combined: k = 5

```
                    proportion        95%-CI
```

| | | |
|---|---|---|
| Fixed effect model | 0.2741 | [0.2400; 0.3111] |
| **Random effects model** | **0.2933** | **[0.1129; 0.5750]** |

Quantifying heterogeneity:

tau^2 = 1.7601 [0.5926; 15.1195]; tau = 1.3267 [0.7698; 3.8884];
I^2 = 97.4% [95.8%; 98.4%]; H = 6.22 [4.90; 7.91]

Test of heterogeneity:

| Q | d.f. | p-value |
|---|---|---|
| 154.83 | 4 | < 0.0001 |

**Meta-regression with Period as a Categorical Moderator:**

Mixed-Effects Model (k = 5; tau^2 estimator: REML)

tau^2 (estimated amount of residual heterogeneity): 2.3487 (SE = 1.9659)
tau (square root of estimated tau^2 value): 1.5325
I^2 (residual heterogeneity / unaccounted variability): 97.75%
H^2 (unaccounted variability / sampling variability): 44.35
R^2 (amount of heterogeneity accounted for): 0.00%

Test for Residual Heterogeneity:

QE(df = 3) = 107.7237, p-val < .0001

Test of Moderators (coefficient 2):

**QM(df = 1) = 0.0254, p-val = 0.8734**

Model Results:

| | estimate | se | zval | pval | ci.lb | ci.ub |
|---|---|---|---|---|---|---|
| intrcpt | -1.0158 | 1.0943 | -0.9282 | 0.3533 | -3.1607 | 1.1291 |
| periodB | 0.2254 | 1.4150 | 0.1593 | 0.8734 | -2.5479 | 2.9988 |

---
Signif. codes: 0 ‘***’ 0.001 ‘**’ 0.01 ‘*’ 0.05 ‘.’ 0.1 ‘ ’ 1

➔ There are **no significant differences** between the two subsets.

**Outliers :**

Not detected.

**Influencers :**

"Wei Wycliffe et al. 2020"

**Results with influencers removed:**

Number of studies combined: k = 4

| | proportion | 95%-CI |
|---|---|---|
| Fixed effect model | 0.3039 | [0.2660; 0.3446] |
| **Random effects model** | **0.3904** | **[0.1838; 0.6456]** |

Quantifying heterogeneity:

tau^2 = 1.0922 [0.3264; 15.2694]; tau = 1.0451 [0.5713; 3.9076];
I^2 = 97.6% [95.9%; 98.6%]; H = 6.45 [4.93; 8.45]

Test of heterogeneity:

| Q | d.f. | p-value |
|---|---|---|
| 125.00 | 3 | < 0.0001 |

---

#### AP - Asymptomatic patients

-----------------------------------------------------------------------------------------------------------

Number of studies combined: k = 14

| | proportion | 95%-CI |
|---|---|---|
| Fixed effect model | 0.0587 | [0.0482; 0.0713] |
| Random effects model | 0.0854 | [0.0404; 0.1718] |

Quantifying heterogeneity:
tau^2 = 1.8933 [0.7934; 4.9080];
tau = 1.3760 [0.8907; 2.2154];
I^2 = 89.8% [84.7%; 93.2%];
H = 3.14 [2.56; 3.84]

Test of heterogeneity:

| Q | d.f. | p-value |
|---|---|---|
| 127.83 | 13 | < 0.0001 |

**Meta-regression with period as moderator**

Mixed-Effects Model (k = 14; tau^2 estimator: REML)

tau^2 (estimated amount of residual heterogeneity): 2.0653 (SE = 1.0156)
tau (square root of estimated tau^2 value): 1.4371
I^2 (residual heterogeneity / unaccounted variability): 91.85%
H^2 (unaccounted variability / sampling variability): 12.27
R^2 (amount of heterogeneity accounted for): 0.00%

Test for Residual Heterogeneity:
QE(df = 12) = 122.8295, p-val < .0001

Test of Moderators (coefficient 2):
QM(df = 1) = 0.0219, p-val = 0.8822

Model Results:

| | estimate | se | zval | pval | ci.lb | ci.ub | |
|---|---|---|---|---|---|---|---|
| intrcpt | -2.4026 | 0.4815 | -4.9900 | <.0001 | -3.3463 | -1.4589 | *** |
| periodB | 0.1486 | 1.0029 | 0.1481 | 0.8822 | -1.8170 | 2.1142 | |

---
Signif. codes: 0 ‘***’ 0.001 ‘**’ 0.01 ‘*’ 0.05 ‘.’ 0.1 ‘ ’ 1

Identified outliers
----------------------------------------

"Hu et al. 2020", "Tian Suochen et al. 2020", "Wang X. et al. 2020"

Results with outliers removed
-----------------------------

Number of studies combined: k = 11

| | proportion | 95%-CI |
|---|---|---|
| Fixed effect model | 0.0546 | [0.0434; 0.0685] |
| Random effects model | 0.0544 | [0.0417; 0.0706] |

Quantifying heterogeneity:
tau^2 = 0.0251 [0.0000; 1.1343];
tau = 0.1583 [0.0000; 1.0651];
I^2 = 17.9% [0.0%; 58.0%];
H = 1.10 [1.00; 1.54]

Test of heterogeneity:

```
            Q d.f. p-value
         12.19   10  0.2727

Identified influencers
----------------------------------------
            "Hu et al. 2020"

Results with influencer removed
-----------------------------

Number of studies combined: k = 13

                              proportion         95%-CI
            Fixed effect model          0.0489         [0.0399; 0.0598]
            Random effects model     0.0609         [0.0352; 0.1033]

            Quantifying heterogeneity:
                  tau^2 = 0.7394 [0.2177; 2.2324];
                  tau = 0.8599 [0.4666; 1.4941];
                  I^2 = 79.2% [65.2%; 87.6%];
                  H = 2.20 [1.69; 2.84]

            Test of heterogeneity:
                       Q d.f.  p-value
 57.83   12 < 0.0001
```

-----------------------------------------------------------------------------------------------------------

#### ICUA - ICU admissions vs all hospitalized patients with COVID 19

-----------------------------------------------------------------------------------------------------------

```
Number of studies combined: k = 19

                              proportion         95%-CI
              Fixed effect model          0.1227         [0.1125; 0.1336]
              Random effects model     0.1309         [0.0914; 0.1841]

              Quantifying heterogeneity:
                      tau^2 = 0.6455 [0.3138; 1.6617];
                      tau = 0.8034 [0.5602; 1.2891];
                      I^2 = 93.9% [91.7%; 95.5%];
                      H = 4.04 [3.48; 4.69]

              Test of heterogeneity:
                         Q d.f.  p-value
                      293.88   18 < 0.0001
```

**Meta-regression with period as moderator**

```
          Mixed-Effects Model (k = 19; tau^2 estimator: REML)

          tau^2 (estimated amount of residual heterogeneity):     0.6649 (SE = 0.2742)
          tau (square root of estimated tau^2 value):           0.8154
          I^2 (residual heterogeneity / unaccounted variability): 93.09%
          H^2 (unaccounted variability / sampling variability):   14.47
          R^2 (amount of heterogeneity accounted for):           0.00%

          Test for Residual Heterogeneity:   QE(df = 17) = 293.8748, p-val < .0001

          Test of Moderators (coefficient 2):  QM(df = 1) = 0.5589, p-val = 0.4547

          Model Results:
                          estimate     se     zval    pval    ci.lb    ci.ub
                 intrcpt   -1.8261  0.2279  -8.0130  <.0001  -2.2728  -1.3794  ***
                 periodB   -0.4234  0.5663  -0.7476  0.4547  -1.5333   0.6866
```

---
Signif. codes:  0 ‘***’ 0.001 ‘**’ 0.01 ‘*’ 0.05 ‘.’ 0.1 ‘ ’ 1

Identified outliers
------------------------------------------
"Guan et al. 2020", "Huang Chaolin et al. 2020 ", "Lian et al. 2020", "Liang et al. 2020", "Wang Dawei et al. 2020", "Wu Chaomin et al. 2020 ", "Zhou Fei et al. 2020"

Results with outliers removed
-----------------------------

Number of studies combined: k = 12

| | proportion | 95%-CI |
|---|---|---|
| Fixed effect model | 0.1711 | [0.1474; 0.1977] |
| Random effects model | 0.1675 | [0.1417; 0.1969] |

Quantifying heterogeneity:
tau^2 = 0.0132 [0.0000; 1.4794];
tau = 0.1148 [0.0000; 1.2163];
I^2 = 40.2% [0.0%; 69.7%];
H = 1.29 [1.00; 1.82]

Test of heterogeneity:

| Q | d.f. | p-value |
|---|---|---|
| 18.39 | 11 | 0.0730 |

Identified influencer
----------------------------------------
" Lian et al. 2020 "

Results with influencer removed
-----------------------------
Number of studies combined: k = 18

| | proportion | 95%-CI |
|---|---|---|
| Fixed effect model | 0.1326 | [0.1215; 0.1446] |
| Random effects model | 0.1470 | [0.1075; 0.1977] |

Quantifying heterogeneity:
tau^2 = 0.4493 [0.2105; 1.4152];
tau = 0.6703 [0.4588; 1.1896];
I^2 = 92.8% [90.1%; 94.8%];
H = 3.74 [3.18; 4.39]

Test of heterogeneity:

| Q | d.f. | p-value |
|---|---|---|
| 237.28 | 17 | < 0.0001 |

---------------------------------------------------------------------------------------------------------

#### DH - Death vs all hospitalized patients with COVID 19

---------------------------------------------------------------------------------------------------------

Number of studies combined: k = 20

| | proportion | 95%-CI |
|---|---|---|
| Fixed effect model | 0.1192 | [0.1090; 0.1302] |
| Random effects model | 0.0762 | [0.0459; 0.1241] |

Quantifying heterogeneity:
tau^2 = 1.2935 [0.6930; 3.5033];
tau = 1.1373 [0.8325; 1.8717];

I^2 = 94.5% [92.6%; 95.8%];
H = 4.25 [3.69; 4.89]

Test of heterogeneity:
Q d.f. p-value
342.49 19 < 0.0001

**Meta-regression with period as moderator**
Mixed-Effects Model (k = 20; tau^2 estimator: REML)

tau^2 (estimated amount of residual heterogeneity): 1.3354 (SE = 0.5127)
tau (square root of estimated tau^2 value): 1.1556
I^2 (residual heterogeneity / unaccounted variability): 95.23%
H^2 (unaccounted variability / sampling variability): 20.96
R^2 (amount of heterogeneity accounted for): 0.00%

Test for Residual Heterogeneity: QE(df = 18) = 341.2798, p-val < .0001
Test of Moderators (coefficient 2): QM(df = 1) = 0.6018, p-val = 0.4379

Model Results:

| | estimate | se | zval | pval | ci.lb | ci.ub | |
|---|---|---|---|---|---|---|---|
| intrcpt | -2.3886 | 0.3136 | -7.6170 | <.0001 | -3.0033 | -1.7740 | *** |
| periodB | -0.5384 | 0.6941 | -0.7758 | 0.4379 | -1.8988 | 0.8219 | |

Identified outliers
----------------------------------------
"Cao Bin et al. 2020", "Guan et al. 2020", "Liang et al. 2020", "Tang et al. 2020", "Tian Sijia et al. 2020", "Wang Dawei et al. 2020", "Wu Chaomin et al. 2020 ", "Zhang Guqin et al. 2020", "Zhou Fei et al. 2020", "Tindale et al. 2020", "Qian et al. 2020"

Results with outliers removed
-----------------------------
Number of studies combined: k = 9

| | proportion | 95%-CI |
|---|---|---|
| Fixed effect model | 0.1323 | [0.1159; 0.1506] |
| Random effects model | 0.1323 | [0.1159; 0.1506] |

Quantifying heterogeneity:
tau^2 < 0.0001 [0.0000; 3.3572]; tau = 0.0014 [0.0000; 1.8323];
I^2 = 41.4% [0.0%; 73.0%]; H = 1.31 [1.00; 1.92]

Test of heterogeneity:
Q d.f. p-value
13.64 8 0.0916

No influencers detected

-----------------

------------------------------------------------------------------------------------------------------------

#### ICUD - Deaths from ICU

------------------------------------------------------------------------------------------------------------

Number of studies combined: k = 5

| | Proportion | 95%-CI |
|---|---|---|
| Fixed effect model | 0.3968 | [0.3516; 0.4439] |
| **Random effects model** | **0.3503** | **[0.1942; 0.5467]** |

Quantifying heterogeneity:
tau^2 = 0.6104 [0.0956; 7.4028]; tau = 0.7813 [0.3092; 2.7208];
I^2 = 79.0% [49.9%; 91.2%]; H = 2.18 [1.41; 3.36]

Test of heterogeneity:

| Q | d.f. | p-value |
|---|---|---|
| 19.01 | 4 | 0.0008 |

**Meta-regression with Period as a Categorical Moderator:**

Mixed-Effects Model (k = 5; tau^2 estimator: REML)

tau^2 (estimated amount of residual heterogeneity): 0.3267 (SE = 0.4304)
tau (square root of estimated tau^2 value): 0.5715
I^2 (residual heterogeneity / unaccounted variability): 74.19%
H^2 (unaccounted variability / sampling variability): 3.88
R^2 (amount of heterogeneity accounted for): 46.49%

Test for Residual Heterogeneity:
QE(df = 3) = 12.2213, p-val = 0.0067

Test of Moderators (coefficient 2):
**QM(df = 1) = 3.3701, p-val = 0.0664**

Model Results:

| | estimate | se | zval | pval | ci.lb | ci.ub | |
|---|---|---|---|---|---|---|---|
| intrcpt | -1.2401 | 0.4930 | -2.5151 | 0.0119 | -2.2064 | -0.2737 | * |
| periodB | 1.2013 | 0.6544 | 1.8358 | 0.0664 | -0.0813 | 2.4838 | . |

---
Signif. codes: 0 ‘***’ 0.001 ‘**’ 0.01 ‘*’ 0.05 ‘.’ 0.1 ‘ ’ 1

➔ There are **no significant differences** between the two subsets.

**Outliers :**
"Wang Dawei et al. 2020", "Yang et al. 2020"

**Results with outliers removed:**

Number of studies combined: k = 3

| | proportion | 95%-CI |
|---|---|---|
| Fixed effect model | 0.3830 | [0.3343; 0.4341] |
| **Random effects model** | **0.3830** | **[0.3343; 0.4341]** |

Quantifying heterogeneity:
tau^2 < 0.0001 [0.0000; 28.0285]; tau = 0.0015 [0.0000; 5.2942];
I^2 = 0.0% [0.0%; 89.1%]; H = 1.00 [1.00; 3.03]

Test of heterogeneity:

| Q | d.f. | p-value |
|---|---|---|
| 1.91 | 2 | 0.3853 |

**Influencers :**
"Wang Dawei et al. 2020", "Yang et al. 2020"

**Results with influencers removed:**

Number of studies combined: k = 3

| | proportion | 95%-CI |
|---|---|---|
| Fixed effect model | 0.3830 | [0.3343; 0.4341] |
| **Random effects model** | **0.3830** | **[0.3343; 0.4341]** |

Quantifying heterogeneity:

tau^2 < 0.0001 [0.0000; 28.0285]; tau = 0.0015 [0.0000; 5.2942];
I^2 = 0.0% [0.0%; 89.1%]; H = 1.00 [1.00; 3.03]

Test of heterogeneity:

| Q | d.f. | p-value |
|---|---|---|
| 1.91 | 2 | 0.3853 |

---

#### DisH - Discharged from all hospitalized patients with COVID 19

---

Number of studies combined: k = 25

| | proportion | 95%-CI |
|---|---|---|
| Fixed effect model | 0.3606 | [0.3444; 0.3771] |
| Random effects model | 0.4934 | [0.3601; 0.6276] |

Quantifying heterogeneity:
tau^2 = 1.8064 [1.1515; 4.6959];
tau = 1.3440 [1.0731; 2.1670];
I^2 = 97.4% [96.9%; 97.9%];
H = 6.26 [5.67; 6.90]

Test of heterogeneity:

| Q | d.f. | p-value |
|---|---|---|
| 939.39 | 24 | < 0.0001 |

**Meta-regression with period as moderator**

Mixed-Effects Model (k = 25; tau^2 estimator: REML)

tau^2 (estimated amount of residual heterogeneity): 1.9066 (SE = 0.6032)
tau (square root of estimated tau^2 value): 1.3808
I^2 (residual heterogeneity / unaccounted variability): 98.00%
H^2 (unaccounted variability / sampling variability): 49.98
R^2 (amount of heterogeneity accounted for): 0.00%

Test for Residual Heterogeneity: QE(df = 23) = 936.8121, p-val < .0001
Test of Moderators (coefficient 2): QM(df = 1) = 0.0682, p-val = 0.7939

Model Results:

| | estimate | se | zval | pval | ci.lb | ci.ub |
|---|---|---|---|---|---|---|
| intrcpt | -0.0809 | 0.3660 | -0.2211 | 0.8250 | -0.7982 | 0.6364 |
| periodB | 0.1541 | 0.5898 | 0.2612 | 0.7939 | -1.0020 | 1.3101 |

---
Signif. codes: 0 ‘***’ 0.001 ‘**’ 0.01 ‘*’ 0.05 ‘.’ 0.1 ‘ ’ 1

Identified outliers

----------------------------------------

"Cao Jianlei et al. 2020", "Chen TL et al. 2020", "Chen T. et al. 2020", "Guan et al. 2020", "Huang Chaolin et al. 2020 ", "Lei et al. 2020", "Lian et al. 2020", "Tian Sijia et al. 2020", "Tindale et al. 2020", "Wu Jian et al. 2020", "Zhang Guqin et al. 2020", "Zhao Wen et al. 2020", "Zhou Fei et al. 2020", "Liu Lei et al. 2020",”Tian Suchen et al. 2020”

Results with outliers removed

-----------------------------

Number of studies combined: k = 10

| | proportion | 95%-CI |
|---|---|---|
| Fixed effect model | 0.3493 | [0.3161; 0.3841] |
| Random effects model | 0.3493 | [0.3161; 0.3841] |

Quantifying heterogeneity:
tau^2 = 0 [0.0000; 0.0267];
tau = 0 [0.0000; 0.1633];
I^2 = 0.0% [0.0%; 5.7%];
H = 1.00 [1.00; 1.03]

Test of heterogeneity:
Q d.f. p-value
3.59 9 0.9361

No influencers detected
-----------------

-----------------------------------------------------------------------------------------------------------

#### DisICU - Discharged from ICU

-----------------------------------------------------------------------------------------------------------

Number of studies combined: k = 6

| | proportion | 95%-CI |
|---|---|---|
| Fixed effect model | 0.4981 | [0.4518; 0.5444] |
| **Random effects model** | **0.4496** | **[0.2573; 0.6582]** |

Quantifying heterogeneity:
tau^2 = 0.9164 [0.2369; 9.0927]; tau = 0.9573 [0.4868; 3.0154];
I^2 = 85.4% [70.1%; 92.9%]; H = 2.62 [1.83; 3.74]

Test of heterogeneity:
Q d.f. p-value
34.21 5 < 0.0001

**Meta-regression with Period as a Categorical Moderator:**

Mixed-Effects Model (k = 6; tau^2 estimator: REML)

tau^2 (estimated amount of residual heterogeneity): 0.8973 (SE = 0.8094)
tau (square root of estimated tau^2 value): 0.9473
I^2 (residual heterogeneity / unaccounted variability): 85.75%
H^2 (unaccounted variability / sampling variability): 7.02
R^2 (amount of heterogeneity accounted for): 2.08%

Test for Residual Heterogeneity:
QE(df = 4) = 15.5983, p-val = 0.0036

Test of Moderators (coefficient 2):
**QM(df = 1) = 0.6998, p-val = 0.4029**

Model Results:

| | estimate | se | zval | pval | ci.lb | ci.ub |
|---|---|---|---|---|---|---|
| intrcpt | -0.4952 | 0.5562 | -0.8903 | 0.3733 | -1.5854 | 0.5950 |
| periodB | 0.7434 | 0.8887 | 0.8365 | 0.4029 | -0.9984 | 2.4852 |

---
Signif. codes: 0 '***' 0.001 '**' 0.01 '*' 0.05 '.' 0.1 ' ' 1
➔ There are **no significant differences** between the two subsets.

**Outliers :**
"Wang Dawei et al. 2020", "Yang et al. 2020"

**Results with outliers removed:**

Number of studies combined: k = 4

```
                 proportion           95%-CI
Fixed effect model          0.5461 [0.4967; 0.5946]
```

**Random effects model 0.5461 [0.4967; 0.5946]**

Quantifying heterogeneity:

tau^2 < 0.0001 [0.0000; 9.5866]; tau = 0.0008 [0.0000; 3.0962];
I^2 = 1.8% [0.0%; 85.0%]; H = 1.01 [1.00; 2.58]

Test of heterogeneity:

```
   Q d.f. p-value
3.06    3  0.3832
```

**Influencers :**

No decteted.

---

#### H - In hospital

---

Number of studies combined: k = 21

| | proportion | 95%-CI |
|---|---|---|
| Fixed effect model | 0.6443 | [0.6263; 0.6620] |
| Random effects model | 0.5979 | [0.4701; 0.7136] |

Quantifying heterogeneity:

tau^2 = 1.3358 [0.7731; 3.4337];
tau = 1.1558 [0.8793; 1.8530];
I^2 = 97.0% [96.3%; 97.6%];
H = 5.81 [5.19; 6.50]

Test of heterogeneity:

| Q | d.f. | p-value |
|---|---|---|
| 674.62 | 20. | < 0.0001 |

**Meta-regression with period as moderator**

Mixed-Effects Model (k = 21; tau^2 estimator: REML)

tau^2 (estimated amount of residual heterogeneity): 1.3585 (SE = 0.4784)
tau (square root of estimated tau^2 value): 1.1655
I^2 (residual heterogeneity / unaccounted variability): 97.14%
H^2 (unaccounted variability / sampling variability): 34.97
R^2 (amount of heterogeneity accounted for): 0.00%

Test for Residual Heterogeneity: QE(df = 19) = 567.9055, p-val < .0001
Test of Moderators (coefficient 2): QM(df = 1) = 0.9029, p-val = 0.3420

Model Results:

```
          estimate     se     zval    pval    ci.lb   ci.ub
intrcpt    0.5765  0.3259   1.7689  0.0769  -0.0623  1.2153  .
periodB   -0.5342  0.5622  -0.9502  0.3420  -1.6360  0.5676

---
Signif. codes:  0 ‘***’ 0.001 ‘**’ 0.01 ‘*’ 0.05 ‘.’ 0.1 ‘ ’ 1
```

Identified outliers

---

"Guan et al. 2020", "Huang Chaolin et al. 2020 ", "Lei et al. 2020", "Tang et al. 2020", "Tian Sijia et al. 2020", "Wen et al. 2020", "Xu Xiao-Wei et al. 2020", "Zhang Guqin et al. 2020", "Zhao Wen et al. 2020"

Results with outliers removed
-----------------------------
Number of studies combined: k = 12

| | proportion | 95%-CI |
|---|---|---|
| Fixed effect model | 0.6379 | [0.6142; 0.6611] |
| Random effects model | 0.6313 | [0.5984; 0.6630] |

Quantifying heterogeneity:
tau^2 = 0.0134 [0.0000; 0.1403];
tau = 0.1157 [0.0000; 0.3745];
I^2 = 21.2% [0.0%; 59.5%];
H = 1.13 [1.00; 1.57]

Test of heterogeneity:

| Q | d.f. | p-value |
|---|---|---|
| 13.96 | 11 | 0.2355 |

No influencers detected
-----------------

---

#### HICU - In hospital (ICU)

---

Number of studies combined: k = 7

| | proportion | 95%-CI |
|---|---|---|
| Fixed effect model | 0.1524 | [0.1209; 0.1902] |
| **Random effects model** | **0.1663** | **[0.0853; 0.2991]** |

Quantifying heterogeneity:
tau^2 = 0.7573 [0.1803; 3.9796]; tau = 0.8702 [0.4246; 1.9949];
I^2 = 85.8% [72.8%; 92.6%]; H = 2.66 [1.92; 3.68]

Test of heterogeneity:

| Q | d.f. | p-value |
|---|---|---|
| 42.29 | 6 | < 0.0001 |

**Meta-regression with Period as a Categorical Moderator:**

Mixed-Effects Model (k = 7; tau^2 estimator: REML)

tau^2 (estimated amount of residual heterogeneity): 0.8623 (SE = 0.7387)
tau (square root of estimated tau^2 value): 0.9286
I^2 (residual heterogeneity / unaccounted variability): 82.51%
H^2 (unaccounted variability / sampling variability): 5.72
R^2 (amount of heterogeneity accounted for): 0.00%

Test for Residual Heterogeneity:
QE(df = 5) = 32.9752, p-val < .0001

Test of Moderators (coefficient 2):
**QM(df = 1) = 0.2605, p-val = 0.6098**

Model Results:

| | estimate | se | zval | pval | ci.lb | ci.ub | |
|---|---|---|---|---|---|---|---|
| intrcpt | -1.4147 | 0.5726 | -2.4706 | 0.0135 | -2.5371 | -0.2924 | * |
| periodB | -0.4170 | 0.8170 | -0.5104 | 0.6098 | -2.0183 | 1.1843 | |

```
---
Signif. codes:  0 ‘***’ 0.001 ‘**’ 0.01 ‘*’ 0.05 ‘.’ 0.1 ‘ ’ 1
```

➔ There are **no significant differences** between the two subsets.

**Outliers :**

"Wang Y et al. 2020", "Zheng et al. 2020"

**Results with outliers removed:**

```
Number of studies combined: k = 5

                         proportion        95%-CI
Fixed effect model          0.2049    [0.1444; 0.2822]
Random effects model        0.1630    [0.0794; 0.3053]

Quantifying heterogeneity:
     tau^2 = 0.4789 [0.0000; 5.2371]; tau = 0.6920 [0.0000; 2.2885];
     I^2 = 57.3% [0.0%; 84.2%]; H = 1.53 [1.00; 2.51]

Test of heterogeneity:
     Q d.f. p-value
     9.37    4  0.0524
```

**Influencers :**

No decteted.

#### CHILDREN: Meta-analysis of single means

---

### Incubation period (children)

---

Incubation Period
Number of studies combined: k = 4

```
                    mean        95%-CI
Fixed effect model   6.6901 [5.4908; 8.1512]
Random effects model 6.6901 [5.4908; 8.1512]

Quantifying heterogeneity:
 tau^2 = 0 [<0.0000; <0.0000]; tau = 0 [<0.0000; <0.0000];
 I^2 = 0.0% [0.0%; 0.0%]; H = 1.00 [1.00; 1.00]

Test of heterogeneity:
    Q d.f. p-value
 0.11    3  0.9905
```

---

### Onset of symptoms to diagnosis (children)

---

Number of studies combined: k = 3

```
                    mean        95%-CI
Fixed effect model   2.9875 [2.7860; 3.2037]
Random effects model 3.2650 [1.3457; 7.9216]

Quantifying heterogeneity:
 tau^2 = 0.5815 [0.1261; 25.6104]; tau = 0.7626 [0.3551; 5.0607];
 I^2 = 92.5% [81.2%; 97.0%]; H = 3.64 [2.31; 5.75]

Test of heterogeneity:
     Q d.f.  p-value
 26.53    2 < 0.0001
```

---

### Hospital stay (children)

---

Number of studies combined: k = 3

```
                     mean          95%-CI
Fixed effect model   13.9679 [13.1118; 14.8798]
Random effects model 12.7733 [ 8.5111; 19.1701]

Quantifying heterogeneity:
 tau^2 = 0.1197 [0.0234; 5.4996]; tau = 0.3460 [0.1530; 2.3451];
 I^2 = 89.8% [72.5%; 96.2%]; H = 3.12 [1.91; 5.12]

Test of heterogeneity:
     Q d.f.  p-value
 19.53    2 < 0.0001
```

#### Asymptomatic patients (children)

Number of studies combined: k = 4

| | proportion | 95%-CI |
|---|---|---|
| Fixed effect model | 0.1426 | [0.1216; 0.1666] |
| **Random effects model** | **0.1666** | **[0.1152; 0.2348]** |

Quantifying heterogeneity:
tau^2 = 0.1035 [0.0000; 2.2479]; tau = 0.3218 [0.0000; 1.4993];
I^2 = 55.6% [0.0%; 85.3%]; H = 1.50 [1.00; 2.61]

Test of heterogeneity:

| Q | d.f. | p-value |
|---|---|---|
| 6.75 | 3 | 0.0803 |

#### ICU admissions vs all hospitalized patients with COVID 19 (children)

Number of studies combined: k = 4

| | proportion | 95%-CI |
|---|---|---|
| Fixed effect model | 0.0418 | [0.0193; 0.0879] |
| **Random effects model** | **0.0506** | **[0.0164; 0.1460]** |

Quantifying heterogeneity:
tau^2 = 0.6147 [0.0000; 13.4537]; tau = 0.7840 [0.0000; 3.6679];
I^2 = 42.0% [0.0%; 80.5%]; H = 1.31 [1.00; 2.26]

Test of heterogeneity:

| Q | d.f. | p-value |
|---|---|---|
| 5.18 | 3 | 0.1594 |

#### Discharged from all hospitalized patients with COVID 19 (children)

Number of studies combined: k = 3

| | proportion | 95%-CI |
|---|---|---|
| Fixed effect model | 0.1453 | [0.1001; 0.2062] |
| **Random effects model** | **0.1939** | **[0.0339; 0.6223]** |

Quantifying heterogeneity:
tau^2 = 2.4140 [0.3768; >100.0000]; tau = 1.5537 [0.6138; >10.0000];
I^2 = 86.4% [60.9%; 95.3%]; H = 2.72 [1.60; 4.61]

Test of heterogeneity:

| Q | d.f. | p-value |
|---|---|---|
| 14.75 | 2 | 0.0006 |

-----------------------------------------------------------------------------------------------------------

#### In hospital (children)

-----------------------------------------------------------------------------------------------------------

Number of studies combined: k = 3

| | proportion | 95%-CI |
|---|---|---|
| Fixed effect model | 0.8497 | [0.7885; 0.8956] |
| **Random effects model** | **0.8027** | **[0.3787; 0.9645]** |

Quantifying heterogeneity:
tau^2 = 2.3384 [0.3624; >100.0000]; tau = 1.5292 [0.6020; >10.0000];
I^2 = 86.1% [59.7%; 95.2%]; H = 2.68 [1.58; 4.57]

Test of heterogeneity:

| Q | d.f. | p-value |
|---|---|---|
| 14.40 | 2 | 0.0007 |
